# A systematic review of Nipah virus disease epidemiological parameters, outbreaks and mathematical models

**DOI:** 10.64898/2026.03.19.26348815

**Authors:** Tristan Naidoo, Christian Morgenstern, Patrick Doohan, Rhys Earl, Thomas Rawson, Richard J Sheppard, Joseph T Hicks, Sreejith Radhakrishnan, Rob Johnson, Anna-Maria Hartner, Lorenzo Cattarino, Kelly McCain, Anna Vicco, Natsuko Imai-Eaton, Pathogen Epidemiology Review Group, Sabine van Elsland, Anne Cori, Ruth McCabe, Sangeeta Bhatia

**Author notes:** Contributed equally (Ruth McCabe, Sangeeta Bhatia). Contributed equally (Tristan Naidoo, Christian Morgenstern). Membership of group authorship is listed in the Supplementary Information (Pathogen Epidemiology Review Group).

## Abstract

We conducted a systematic review (PROSPERO CRD42023393345) characterising the epidemiology, outbreaks and mathematical models of Nipah virus (NiV), an important public health threat in South and Southeast Asia. We extracted 243 parameters, 89 risk factors, 39 models and 23 distinct outbreaks from 119 papers. IgG seroprevalence estimates ranged from 0% to 12.5%. NiV causes severe disease, with pooled case-fatality ratio estimates ranging widely from 9.1% (95%CI: 0.2%-41.3%) in Singapore to 81.9% (95%CI: 71.9%-88.9%) in Bangladesh. NiV’s natural history is poorly characterised; we estimated a median incubation period of 8.77 days (95%CI: 7.53-10.02) based on 8 studies. Transmission parameter estimates were scarce, and all but one central estimate of the basic reproduction number were below 1. NiV mathematical models (n=39) were rarely fitted to data (n=8). All extracted information is accessible via our R package, *epireview*, a dynamic resource for informing responses to future outbreaks of NiV and related pathogens.

## Introduction

Infectious diseases pose a continuous threat to global health security, with grave societal and economic consequences, as exemplified by recent public health crises including mpox and COVID-19^1–3^. To guide pandemic preparedness, the World Health Organisation (WHO) has identified pathogens with epidemic and pandemic potential for prioritisation for research and development. Nipah virus (NiV), belonging to the Paramyxoviridae family of viruses, which includes henipaviruses, is one such priority pathogen^4^.

NiV was first identified in 1998 in Peninsular Malaysia via an outbreak in commercially-farmed pigs, which then spread nationally through infected pig movements^5^ and repeated spillover into humans, and to Singapore through exported pig meat^6,7^. Most human cases resulted from exposure to pigs^8^. The outbreak was contained through livestock movement restrictions and culling of over a million pigs^9^. Between its emergence in 1998 and effective control in 1999, 283 cases of acute encephalitis, a severe symptom of NiV, and 109 deaths were reported in Malaysia, with likely many undetected infections^10–12^. A further 11 cases and one death were reported in Singapore^13^ during this period. The outbreak resulted in long-term changes to pig-farming in Malaysia, with pig-rearing banned in some regions or outside of designated “Pig Farming Areas”. Malaysia was declared and has remained NiV-free since 2001^14^.

The next documented NiV emergence occurred in 2001 in Bangladesh^15^ and the neighbouring Indian state, West Bengal^16^, both identified retrospectively using serology. Direct spillover from bats to humans and onward human-to-human transmission were documented, suggesting distinct epidemiology and transmission dynamics compared to the first outbreak^8^. Since 2001, Bangladesh has experienced spillover almost annually, becoming the centre of global NiV activity^17^. Since 2018, isolated outbreaks have also been reported in Kerala (India)^18^, and confirmed cases were reported in West Bengal in early 2026^19^. The Philippines experienced an outbreak in 2014, with documented spillover from horses to humans and human-to-human transmission ^20^.

As of February 2026, documented NiV activity in humans is limited to India and Bangladesh. However, the virus is present in bats across South and Southeast Asia^21,22^, suggesting a broad geographical area of potential zoonotic spillover risk. This, combined with past evidence of human-to-human transmission and the lack of medical countermeasures, makes NiV a potent threat. A comprehensive understanding of NiV is therefore essential for pandemic preparedness. In particular, epidemiological parameters form crucial inputs for designing outbreak control policies and mathematical models that can be used to inform public health response and rollout strategies should pharmaceutical interventions, e.g. a vaccine, become available^23^. Despite a large body of literature, there is currently no systematic, up-to-date and dynamic collation of NiV epidemiological parameter estimates; current reviews focus only on specific aspects of its epidemiology, such as severity.

As part of a broader effort to collate a comprehensive library of the epidemiology of WHO priority pathogens, we have previously conducted systematic reviews of outbreaks, mathematical models, and epidemiological parameter estimates for five other priority pathogens^24–28^. This study presents a systematic review of published epidemiological parameter estimates, outbreaks and mathematical models of NiV. We have made the extracted data available in a user-friendly, open-source R package, *epireview*^29^, which can be easily accessed and updated by the research community.

## Methods

We followed the Preferred Reporting Items for Systematic Reviews and Meta-Analyses (PRISMA) guidelines and registered our study protocol with PROSPERO (International Prospective Register of Systematic Reviews, #CRD42023393345).

### Search strategy and selection criteria

We searched PubMed and Web of Science for studies published from database inception up to 8 March 2019 and repeated the search to include publications through 14 March 2025. Results were imported into Covidence (2024)^30^ and de-duplicated. Titles and abstracts, and then full texts, were independently screened by two reviewers (selected from TN, CM, TR, PD, JH, AM, RE, SR, LC, NI-E, RM, SB), and conflicts resolved by consensus. Non-peer-reviewed literature and non-English language studies were excluded (see appendix p.4, p.58 and Table A.1).

### Data extraction

Twelve individuals (TN, CM, TR, PD, RJ, RS, JH, AH, RE, AC, RM, SB) extracted information from the studies retained after full-text screening. 23.5% (n=28) of all included papers (n=119) meeting the inclusion criteria were randomly selected and double-extracted to validate the extraction process. A consensus on discordant results was established, after which the 12 extractors independently conducted single extraction on the remaining studies. Data on publication details, transmission model details, historical outbreaks, any epidemiological delays (e.g. incubation period or symptom-onset-to-hospitalisation or hospitalisation-to-outcome delays), case fatality ratios (CFRs), basic and effective reproduction numbers, attack rates, growth rates, overdispersion, seroprevalence, and risk factors, were extracted using Research Electronic Data Capture (REDCap) hosted at Imperial^31,32^. Each extractor also undertook a quality assessment (QA) of the paper (using a customised questionnaire; appendix p.11 and p.16). We extracted statistical uncertainty and population variability across all parameters (appendix p.14). For risk factors, we only extracted the outcome (e.g. infection), whether a factor was reported as statistically significant, and whether the analysis was adjusted for other covariates. We did not extract quantitative measures of association (e.g. odds ratios) as differences in reference groups and stratification made comparison across studies challenging.

We excluded systematic reviews from our study but used them to cross-check that all eligible studies were included (appendix p.58). Full details of the data and extraction process are provided in the appendix (appendix p.6; Tables A.2-A.4; Tables B.8-B.14).

### Data analysis

QA scores for each article were calculated as the proportion of ‘Yes’ responses to applicable questions. Temporal trends in QA scores were assessed using local polynomial regression (appendix Figure B.2).

Historical outbreaks extracted from the literature were deduplicated to account for multiple articles reporting the same information (appendix p.12, p.20-23).

Extracted CFR parameters were deduplicated ahead of meta-analysis (appendix p.13). We supplemented our severity analysis with CFR estimates obtained from two complementary data sources. The first was the deduplicated extracted historical outbreaks, providing more comprehensive information, particularly on cases in India. The second, specific to Bangladesh, was surveillance data from the Institute of Epidemiology, Disease Control and Research (IEDCR), providing a unique opportunity for temporal analyses using a single surveillance system (appendix p.14 and Table B.8).

For our primary severity analysis, we conducted a meta-analysis of deduplicated CFR using the *meta* R package^33^. We also conducted a meta-analysis of incubation period estimates. Common and random effects models were used to generate pooled estimates with 95% confidence interval (CI) and *I*^2^ heterogeneity estimates (appendix p.13-14). Unless otherwise stated, we report pooled random-effects estimates. We did not perform meta-analyses for other parameters because too few central estimates with suitable measures of population variability were available.

For each of the two complementary severity data sources, we estimated naïve CFRs (number of reported deaths divided by number of reported cases). For deduplicated outbreaks, we estimated a pooled CFR using a mixed-effect model, following the same approach used for the extracted parameter CFR meta-analysis described above. In stratified analyses, some groups may contain only a single estimate; in such cases, the naïve CFR is reported. Because IEDCR surveillance data originate from a single source, accounting for between-estimate heterogeneity was unnecessary, and we instead report naïve CFRs with binomial confidence intervals.

Analyses were conducted in *R* (version 4.2.2)^34^; curated data on outbreaks, models, and epidemiological parameters are available from the *epireview* R package^29^ (appendix p.59).

## Results

The search returned 2,863 potentially relevant articles. De-duplication retained 1,970 articles for title and abstract screening. Subsequently, 407 studies underwent full-text screening, of which 119 met the inclusion criteria (appendix, p.5). The main reasons for study exclusion were “no original estimates” (n=148) and “no reported parameters or models of interest” (n=76) (Figure 1, see appendix p.5 for Cohen’s kappa for the screening and full-text review).

**Figure 1:**
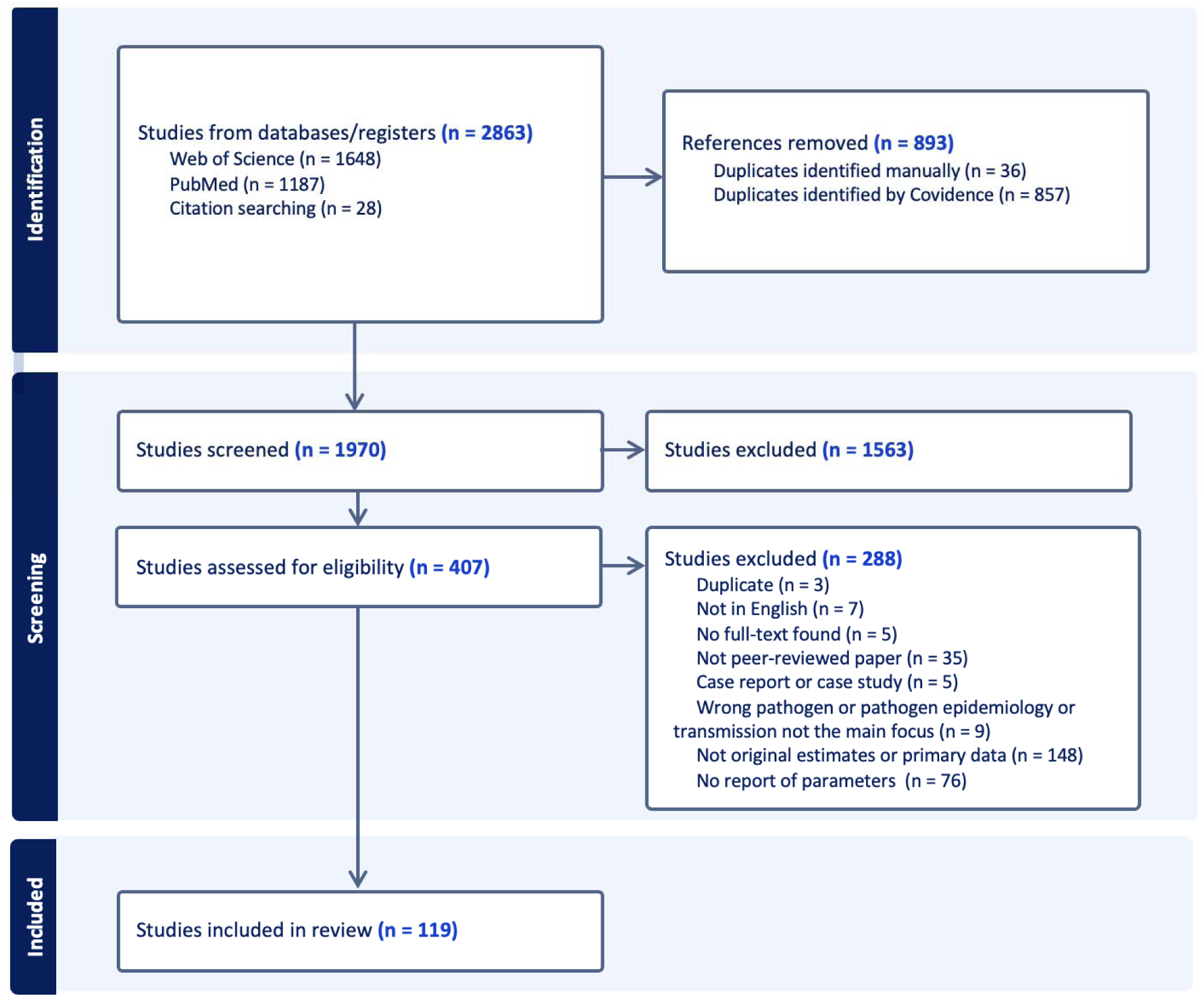
Study selection according to PRISMA guidelines and criteria described in SM-Table B.1. (Reasons for abstract exclusion not recorded in Covidence).

We extracted 243 parameter estimates and 89 risk factors from 119 articles (appendix Tables B.6-B.7). Furthermore, we extracted 66 outbreaks from 30 articles (appendix p.20-23), corresponding to 23 distinct outbreaks, totalling 656 cases and 351 deaths, after accounting for duplicated reporting. We also extracted 39 models from 39 articles (appendix p.24-26).

After deduplication of outbreaks, we identified 14 outbreaks in Bangladesh (range 1-44 cases per outbreak), six in India (1-66 cases per outbreak), and one each in Malaysia (283 cases), Singapore (11 cases), and the Philippines (17 cases) (Figure 2A).

**Figure 2:**
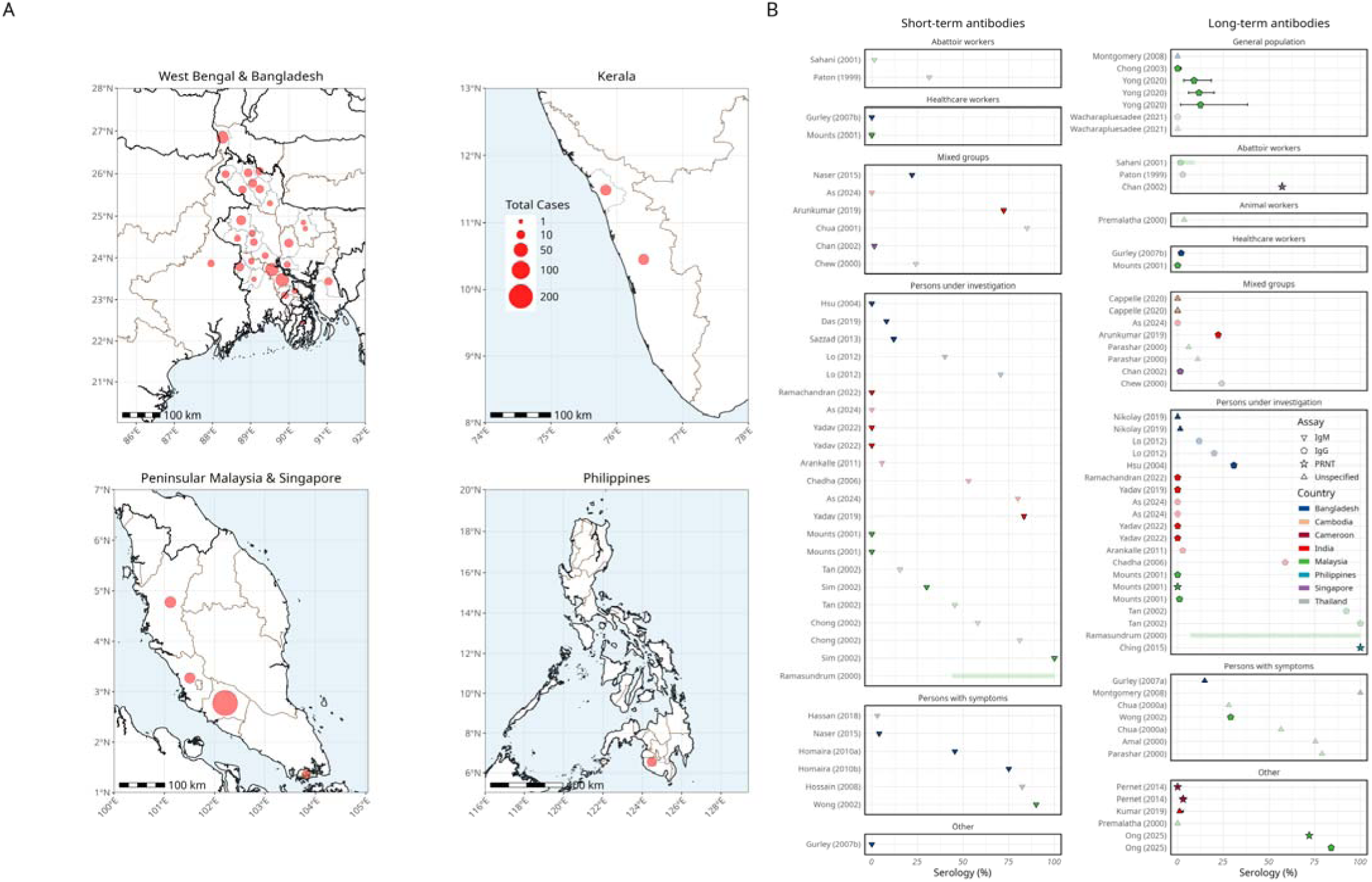
Overview of extracted outbreaks and seroprevalence estimates. (A) Map of total cases reported in West Bengal and Bangladesh (top left), Kerala (top right), Peninsular Malaysia and Singapore (bottom left), and the Philippines (bottom right). Lines in the outbreak map are coloured as follows, with country boundaries shown in black; state boundaries i countries in which outbreaks occurred, regardless of whether an outbreak occurred in that specific state; and district boundaries in grey where specified for an outbreak. (B brown for all Forest plot of seroprevalence estimates stratified by short-term antibodies (left) and long-term antibodies (right), and by population type (panels); colours represent countries; parameters with a QA score ≤ 50% are displayed with a lighter shade; inverted triangles, pentagons, stars, and triangles represent IgM, IgG, PRNT, and unspecified assay estimates respectively; black error bars indicate estimate uncertainty, and thick coloured bands indicate ranges across central estimates of disaggregated parameters (e.g. age, sex, time).

We extracted 93 seroprevalence estimates from 43 articles (Figure 2B), reported from seven countries (Malaysia n=36 estimates; Bangladesh n=22; India n=21; Singapore n=7; Cambodia n=2; Cameroon n=2; Thailand n=2; Philippines n=1). Only 7 estimates related to the general population, from studies undertaken in Malaysia (n=4 estimates), Thailand (n=2) and Bangladesh (n=1) with central estimates of IgG seroprevalence ranging between 0% and 12.5%. Ten estimates were among groups at potentially high risk of exposure: six estimates came from abattoir workers in Malaysia (n=3) and Singapore (n=3), with central estimates ranging from 1.4% (IgM)^35^ to 57.1% (neutralising antibodies)^6^; while four estimates focussed on healthcare workers in Malaysia (n=2 estimates) and Bangladesh (n=2), with only 2 of 105 individuals in Bangladesh testing positive for IgG antibodies and all others showing no evidence of seropositivity. The remaining estimates (n=55) were predominantly related to persons under investigation and were highly variable (from 0% to 100%) with no clear driver of heterogeneity, such as location, population group, or assay type. Notably, Pernet et al.^36^ reported a seroprevalence of 3% (n=7 positives of 227 tests using a neutralisation assay) among individuals exposed to bats in Cameroon, a country which has never reported cases of NiV. The studies in Thailand and Cambodia found no evidence of seropositivity^37^.

We extracted two parameters characterising disease severity: the proportion of cases who are symptomatic, and CFRs. Estimates of the proportion of symptomatic cases were scarce but consistently high, with central estimates from three studies^38–40^ ranging from 78.3% to 100% (Figure 3A). CFR estimates were more commonly reported; we extracted 64 estimates from 36 articles (appendix Table B.12), which were reduced to 17 estimates from 15 articles after deduplication. The deduplicated extracted CFR central estimates were quite variable, ranging from 9.1% to 100% (Figure 3B), with a pooled estimate of 69.4% (95%CI 53.7%-81.6%, *I*^2^: 85.9%) (Figure 3C). These findings underline both the high lethality of NiV, and high heterogeneity between estimates.

**Figure 3.**
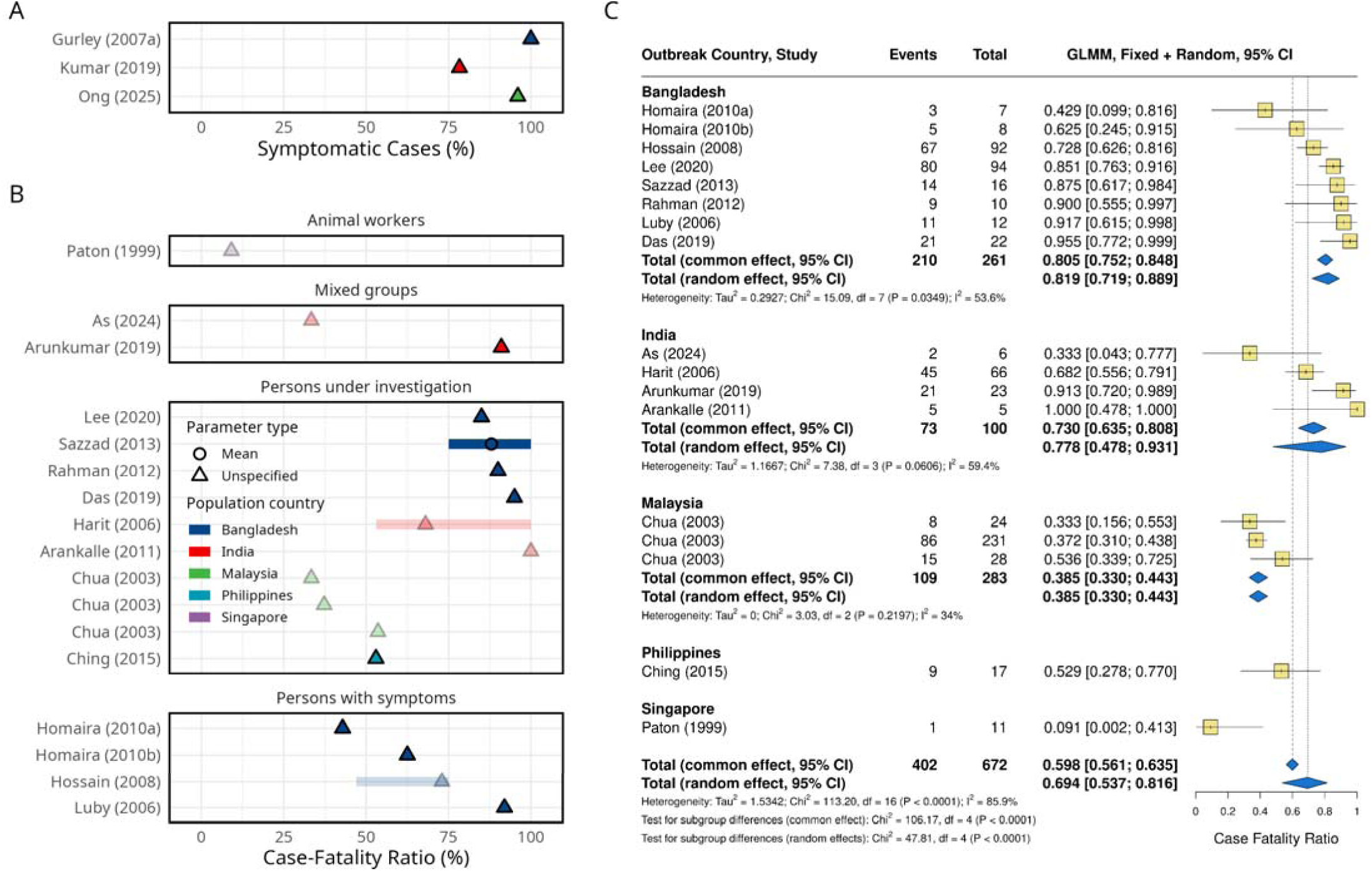
Overview of severity estimates. (A) Forest plot of extracted proportion of symptomatic cases. (B) Forest plot of deduplicated extracted CFR estimates stratified by population group. (C) Meta-analysis of deduplicated extracted CFR estimates by country. In panels (A) and (B), colours represent countries; parameters with a QA score ≤ 50% have a lighter shade; circles and triangles represent mean and unspecified estimate types respectively; and coloured bands indicate ranges across central estimates of disaggregated parameters (e.g. age, sex, time). In panel (C), squares represent individual CFR estimates, with black horizontal lines indicating 95%CIs; diamonds represent pooled estimates from common-effect and random-effects models, with widths corresponding to 95%CIs. Appendix B, Section 4.1 (p.48-56) presents additional severity results.

To explore potential drivers of heterogeneity in extracted CFR estimates, we performed multiple stratified analyses. There was no obvious variation by population group (Figure 3B, appendix Figure B.9B), but heterogeneity was evident over time and countries (appendix Figure B.8, Figure 3C). Stratification by time suggested an increase in CFRs, with pooled estimates increasing from 37.4% (95%CI: 32.1-43.1%, *I*^2^*: 50.5*%) in 1998-1999 to 72.5% (95%CI: 65.9-78.2%, *I*^2^: 10.0%) in 2000-2009 and 85.4% (95%CI: 71.0%-93.3%, *I*^2^: 68.5%) in 2010-2019. Only one deduplicated estimate was available from 2020 onwards (33.3%, India) (appendix Figure B.9A). However, these results may also reflect geographic heterogeneity in CFRs. Early outbreaks in Malaysia (pooled effect CFR estimate: 38.5%, 95%CI 33.0-44.3%, *I*^2^*: 34.0*%) and Singapore (9.1%, 95%CI: 0.2-41.3%) had significantly lower estimated CFRs than later outbreaks in Bangladesh (81.9%, 95%CI 71.9-88.9%, *I*^2^: 53.6%) and India (77.8%, 95%CI 47.8-93.1%, *I*^2^*: 59*.4%). The CFR in the Philippines was between these two extremes (52.9%, 95%CI 27.8-77.0%).

We further explored geographic heterogeneity in CFR using the estimates from the deduplicated outbreak data. This pooled CFR (69.8%, 95%CI: 57.8-79.7%, *I*^2^: 73.8%) was very similar to that obtained from the extracted parameters (Figure 3C), and stratified analyses confirmed substantial heterogeneity by country (appendix Figure B.6). The IEDCR data provided an additional opportunity to examine temporal variation in CFR. The naive CFR over the entire period (2001-2025) was 71.8% (95%CI 66.7%-76.4%), lower than estimates for this country based on either extracted parameters (Figure 3C) or outbreaks (appendix Figure B.6). We found no clear temporal trend (appendix Figure B.8). Taken together, results across all three data sources suggest that country was the most important driver of CFR heterogeneity.

We identified 69 estimates of epidemiological delay parameters reported in 30 articles (Figure 4).

**Figure 4:**
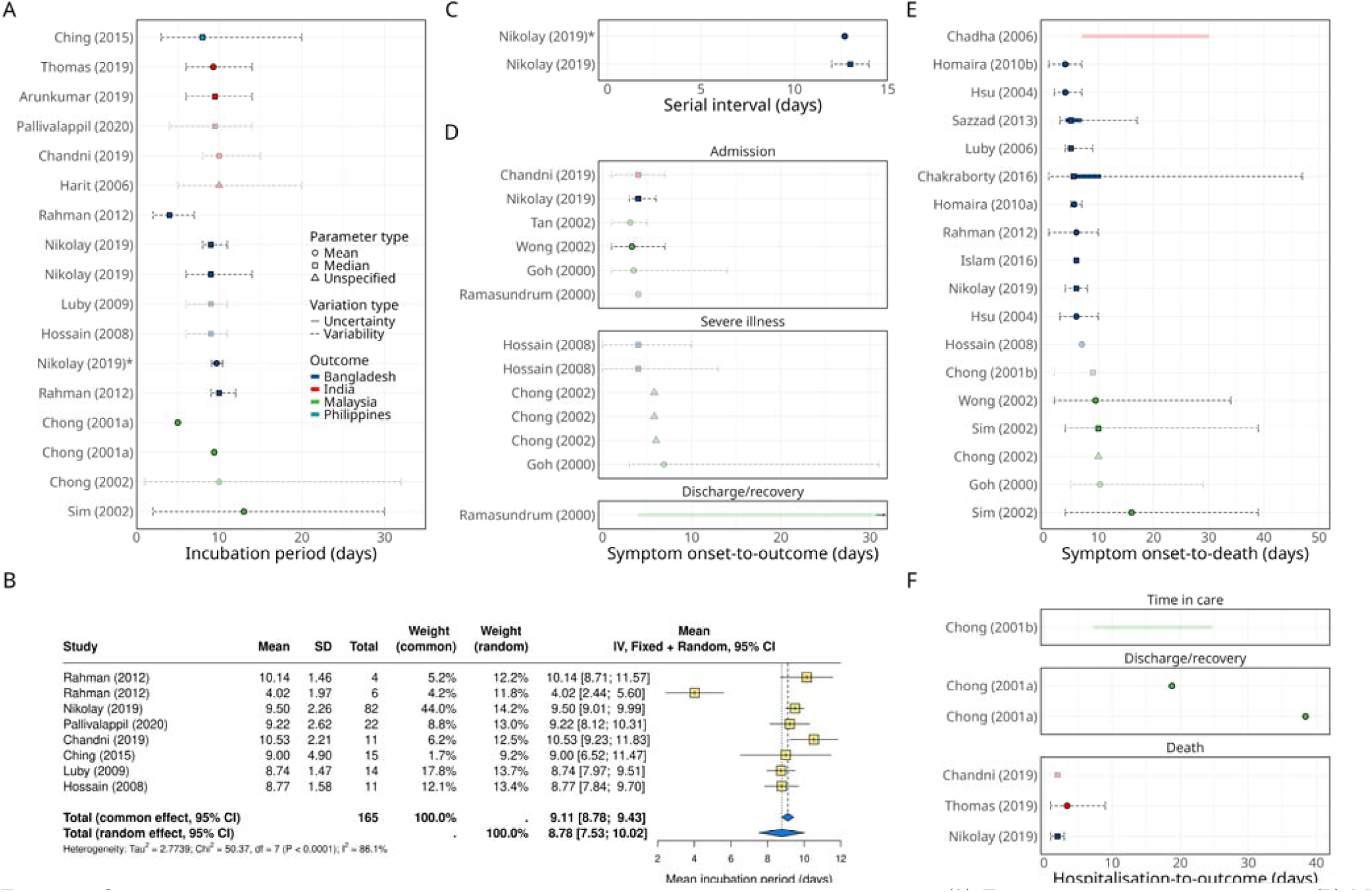
Overview of extracted epidemiolocal delays and a meta-analysis of incubation periods. (A) Forest plot of incubation periods. (B) Meta-analysis of incubation periods; only delays reported as median (interquartile range), median (range), and mean (standard deviation) are included (appendix p.14). (C) Forest plot of serial intervals. (D) Forest plot of symptom-onset to different outcomes. (E) Forest plot of symptom-onset to death. (F) Forest plot of hospitalisation to outcome. In panels (A, C-E), colours represent countries; parameters from studies with a QA score ≤ 50% are displayed with a lighter shade; circles, squares, and triangles represent mean, median, and unspecified estimate types respectively; solid and dashed black error bars indicate estimate uncertainty and population variability respectively; and thick coloured bands indicate ranges of central estimates of disaggregated parameters across different groups (e.g. age, sex, time). *indicates estimated parameters while all other parameters are observed sample statistics. In panel (D), the discharge/recovery delay range of central estimates reported by Ramasundrum (2000) extends to 83 days. In panel (B), squares represent incubation period estimates from individual studies, with black horizontal lines indicating 95%CIs; diamonds represent pooled estimates from common-effect and random-effects models, with widths corresponding to 95%CIs. Estimates from four studies that reported only population variability without central estimates, as well as a single estimate for symptom onset to death/recovery (combined), were excluded from the figure for readability.

We extracted 21 estimates of the incubation period from 16 articles (Figure 4A). Estimates were broadly consistent across location, time and population group, centring around 8-10 days (n=14 estimates). Nonetheless, some central estimates were as low as 4 days^41^, in persons under investigation in Bangladesh in 2008, and as high as 13 days^42^, among hospitalised patients in Malaysia throughout 1998-1999. Population variability in the incubation period was wider in the initial Malaysian outbreak (2 days-2 months) than in later outbreaks in India (4-20 days), Bangladesh (2-14 days), and the Philippines (3-20 days) across all estimates. The total random effect mean estimate of the median incubation period was 8.77 days (95%CI: 7.53-10.02, *I*^2^: 86.3%) (Figure 4B).

We found a single estimate of the serial interval in Nikolay et al.^43^, with an observed median of 13 days, an interquartile range of 12 to 14 days, and an estimated mean of 12.7 days and a standard deviation of 3 days, obtained by fitting a gamma distribution to observed data from cases in Bangladesh (Figure 4C).

We extracted 35 estimates of delays from symptom onset to various outcomes (Figure 4D-E). In general, estimates within countries were broadly consistent with notable variability across countries. The time between onset to admission (n=7 estimates) was very short, with central estimates ranging from 3.1-4 days in Malaysia (n=5), and consistent at 4 days for both Bangladesh (n=1) and India (n=1). In Bangladesh, two estimates of the time between onset and severe illness (appendix p.32-33) were also 4 days, suggesting that admission coincided with severe symptoms. In contrast, in Malaysia, the time from onset to severe illness was longer, ranging from 5.8-6.9 days (n=4 estimates), suggesting that typically, patients became severely ill after admission.

Estimates of onset to death were predominantly from Bangladesh (n=11), with central estimates ranging from 4-6 days, only marginally longer than the time to admission and severe illness, and were substantially lower than the corresponding estimates for Malaysia (n=6 estimates, range 9.5-16 days) and for India (n=1 estimate, central estimate 18.5 days) (Figure 4E). The time between hospital admission and recovery or death was less well characterised than the onset-to-outcome delays (Figure 4F). The two admission-to-recovery estimates were from Malaysia (18.8 days and 34.4 days) and were substantially greater than the three admission-to-death estimates (2 days in Bangladesh, 2–3.4 days in India).

Transmissibility was poorly characterised overall; all extracted estimates related to Bangladesh. We extracted five basic reproduction number estimates, with all but one of the central estimates falling below the critical threshold of 1 (range 0.2-0.88) (Figure 5A)^44–47^. A central estimate of 1.1 was estimated for a subgroup of patients over 45 years old and suffering from breathing difficulties; however, the overall estimate from this study was 0.33 (95%CI 0.19-0.59)^43^. We did not find any effective reproduction number estimates in the literature. Only one article, by Hsu et al^15^, reported attack rates, namely 1.1% for the Naogaon outbreak and 2.1% for the Meherpur outbreak (Figure 5B). Overdispersion, measured as the maximum number of secondary cases caused by an infector, was reported as 21 and 22 in two articles^38,43^ (Figure 5C), although we were unable to verify if this referred to the same individual.

**Figure 5:**
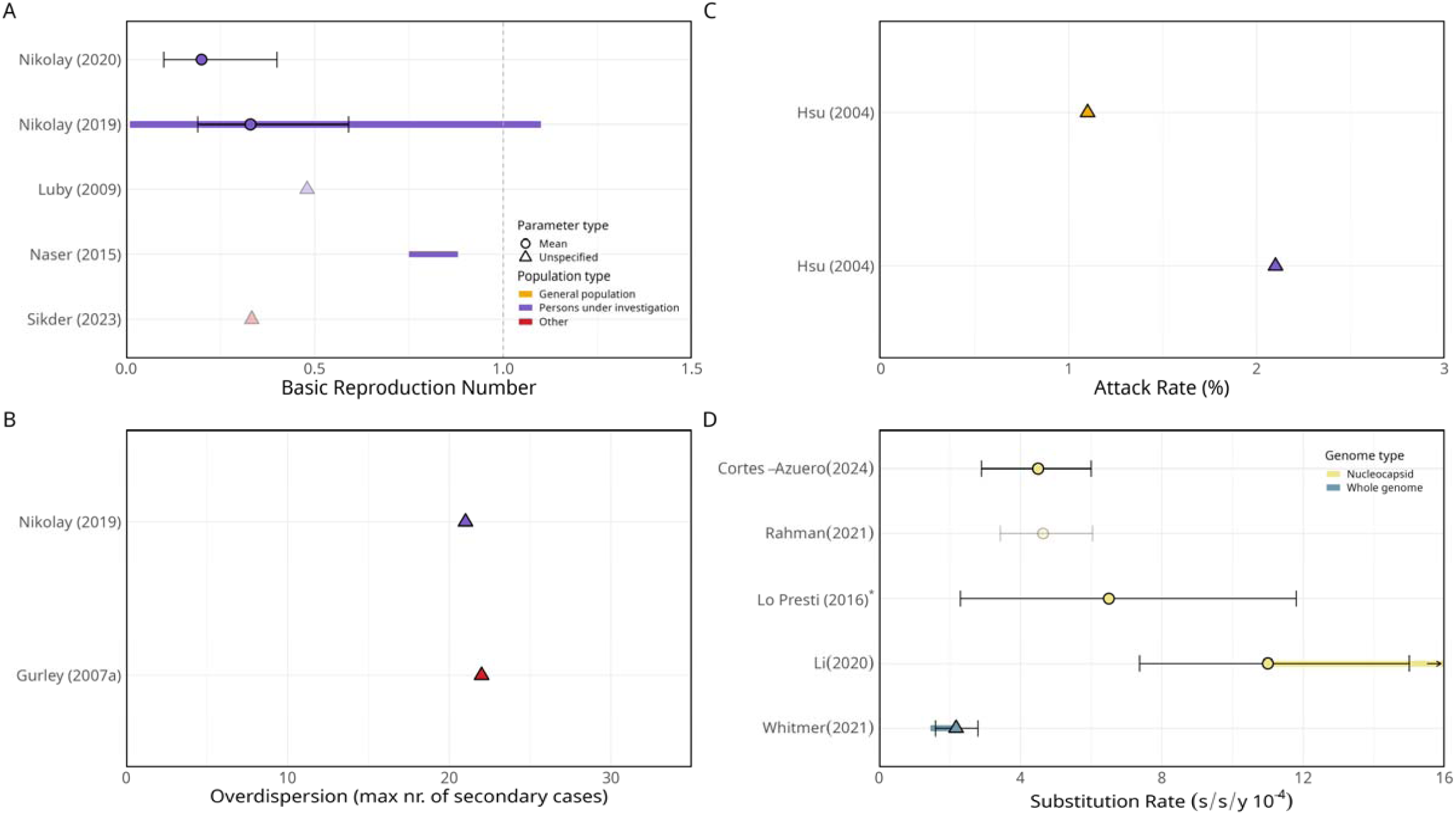
Overview of transmission and genetic parameters. (A) Forest plot of basic reproduction number. (B) Forest plot of attack rates. (C) Forest plot of overdispersion, where the x-axis represents the maximum number of secondary cases caused by an infector. (D) Forest plot of the nucleotide substitution rates, reported in substitutions per site per year (s/s/y). Panels (A-C) are stratified by population type. Parameters with a QA score ≤ 50% are displayed with a lighter shade; circles and triangles represent mean and unspecified estimates respectively; and black error bars indicate estimate uncertainty, and thick coloured bands indicate ranges across central estimates of disaggregated parameters (e.g. age, sex, time). *Lo Presti (2016) reported this metric as an evolutionary rate; a term often used interchangeably with substitution rate in viral phylogenetic analyses. The range of substitution rate central estimates reported by Li (2020) extends to 143 × 10^−4^ s/s/y.

Nucleotide substitution rates were reported in five articles^48–52^, with substantial variation in central estimates, ranging from 2.18 × 10^−4^ to 11.0 × 10^−4^ substitutions per site year (s/s/y) (Figure 5D). We note that the highest estimate, from Li et al. ^50^, is almost twice the next largest estimate of Lo Presti et al.^48^ , has wide uncertainty (95% highest posterior density interval of 7.37 × 10^−4^ to 15.0 ×10^−4^ s/s/y), and an extremely wide range of disaggregated central estimates (11.0 × 10^−4^ to 143.0 ×10^−4^ s/s/y ).

Risk factors were recorded for six outcomes, with information predominantly coming from Bangladesh and Malaysia: spillover risk (Bangladesh (n=17) and Thailand (n=1)), infection ( Bangladesh (n=30) and Malaysia (n=6)), seropositivity (Malaysia (n=5) and Bangladesh, Cameroon and Singapore (n=2 each)), death (Malaysia (n=6) and Bangladesh (n=3)), neurological symptoms (n=1 from Malaysia), and other (Bangladesh (n=6), Malaysia (n=7) and India (n=1); for example, isolation of the virus from samples or risk of specific clinical manifestations including seizures) (Table B.14 and B.16).

Contact with animals was considered as a risk factor for spillover in only one study in Bangladesh, which found a significant association^53^. Environmental variables such as rainfall or temperature were frequently considered as potential risk factors for spillover (n=4; n=5; n=10 estimates, respectively), but articles reported both significant and non-significant associations.

Close contact was often found as a significant risk factor for infection (n=11 articles reporting it as significant versus n=4 reporting it as not significant, all from Bangladesh). Contact with animals was more often found not significant (n=12) than significant (n=6) for infection as reported from studies in Bangladesh and Malaysia. Sex (n=10 parameters), occupation (n=9), age (n=5), and environmental factors (n=3) were reasonably balanced in terms of being reported as significant or not significant for infection.

Sex and close contact were found as significant risk factors for seropositivity across all studies, whereas the environment was a significant factor in the study from Cameroon. Age and contact with animals were reported as risk factors for serology, with articles noting both significant (age: n=2 from Bangladesh and Malaysia; contact with animals: n=3 from Bangladesh, Cameroon and Singapore) and non-significant associations (age: n=3; contact with animals: n=4, both from Malaysia and Singapore).

Although age was significantly associated with death from NiV (n=4 from Bangladesh and Malaysia), contact with animals (n=1 from Malaysia) and sex (n=1 from Bangladesh) were not significant^54,55^. Full details of the remaining risk factors are provided in the appendix Table B.14.

Mathematical models for NiV transmission began to be published from 2020 onwards (Figure B.4 and Table B.9). All 39 extracted models were compartmental models, except one, which used a branching process^56^; 35 were deterministic, while the remaining four were stochastic. 20 models included zoonotic spillover, and three had spatial structure. However, only eight models were fitted to data, and only six models considered uncertainty (e.g. through multiple stochastic runs or sensitivity analyses). 13 papers specified the coding language used, with six articles using Python, five using MATLAB, and two using R. Only three articles published their code alongside the articles.

Our temporal analysis of QA scores (excluding mathematical modelling studies) showed that the mean QA score assigned by extractors steadily increased since 1999 (appendix p.16). Overall, the data extractors tended to assign lower quality scores to modelling studies, possibly because many of these studies used transmission models that were not fitted directly to data (appendix p.24-26).

We assessed the risk of publication bias associated with the meta-analyses presented here to be low (appendix Figures B.10, B.17) since we conducted meta-analyses on observed disease parameters (severity and the median incubation period), which are themselves less likely to be subject to selective reporting of positive results.

## Discussion

In this systematic review, we have collated and analysed epidemiological parameters, outbreaks and mathematical models of NiV to provide a comprehensive overview of NiV epidemiology.

After the initial outbreaks of NiV in Malaysia and Singapore in 1998, reported cases in humans have been confined to South and Southeast Asia, with recurrent outbreaks in Bangladesh (since 2001) and Kerala, India (since 2018) and sporadic outbreaks in West Bengal^57^, India (since 2001), and a single documented outbreak in the Philippines (2014)^58,59^. The outbreaks in South Asia have been notably smaller than in the initial Malaysian outbreak, often reporting less than 30 cases, with some notable exceptions, for example, the 2001 outbreak in West Bengal which recorded 66 cases^9,60^.

Despite the temporal proximity of the 1998-1999 outbreak in Malaysia and the 2001 outbreak in Bangladesh, extensive evidence, including genomic investigations^61^, suggests that these two outbreaks were unlinked, distinct events, caused by distinct NiV strains, NiV Malaysia (NiV-M)^62^ and NiV Bangladesh (NiV-B)^16,61^. These differences are reflected in parameter estimates, particularly transmissibility, epidemiological delays and severity, for which we found substantial spatiotemporal heterogeneity. However, it is challenging to disentangle the potential contributions of time, transmission route, strain-specific characteristics and local factors (e.g. related to disease surveillance, health care systems or environmental conditions) as drivers of the observed heterogeneity^28,63,64^.

The transmissibility of NiV (reproduction numbers, attack rates, and overdispersion) was generally poorly characterised in the literature. The limited parameter estimates were all from Bangladesh, despite Malaysia reporting the largest documented NiV outbreak to date^9^. The estimates of the basic reproduction number, nearly all below the critical threshold of one, suggest a lack of sustained human-to-human transmission. However, when there was human-to-human transmission, extracted overdispersion estimates suggest high potential for superspreading, presenting a challenge for outbreak control^65,66^.

Seroprevalence studies can shed further light on the transmissibility of NiV, with the low estimates in general populations supporting a lack of widespread transmission, potentially due to a variety of factors including localised risk of spillover, limited onward transmission chains, or the time elapsed since exposure. However, only seven of 93 seroprevalence estimates were undertaken in the general population, and did not cover all areas with reported cases, namely India, Singapore or the Philippines. Of the limited evidence among high-risk groups, seropositivity appears to be higher among abattoir workers than in healthcare workers, but further studies are required before any strong conclusions can be drawn regarding this risk.

The 89 extracted risk factors were predominantly evaluated for Bangladesh, with infection and spillover risk the outcomes most frequently considered. There was a lack of conclusive evidence: many risk factors, including sex and environmental factors, presented as significant an equal number of times as they presented as non-significant. Age was consistently associated with mortality; close contact with infected individuals was unsurprisingly found to be a driver of infection across multiple studies. Throughout, contact with animals was often not found to have a significant association with infection or serology, despite the documented role of zoonotic spillover of NiV. This may be attributable to study design, including definitions of animal contact that are too broad or unspecific^67^.

We found paucity of data on the natural history of NiV, with only one estimate of the serial interval, and a small number of estimates for the time between key milestones, including symptom onset, hospital admission and recovery. The only exceptions were the incubation period (21 estimates) and time between symptom onset and death (18 estimates). Parameters estimates were generally consistent within country and exhibited substantial heterogeneity across countries. In Malaysia, severe illness generally occurred after hospitalisation, whereas in Bangladesh, the evidence suggests that these two events coincided. This may imply that hospitalisation in Bangladesh is potentially triggered by severe symptoms, but it could also signify different clinical courses of NiV in these locations.

Our results underline the high severity of NiV. Available estimates of the proportion of symptomatic cases, while sparse, suggest that more than three-quarters of infections are symptomatic^38–40^. Furthermore, the disease progression tends to be rapid (4-7 days from onset to severe disease) often culminating in a fatal outcome. Our overall meta-analysis of CFR estimates yielded a high pooled estimate of 69.4% (95%CI 53.7-81.6%). Since multiple studies can report CFR that rely on the same data, we deduplicated the published estimates to ensure a robust and unbiased meta-analysis. While most published estimates of the overall CFR were consistent with our estimate, our deduplication of estimates likely explains any differences^68–70^.

Consistent with other parameters, we found that geography was the primary driver of heterogeneity across CFR estimates, with lower estimates for the outbreaks in Malaysia and Singapore (38.5% and 9.1%, respectively) than for outbreaks in India and Bangladesh (77.8% and 81.9%, respectively). The CFR for the outbreak in the Philippines (52.9%) was between these two extremes, although this must be considered in the context of an isolated outbreak with a distinct spillover mechanism. Temporal trends in CFR could serve as an alternative explanation for these differences, with initial outbreaks having a lower CFR. To investigate these alternative hypotheses, we also analysed IECDR data^71^, which covers each year since 2002. This analysis did not show a significant temporal association with the CFR (appendix p.49). While preliminary, these results suggest that geographic differences are a plausible explanation for the observed heterogeneity. Disentangling the drivers of these differences across geographies is challenging, as observed severity can reflect a combination of intrinsic virulence, potentially influenced by viral lineage, and contextual factors including disease surveillance, diagnostic capacity, access to healthcare, and therapeutic options, all of which warrant further investigation^72–74^.

The heterogeneity across countries in key parameters is reflected in the distinct dynamics and the host species. Three different species have played a key role in spillover across different groups of outbreaks: pigs in Malaysia and Singapore, horses in the Philippines and bats in Bangladesh and India, all of which are present globally. Serosurveys conducted in animals indicate the presence of anti-Nipah antibodies in *Pteropus medius* bats in Indian states that have so far not reported any Nipah activity in humans^75,76^. Similar findings from other regions in the world^77^ outside the “Nipah belt” suggest a wider geographical zone with potential spillover risk mediated by seasonal dynamics, viral shedding in bats, bat behaviour, and local human practices such as the consumption of contaminated fruits. For example, the 2014 study by Pernet et al. found NiV cross-neutralising antibodies in 3-4% of 227 bat-exposed individuals in Cameroon, a region where no NiV activity in humans has yet been reported^78^. The symptoms of NiV disease are clinically similar to other pathogens that cause febrile illness and so it is plausible that, as with the Malaysian outbreak, spillover incidents are misdiagnosed or undetected^58^. The fact that NiV is characterised by recurring spillover events from infected hosts underlines it as a canonical example of the need for a One Health approach to surveillance.

Mathematical modelling is increasingly being used to answer key policy questions that arise during public health crises, ranging from an assessment of severity to that of the efficacy of interventions, or designing clinical trials^79,80^. Our review identified only 39 mathematical modelling studies of NiV, all of which were published since 2020 despite most reported cases and outbreaks occurring far before this period. Only eight (of 39) studies were fitted to data and only three had available code. It is noteworthy that all models in the review were published since 2020, similar to findings from our related reviews on other pathogens^25,26^, with one hypothesis being that the attention modelling received during the COVID-19 pandemic inspired a surge in its use across other pathogens. However, the studies with mathematical models in our review generally had low QA scores indicating an important knowledge gap deserving of further research; we consider that this review provides a strong foundation upon which to begin addressing this.

Our study has some limitations. First, we excluded literature that was not in English or not peer reviewed. This includes information on most of the outbreaks that have occurred in India^81,82^, which are only documented in the grey literature. However, we do not expect to have missed many parameter estimates or models, which are predominantly published in peer-reviewed journals. Second, this review focused on human hosts and excluded studies focused on animal hosts only, to limit the scope of our research. Third, we did not extract all disaggregated data from studies on factors such as age, region, or time due to limited resources; we have noted in the associated *epireview* database all instances where disaggregated data is available so that researchers can access and/or extract it in the future. Fourth, we extracted risk factors without quantitative estimates because the underlying methods and group definitions varied widely. Information on risk factor studies are annotated in *epireview* to make them easier to find. Fifth, despite our best efforts to achieve concordance among data extractors, some discrepancies may remain. We have extensively documented our data extraction guidelines for transparency.

In this study, we have provided a comprehensive overview of past outbreaks, key epidemiological parameters, and mathematical models for NiV. Our work confirms that NiV is a concerning pathogen with potential for causing outbreaks with high severity. We have also highlighted critical knowledge gaps - we note that a large body of evidence is drawn from the outbreaks in Bangladesh. Taken together with the observed spatial variation in NiV epidemiology, it suggests a blind spot in our understanding of this virulent virus. The data extracted in this study, and in our previous works for Marburg, Ebola, Lassa, SARS, and Zika are available to the wider community via our open-source R package *epireview* to inform future research and public health responses. *epireview* enables easy updating of our data library, turning the current collation into a living and evolving resource for the public health research community, which we hope will be expanded upon as the community continues to monitor and research this highly lethal pathogen.

## Data Availability

All data are available from: https://github.com/mrc-ide/epireview/tree/main/data.

## Declarations

### Availability of data and materials

https://github.com/mrc-ide/epireview/tree/main/data

### Code availability

https://github.com/mrc-ide/epireview; https://github.com/mrc-ide/priority-pathogens

### PROSPERO

CRD42023393345 (https://www.crd.york.ac.uk/prospero/display_record.php?RecordID&RecordID=393345)

### Authors’ contributions

SB, SE, AC, and NI conceptualised this systematic review. TN, CM, TR, SR, LC, and SB searched the literature and screened the titles and abstracts. TN, CM, TR, PD, RJ, RS, JH, AH, RE, AC, RM, and SB reviewed all full-text articles. TN, CM, TR, PD, RJ, RS, JH, AMH, RE, AC, RM, and SB extracted the data. TN, CM, RM, and SB formally analysed, visualised, and validated the data. TN, CM, and SB were responsible for the software infrastructure. AC acquired funding. CM, SB, RM, and AC were responsible for project administration. TR and SE were responsible for training individuals to use Covidence and for designing the REDCap form. CM, RM, and SB supervised the systematic review. TN, CM, RM, and SB wrote the original manuscript draft. All authors were responsible for the methodology, review, and editing of the manuscript. All authors debated, discussed, edited, and approved the final version of the manuscript. All authors had final responsibility for the decision to submit the manuscript for publication.

