## Appendix for "A systematic review of Nipah virus disease epidemiological parameters, outbreaks and mathematical models"

**C Other Systematic Reviews** 58

**D Excluded Studies** 58

**E Software and epireview** 59

**F PRISMA 2020 Checklists** 60

**G Pathogen Epidemiology Review Group (PERG) Membership** 63

G.19 Pathogen Epidemiology Review Group (PERG) membership as of Janaury 2025began . 64

### Overview

This supplement provides additional details of the methodology used in our literature review (section A) as well as additional results (section B), including full details of the information extracted as part of the review and synthesised in the main text. Section E introduces our R package `epireview`, where all the data gathered in this review is stored. Section C details other Nipah virus systematic reviews identified in our initial literature search and which we used to ensure no relevant study was omitted in our review. Section D lists all the studies we excluded. Section F contains our PRISMA checklists. Finally, section G lists all members of the Pathogen Epidemiology Review Group who contributed to this work.

### A Additional Methods

This systematic review and meta-analysis of Nipah virus is part of a series of systematic reviews of nine priority pathogens on the WHO's R&D blueprint list, which was registered on PROSPERO (International Prospective Register of Systematic Reviews, CRD42023393345). Other systematic reviews in this series completed to date studied Marburg virus disease [1], Ebola virus disease [2], Lassa fever [3], SARS-CoV-1[4], and Zika virus disease [5].

The code to reproduce our results is available at `https://github.com/mrc-ide/priority-pathogens`.

#### A.1 Search strategy and screening

##### A.1.1 Search term

The search term used to identify relevant studies to include in this review is given below. This search term is common to all pathogen systematic reviews in the series (apart from the first field containing the pathogen name). The search was initially conducted on 8 March 2019 and subsequently repeated to include publications up to 14 March 2025.

```
Nipah AND (
(transmissi* OR epidemiolog*)
 OR (model* NOT imag*)
 OR (severity OR ``case fatality ratio*'' OR CFR OR ``case fatality rate*'' OR ``mortality
     ↪ rate*'' OR ``attack rate*'')
 OR (``infectious period*'' OR ``serial interval*'' OR ``incubation period*'' OR ``generation
     ↪  time*'' OR ``generation interval*'' OR ``latent period*'' OR latency)
 OR (heterogeneit* OR superspread* OR ``super spread*'' OR superspread* OR overdispersion OR
     ↪ overdispersed OR over-dispersion OR over-dispersed OR ``over dispersion'' OR ``over
     ↪ dispersed'')
 OR (infectivity OR infectiousness OR ``growth rate*'' OR ``reproduction number*'' OR ``
     ↪ reproductive number*'' OR R0 OR ``reproduction ratio*'' OR ``reproductive rate*'')
 OR (``pre-existing immunity'' OR serological OR serology OR serosurvey*)
 OR (evolution* OR mutation* OR substitution*)
 OR (outbreak* OR cluster* OR epidemic*)
 OR (``risk factor*'')
 )
```

##### A.1.2 Screening

Papers identified in the search using the term given in the preceding Section were imported into *Covidence*, a software program used to manage systematic reviews [6]. Original research papers in English were included if reporting on Nipah transmission, evolution, natural history, severity, seroprevalence, size of previous outbreaks or published mathematical transmission models. Non-peer-reviewed literature was excluded. The full inclusion and exclusion criteria is given in Table A.1.

Using this criteria, across a team of twelve reviewers, two independent reviewers first screened titles and abstracts, followed by full-text reviews to assess eligibility for data extraction. Disagreements in eligibility determination were resolved by consensus between the two independent reviewers. The concordance between each unique reviewer pair is shown in Figure A.1.

**Screening**

| Reviewer A | Reviewer B | n | Proportionate Agreement | Yes Probability | No Probability | Random Agreement Probability | Cohen's Kappa |
|---|---|---|---|---|---|---|---|
| P1 | P2 | 1009 | 92% | 5% | 60% | 65% | 76% |
| P3 | P4 | 562 | 84% | 5% | 59% | 65% | 54% |
| P5 | P4 | 387 | 88% | 1% | 78% | 79% | 41% |
| P1 | P6 | | *too few votes to calculate Cohen's Kappa* | | | | |
| P5 | P3 | | *too few votes to calculate Cohen's Kappa* | | | | |

**Full texts**

| Reviewer A | Reviewer B | n | Proportionate Agreement | Yes Probability | No Probability | Random Agreement Probability | Cohen's Kappa |
|---|---|---|---|---|---|---|---|
| P1 | P6 | 2 | 100% | 25% | 25% | 50% | 100% |
| P7 | P1 | 5 | 100% | 16% | 36% | 52% | 100% |
| P1 | P2 | 265 | 96% | 8% | 51% | 59% | 91% |
| P5 | P3 | 94 | 76% | 24% | 26% | 50% | 51% |
| P3 | P4 | 18 | 89% | 0% | 89% | 89% | 0% |
| P5 | P4 | 14 | 64% | 3% | 67% | 70% | -21% |
| P1 | P9 | | *too few votes to calculate Cohen's Kappa* | | | | |
| P1 | P10 | | *too few votes to calculate Cohen's Kappa* | | | | |
| P8 | P2 | | *too few votes to calculate Cohen's Kappa* | | | | |
| P11 | P2 | | *too few votes to calculate Cohen's Kappa* | | | | |
| P10 | P2 | | *too few votes to calculate Cohen's Kappa* | | | | |
| P1 | P8 | | *too few votes to calculate Cohen's Kappa* | | | | |
| P5 | P12 | | *too few votes to calculate Cohen's Kappa* | | | | |

Figure A.1: Cohen's Kappa for screening and full text review. P1-P12 are members of PERG (see supplement G), and we have provided who participated at each stage in the methods section of the main text. $n$ is the number of studies considered by each Reviewer A/B pair. We highlight that *all* studies were screened by two people and any conflicts were resolved by discussion and mutual agreement of Reviewer A and B. The same applies to the subsequent full text reviews.

##### A.1.3 Inclusion and exclusion criteria

| Inclusion | Exclusion |
|---|---|
| Measures/estimates of human: Reproduction numbers (R, $R_0$, $R_t$, r, $R_e$), growth rate (r), doubling times, generation time, serial interval, incubation/latent period, case fatality ratio (CFR), attack rate, mutation rate (e.g. from phylogenetic study), overdispersion, risk factors (risk and the measure). | Non-English language publication |
| Mention of historical or any outbreak in humans: size, year, location, duration, spatial scale | Studies of co-infections. (local, regional, national, international). |
| Measures/estimates of animal: R, $R_0$, $R_t$, r, $R_e$, growth rate, mutation rate. | Animal studies that do not report R, $R_t$ etc. |
| Mathematical or statistical model of transmission. | Qualitative studies, e.g., KAP studies. |
| Measures of seroprevalence and negative seroprevalence in humans. | Pathogen not the primary focus of study. |
| Relative ratio of human-human vs animal introductions. | Duplicates. |

Continued on next page

Continued from previous page

| Inclusion | Exclusion |
|---|---|
| Reviews that report inclusion criteria for reference checking. | Does not match any of the inclusion criteria. |
| For "small" pathogens[1], include case reports to potentially reconstruct serial interval distribution etc. | In-vitro studies. |
| | Non-peer reviewed publications, conference proceedings, abstracts, posters, letters to the editor |

Table A.1: Inclusion and exclusion criteria for papers.

#### A.2 Extraction Fields

##### A.2.1 Outbreak extraction fields

"Outbreaks" or "epidemics" were extracted from studies only if described as such by the study authors. We note that many outbreaks are reported in non-peer-reviewed literature, such as reports or correspondences. However, these have been excluded from this review due to the inclusion/exclusion criteria (Table A.1). For each outbreak included in this review, we extracted, where available, the number of confirmed (including the mode of detection), probable, and suspected cases, as well as the number of asymptomatic cases, severe cases, and deaths. Full details of all variables, including contextual ones, are provided in Table A.2.

| Data field | Expected type | Variable name | Notes |
|---|---|---|---|
| Article ID | integer | article_id | ID to connect to article form |
| Outbreak data ID | integer | outbreak_data_id | ID assigned by database |
| Pre-outbreak baseline | Logical | pre_outbreak_baseline | Dropdown list for whether a baseline period was specified before the outbreak |
| Outbreak start | Date (day, month, year) | outbreak_start_day, outbreak_start_month, outbreak_start_year | Start date of the outbreak |
| Outbreak end | Date (day, month, year) | outbreak_end_day, outbreak_end_month, outbreak_end_year | End date of the outbreak |
| Outbreak ongoing | Logical | outbreak_ongoing | Tick if the outbreak was ongoing at time of reporting |
| Duration (months) | Numeric | outbreak_duration_months | Duration of the outbreak in months |
| Outbreak country | Character | outbreak_country | Country where the outbreak occurred (dropdown list) |
| Outbreak location | Character | outbreak_location | Subnational region or city of the outbreak |
| Number of cases confirmed | Integer | cases_confirmed | Total confirmed cases |
| Mode detection | Character | cases_mode_detection | How a case was detected (dropdown list) |
| Number of cases suspected | Integer | cases_suspected | Total suspected cases |
| Number of unspecified cases | Integer | cases_unspecified | Number of cases without classification |
| Number of asymptomatic cases | Integer | cases_asymptomatic | Number of reported asymptomatic infections |
| Asymptomatic transmission described | Logical | asymptomatic_transmission | Tick if asymptomatic transmission was described |
| Number of severe cases | Integer | cases_severe | Total number of severe cases |
| Number of deaths | Integer | deaths | Total number of deaths due to the outbreak |
| Population size of geographical area | Integer | population_size | Total population of the affected area |
| Type of cases disaggregated by sex | Character | type_cases_sex_disagg | Type of sex disaggregation (dropdown list) |

[1]The inclusion/exclusion criteria are reported as stated in the PROSPERO registration for the overall priority pathogen project. The threshold is determined on a case-by-case basis for each pathogen and Nipah is considered a "small" pathogen.

Continued from previous page

| Data field | Expected type | Variable name | Notes |
|---|---|---|---|
| Male cases | Integer | male_cases | Number of male cases |
| Proportion of cases in males | Numeric | prop_male_cases | Proportion of cases that are male |
| Female cases | Integer | female_cases | Number of female cases |
| Proportion of cases in females | Numeric | prop_female_cases | Proportion of cases that are female |
| Number of probable cases | Integer | outbreak_probable | Total probable cases (as stated in study) |
| Outbreak location type | Character | outbreak_location_type | Urban, Rural or unspecified |
| Outbreak source | Character | outbreak_source | potential source of outbreak (domestic animals, wild animals, date palm sap, etc. |
| Outbreak notes | Character | outbreak_notes | notes added by extractors |

Table A.2: Outbreak form fields: refer to epireview package documentation for dropdown options.

##### A.2.2 Model extraction fields

We extracted the information about Nipah virus human transmission models. This included model type, whether it modelled deterministic or stochastic processes, which transmission pathways were included. For compartmental models, we documented the compartmental structure of human health states (e.g., SIR, SEIR). We also extracted whether the model was spatial and whether it incorporated a mechanism of spillover. Furthermore, we extracted information on underlying model assumptions, whether the model is theoretical or fitted to data, he availability of the data used for fitting,the interventions considered, and the availability of model code (including programming language). Full details of all variables are provided in Table A.3.

| Data field | Expected type | Variable name | Notes |
|---|---|---|---|
| Article ID | integer | article_id | ID to connect to article form |
| Model data ID | integer | model_data_id | Generated ID based on extraction order |
| Model type | character | model_type | General type of model - from dropdown list |
| Stochastic or deterministic | character | stoch_deter | Stochastic or deterministic model as reported |
| Transmission route | character | transmission_route | Transmission route(s) modelled - from dropdown list |
| Assumptions | character | assumptions | General assumptions for the model - from dropdown list |
| Compartmental type | character | compartmental_type | Specific type of compartmental model - from dropdown list |
| Theoretical model | logical | theoretical_model | Tick box whether the model was fitted to data (TRUE) or just theoretical (NA) |
| Intervention type | character | interventions_type | Type of intervention(s) modelled - from dropdown list |
| Code available | logical | code_available | Tick box whether code for model was publicly available and reported in the paper |
| Model compartmental other | character | model_compartmental_other | If the compartmental model is not listed in the compartmental type, field to provide the model type used |
| Model uncertainty | logical | model_uncertainty | If model considers uncertainty |
| Model spatial | logical | model_spatial | if model has spatial component |
| Model spillover | logical | model_spillover | if zoonotic spillover is considered in the model |
| Model fitting method | logical | model_fitting_method | if the model is fitted to data |
| Model language | character | model_language | Programming language the model is written in. |
| Model data | character | model_data | If model data is available, and if so, where. |
| Model README | logical | model_readme | If the readme file for the model has been provided. |
| Model notes | character | model_notes | Additional notes on model captured by extractor. |

Table A.3: Model form fields: refer to epireview package documentation for dropdown options.

##### A.2.3 Parameter extraction fields

For each parameter, we extracted all available information, including types of value estimated (e.g., mean, median, standard deviation), uncertainty intervals (capturing the precision of estimates), and ranges (if, for example, multiple estimates were obtained from different populations or using different methods). Study context for parameter values included survey location and dates, sample size, basic demographic information, and timing of the survey in relation to reported outbreaks. See Table A.4 for full details.

For genomic data, if specified we noted the gene studied and if newly sequenced data were available. For reproduction numbers, we recorded the methods used for estimation (e.g. renewal equations, compartmental models, empirical methods). For case fatality ratios, we extracted whether the estimation approach accounted for cases with unknown final status or not [7]. For both case fatality ratios and seroprevalence, we additionally recorded numerators and denominators where available.

For risk factors, we extracted the outcome (e.g. infection or death), the risk factor for that outcome (e.g. age, sex or occupation), the type of occupation if specified, and whether the risk factor(s) estimates were statistically significant and/or adjusted. We chose not to extract numerical odds ratio estimates because studies may have used different stratifications or reference groups, making it challenging to compare values across studies. The information we extract offers an overview of risk factors explored across studies that may affect the risk of infection and death, which may be useful to consider when designing Nipah transmission models.

For papers which reported multiple parameter values for the same parameter (e.g. for time periods, assumptions, and sub-groups), we applied the 'rule of three', that is, if *more than* three parameter values were reported, we only captured the range of values reported for this parameter. We applied this rule as a pragmatic decision given the wide scope of our review. As an exception to this rule we did not apply the rule when parameters were disaggregated by location up to admin level 2 (sub-regions) of a country. However, we did apply the rule to estimates by neighbourhood for example.

| Data field | Expected type | Variable name | Notes |
|---|---|---|---|
| Article ID | integer | article_ID | ID to connect to article form |
| Parameter data ID | integer | parameter_data_ID | ID assigned by database |
| Parameter type | character | parameter_type | category of parameter - see dropdown list |
| Parameter value | numeric | parameter_value | central parameter value |
| Parameter exponent | integer | exponent | parameter value exponent (base 10) |
| Inverse parameter | logical | inverse_param | tick box to indicate that only inverse of parameter is reported (e.g., recovery rate from fitted model instead of infectious period) |
| Parameter unit | character | parameter_unit | units for parameter value, applies to central estimate and ranges/uncertainty intervals - see dropdown list |
| Parameter value type | character | parameter_value_type | type of central parameter value - see dropdown list |
| Parameter lower bound | numeric | parameter_lower_bound | lower bound of the parameter range if a range was reported or if data are disaggregated |
| Parameter upper bound | numeric | parameter_upper_bound | upper bound of the parameter range if a range was reported or if data are disaggregated |
| Statistical approach | character | statistical_approach | Parameter estimated or observed - see dropdown list |
| Parameter uncertainty - single type | character | parameter_uncertainty_singe_type | type of uncertainty for central parameter value if single value was reported - see dropdown list |
| Parameter uncertainty - single value | numeric | parameter_uncertainty_single_value | value for uncertainty for central parameter value if a single value was reported (e.g. value of std. dev.) |

Continued on next page

Continued from previous page

| Data field | Expected type | Variable name | Notes |
|---|---|---|---|
| Parameter uncertainty paired type | character | parameter_uncertainty_type | type of uncertainty for central parameter value if paired values were reported - see dropdown list |
| Parameter uncertainty - lower value | numeric | parameter_uncertainty_lower_value | lower bound for uncertainty for central parameter value if paired values were reported |
| Parameter uncertainty - upper value | numeric | parameter_uncertainty_upper_value | upper bound for uncertainty for central parameter value if paired values were reported |
| Distribution type | logical | distribution_type | type of distribution for estimated parameter - see dropdown list |
| First distribution parameter type | logical | distribution_par1_type | type of value for first distribution parameter - see dropdown list |
| First distribution parameter value | logical | distribution_par1_value | value for first distribution parameter (e.g. shape or scale parameter for a gamma distribution) |
| First distribution parameter uncertainty | logical | distribution_par1_uncertainty | tick box for whether uncertainty is estimated for the first distribution parameter (true) or not (false) |
| Second distribution parameter type | logical | distribution_par2_type | type of value for second distribution parameter - see dropdown list |
| Second distribution parameter value | logical | distribution_par2_value | value for second distribution parameter (e.g. shape or scale parameter for a gamma distribution) |
| Second distribution parameter uncertainty | logical | distribution_par2_uncertainty | tick box for whether uncertainty is estimated for the second distribution parameter (true) or not (false) |
| Disaggregated data available | logical | method_disaggregated | tick box if disaggregated estimates are available (true) or not (false) |
| Parameter estimates disaggregated by | character | method_disaggregated_by | categories for disaggregation of parameter estimates |
| Only disaggregated data available | logical | method_disaggregated_only | tick box if only disaggregated estimates are available (true) or if a central estimate is also available (false) |
| Is parameter from supplement? | logical | method_from_supplement | tick box for whether parameter was extracted from supplement (true) or not (false) |
| Is parameter in a figure only? | logical | parameter_fromfigure | tick box to indicate that parameter is plotted in a figure but not reported numerically in the text/table |
| Study population country | character | population_country | country of the survey population - see dropdown list |
| Study population location | character | population_location | region/district/province/city of the survey population - see dropdown list |
| Start day of study | integer | population_study_start_day | study start day |
| Start month of study | character | population_study_start_month | study start month - see dropdown list |
| Start year of study | integer | population_study_start_year | study start year - see dropdown list |
| End day of study | integer | population_study_end_day | study end day |
| End month of study | character | population_study_end_month | study end month - see dropdown list |
| End year of study | integer | population_study_end_year | study end year - see dropdown list |
| Survey timing related to outbreak | character | method_moment_value | timing of the survey in relation to the outbreak, if specified in paper - see dropdown list |
| Study population sample size | integer | population_sample_size | sample size of the population used for parameter estimation |
| Study population minimum age (years) | numeric | population_age_min | minimum age of the survey population in years |
| Study population maximum age (years) | numeric | population_age_max | maximum age of the survey population in years |
| Sex of study population | character | population_sex | sex of survey population - see dropdown list |
| Population sample setting | character | population_sample_type | general setting of the survey - see dropdown list |
| Population group | character | population_group | specific group of the survey population - see dropdown list |

Continued on next page

Continued from previous page

| Data field | Expected type | Variable name | Notes |
|---|---|---|---|
| genome site | character | genome_site | site of genome or gene studied |
| Urban/Rural area | character | urban_rural | urban or rural setting of ZIKV circulation - see dropdown list |
| Data availability | character | data_availability | Statement about availability of the data presented in the paper - see dropdown list |
| Gene | logical | Gene | Name of the genomic sequence or gene under analysis |
| Genomic sequence available? | logical | genomic_sequence_available | tick box whether genomic sequence data are available (true) or not (false) |
| Reproduction number pathway | character | R_pathway | transmission pathway that reproduction number is based on for vector-borne diseases |
| Method to estimate R | character | method_R | method used for estimation of the reproduction number - see dropdown list |
| Other delay start point | character | other_delay_start | start point for delays not in the parameter type dropdown list (e.g., delay from ... to ...) |
| Other delay end point | character | other_delay_end | end point for delays not in the parameter type dropdown list (e.g., delay from ... to ...) |
| Numerator | integer | cfr_ifr_numerator | numerator of either cfr/ifr (deaths) or seroprevalence (number seropositive) estimates |
| Denominator | integer | cfr_ifr_denominator | denominator of either cfr/ifr (cases) or seroprevalence (number tested) estimates |
| Is the cfr/ifr estimate adjusted? | character | cfr_ifr_method | is the cfr/ifr estimate adjusted, unadjusted, or unspecified - see dropdown list |
| Is the seroprevalence estimate adjusted? | character | serop_method | is the seroprevalence estimate adjusted (accounting for assay's sensitivity and specificity) - see dropdown list |
| Outcome for risk factor(s) | character | riskfactor_outcome | outcome for risk factor(s) - see dropdown list |
| Risk factor name | character | riskfactor_name | risk factor name - see dropdown list |
| Risk factor occupation | character | riskfactor_occupation | if risk factor is an occupation, then specified occupation as risk factor - see dropdown list |
| Risk factor adjusted | character | riskfactor_adjusted | adjustment status of risk factor(s) - see dropdown list |
| Risk factor significant | character | riskfactor_significant | statistical significance of risk factor(s) - see dropdown list |
| Location type | character | parameter_context_location_type | If a location was captured, what it was, e.g. district or region level. |
| Human delay from | character | parameter_hd_from | starting point for human delay captured |
| Human delay to | character | parameter_hd_to | end point for human delay captured |
| Variability parameter value | numeric | parameter_2_value | variability value |
| Variability exponent | integer | exponent_2 | variability value exponent (base 10) |
| Variability lower bound | numeric | parameter_2_lower_bound | lower bound of the parameter (depending on variability type) |
| Variability upper bound | numeric | parameter_2_upper_bound | upper bound of the parameter (depending on variability type) |
| Variability unit | character | parameter 2 unit | units for variability estimate |
| Variability from supplement | logical | Method$_{2}from_{s}upplement$ | If variabilility parameter was captured from the supplement |
| Variability statistical approach | character | parameter_2_statistical_approach | if observed sample statistic, estimated parameter or unspecified. |
| Variability single uncertainty value | integer | parameter_2_uncertainty_single_value | Value of single type uncertainty of variability. |

Continued on next page

Continued from previous page

| Data field | Expected type | Variable name | Notes |
|---|---|---|---|
| Variability single uncertainty type | character | parameter_2_uncertainty_type | Standard Error or single value uncertainty value of posterior distribution |
| Variability uncertainty lower value | numeric | parameter_2_uncertainty_lower_value | value of lower bound of variability uncertainty |
| Variability uncertainty lower value | numeric | parameter_2_uncertainty_upper_value | value of upper bound of variability uncertainty |
| Variability uncertainty type | character | parameter_2_value_type | Type of variability uncertainty (e.g. 95% Credible Interval) |
| Variability uncertainty paired lower value | numeric | parameter_2_sample_paired_lower | value of paired lower bound of variability uncertainty |
| Variability uncertainty paired upper value | numeric | parameter_2_sample_paired_upper | value of paired upper bound of variability uncertainty |
| Variability distribution type | character | distribution_2_type | Distribution of variability |
| First parameter type of variability distribution | character | distribution_2_par1_type | Shape, scale |
| First parameter value of variability distribution | numeric | distribution_2_par1_value | value of first parameter of variability distribution |
| Second parameter type of variability distribution | character | distribution_2_par2_type | Shape, scale |
| Second parameter type of variability distribution | numeric | distribution_2_par2_value | value of second parameter of variability distribution |
| First parameter of variability distribution has uncertainty associated with it | logical | distribution_2_par2_uncertainty | Yes/no option, not captured in form |
| Parameter distribution | character | distribution_par1_uncertainty | distribution of main parameter captured |
| Variability single type uncertainty | character | parameter_2_uncertainty_single_type | Single type uncertainty of variability |
| Second parameter of variability distribution has uncertainty associated with it | logical | distribution_2_par1_uncertainty | Yes/no option, not captured in form |

Table A.4: Parameters form fields: refer to epireview package documentation for dropdown options.

##### A.2.4 Quality Assessment

For each study, across all outbreaks, transmission models and parameters extracted, we provided an overall quality assessment (QA) score using a custom questionnaire, given in Table A.5. The overall QA score is calculated as the mean number of "Yes" responses, excluding any non-applicable questions. The distribution of QA scores per question and the trend of QA scores over time are shown in Figure B.2.

| **Theme** | **Question** |
|---|---|
| Is the methodological/statistical approach suitable? (how the data are used) | 1. Clear and reproducible<br>2. Robust and appropriate for the aim [subjective criteria] |
| Are the assumptions appropriate? (input parameters/assumptions - what goes into the methodology) | 3. Clear and reproducible<br>4. Justified (published study or analysis of data)[objective criteria] |
| Are the data appropriate for the selected methodological approach? | 5. Clearly described and reproducible<br>6. Are issues in the data clearly discussed and acknowledged?<br>7. Are issues in the data accounted for in the chosen methodological approach? |

Table A.5: Quality assessment questionnaire: possible responses for each question listed were: Yes, No, and NA (non-applicable).

#### A.3 Data Analysis

In this section, we describe the procedures for deduplicating outbreak data and parameter estimates, the subsequent meta-analyses conducted using the deduplicated data, and the supplementary Bangladesh IEDCR surveillance data used.

##### A.3.1 Data deduplication

Many studies included in our review reported data that may partially duplicate study samples, for example through overlapping study periods or population groups that are subsets of one another. Such duplication is problematic for representing unique outbreaks and conducting meta-analyses, as it may lead to overrepresentation of certain estimates and bias pooled estimates by violating the assumption of independent observations. Below we describe the processes we used for deduplication.

**Parameters**
In preparation for meta-analysis, we deduplicated extracted CFR estimates and incubation periods.

We classified duplicates into either assumed or known. Extracted parameters were classified as known duplicates if they referred to the same underlying data (e.g. a subset of reported cases) and as assumed duplicates if they were based on overlapping contexts (e.g. location, population group, or time period). To maximise the number of non-overlapping estimates we adopted the following logic:

- We constructed date ranges for each estimate using the reported study start and end dates and sorted the data accordingly.
- We prioritised estimates covering distinct study periods, population groups, and locations.
- Mutually exclusive disaggregated groups were preferred to aggregated groups.
- Where overlap was unavoidable, the estimate with the larger sample size was selected.

Only two known duplicate incubation period estimates were identified among those suitable for meta-analysis (see Section A.3.2 for the suitability criteria), both related to the study by Nikolay et al. [8]. The first was a subsample of a larger analysis, and the second was an estimated model parameter. In contrast, many more CFR estimates required deduplication. Figure B.11 illustrates the study periods and population groups of all extracted CFR estimates and indicates which were retained or classified as assumed or known duplicates.

**Outbreaks**
Extracted outbreaks were deduplicated to ensure each outbreak was represented once in the dataset, using the following logic:

- Outbreak locations were mapped division/state level to harmonise spelling across extractions.
- The outbreak data was sorted by outbreak date (range) and location.
- For outbreaks which cover the same date range and location, the outbreak which covers a longer time interval was used.
- Studies which reported on outbreaks with a duration over 5 years were excluded as they cover multiple underlying outbreaks.
- If individual country level outbreak data existed those were preferred to outbreak reports which pooled multiple countries.
- If articles reported on *clusters* of outbreaks, we only considered the underlying outbreaks.

Following the deduplication of CFR estimates and outbreaks, we cross-referenced the unique outbreaks identified in both datasets and noted differences for India and Bangladesh. Consequently, we estimated naïve CFRs ($\frac{\text{Total reported deaths}}{\text{Total reported cases}}$) from the outbreak data and synthesised these using the same meta-analysis methodology applied to the extracted CFR estimates for comparison.

##### A.3.2 Meta-analysis

The meta-analysis of case fatality ratio and the incubation period followed a standard approach and was conducted using the `meta` R package [9].

A mixed-effects model is a linear model

$$y_i = \beta_0 + \sum_{j=1} \beta_j x_{ij} + u_i + \epsilon_i$$

where $y_i$ are the observed data, $x_{ij}$ are explanatory variables, $\beta_j$ are fixed effect coefficients, $u_i$ are the random effects (centred around zero and independent across $i$), and $\epsilon_i$ are error terms. A standard linear regression model is a special case of the above linear mixed-effects model without random effects.

Our meta-analysis was conducted using linear regression models with only an intercept term $\beta_0$ and mixed-effects models that additionally included a random-effects term $u_i$. Our meta-analysis was conducted using both linear regression models with only an intercept term $\beta_0$ and mixed-effects models that additionally included a random-effects term $u_i$. The random-effects term captures between-study-estimate heterogeneity not attributable to sampling error alone; Note that we assume estimates reported separately within a study and retained after deduplication represent distinct estimates and treat them as individual studies. In meta-analysis terminology, these correspond to common-effect and random-effects models, respectively. In each case, the intercept $\beta_0$ represents the pooled estimate.

*Data inclusion protocol for meta-analysis*

- The data is taken as extracted (post-data cleaning).
- Any filtering (for subgroup, setting, country) will be stated clearly in the analysis.
- Parameter estimates, which are ranges as the 'rule of 3' was applied, were not included in the meta-analysis.

**Case fatality ratio**

To conduct the CFR meta-analysis, we fitted a binomial generalised linear mixed-effects model with a logit link to estimate the pooled random-effect. Specifically, we modelled

$$\begin{aligned} Y_i &\sim \text{Binomial}(n_i, p_i), \\ \text{logit}(p_i) &= \beta_0 + u_i, \\ u_i &\sim N(0, \tau^2). \end{aligned}$$

where $i$ represents an individual estimate, $Y_i = \text{Deaths}_i$, $n_i = \text{cases}_i$, and $p_i = \text{CFR}_i$, and $\tau^2$ is the between-study-estimate heterogeneity. The corresponding common-effect model was a binomial generalised linear model with a logit link and no random-effects term. We applied an inverse logit to the estimated intercept of each model to report the respective pooled effect:

$$\widehat{\text{pooled CFR}} = \frac{e^{\hat{\beta}_0}}{1 + e^{\hat{\beta}_0}}$$

The meta-analyses conducted report the common and random effects for each subgroup to investigate any patterns between groups and any patterns across datasets. Subgroup results are estimated from separate models, whereas the overall pooled results are estimated from models fitted to all studies. We note that:

- We applied this method to three subgroups of extracted CFR parameters: country of population, population group, and time period shown in (main text) Figure 3, Figure B.9), and Figure xxx respectively.
- As a sensitivity analysis assessing the impact of deduplication, we repeated the meta-analyses of extracted CFR parameters excluding only known duplicates across all subgroups (Population country: Figure B.12, Population group under study: Figure B.13, Over time: Figure B.14). As expected given overlapping estimates, the overall common-effect and random-effects estimates differ between the deduplicated and non-deduplicated meta-analyses. However, these differences are small and do not change the interpretation.

- We applied this method to naïve CFRs estimated from deduplicated outbreak data (Figure B.6)

In stratified analyses, some groups may contain only a single estimate; in such cases, the naïve CFR is reported. Because IEDCR surveillance data originate from a single source, accounting for between-estimate heterogeneity was unnecessary, and we instead report naïve CFRs with binomial confidence intervals.

**Incubation period**
To estimate the pooled random-effect, we modelled the reported sample means for the incubation period because individual-level data were unavailable. Specifically, we modelled

$$\bar{Y}_i = \beta_0 + u_i + e_i,$$
$$e_i \sim \mathcal{N}(0, v_i^2),$$
$$u_i \sim \mathcal{N}(0, \tau^2),$$

where $i$ represents an individual estimate, $\bar{Y}_i$ is the mean incubation period estimate, $e_i$ represents sampling error, and $v_i^2 = \sigma_i^2/n_i$ is the within-study-estimate sampling variance ($\sigma_i$ =standard deviation and $n_i$ =cases), and $\tau^2$ is the between-study-estimate heterogeneity. The corresponding common-effect model was a simple linear regression model with no random-effects term. The meta-analysis methods used are described in comprehensive detail by Harrer et al. [10].

To apply the mode described above, the analysis required paired mean and standard deviation estimates of the incubation period. However, the underlying estimates extracted reported several different combinations of central estimates and measures of population variability (e.g. mean (range), mean (standard deviation), median (interquartile range), median (range) etc.). To address this, we use the functionality of `metamean`, which is implemented in the `meta` package, to infer the means and standard deviations where other combinations were reported. In particular, we used the method of Cai, Zhou, and Pan [11], which accommodates unknown non-normal distributions. However, applied to our extracted estimates, this approach was only suitable for median (interquartile range) and median (range) estimates. This limited our ability to conduct a meta-analysis of symptom-onset-to-death, for which many estimates were available (n=18), but seven were reported as a mean and range.

A single meta-analysis was conducted and is presented in (main text) Figure 4; subgroup analyses were not performed owing to the small sample size.

##### A.3.3 Bangladesh IEDCR data

In addition to extracted CFR parameters and extracted outbreaks, we supplemented our analysis with NiV surveillance data from Bangladesh's Institute of Epidemiology, Disease Control and Research (IEDCR), which provide more complete data on cases and deaths from 2001 to 2025.

This data come from a national, hospital-based sentinel and outbreak surveillance system for NiV that has been running since 2006, with additional retrospective data back to 2001 [12]. The system currently operates in 14 government hospitals across all 8 divisions of the country and captures suspected NiV cases, recording clinical details, exposure history (especially consumption of raw date palm sap), epidemiologic links, and laboratory test results (RT-PCR and ELISA), along with supplementary outbreak investigation information such as contact tracing, transmission chains, and patient outcomes. In essence, these IEDCR data constitute a long-term, nationwide surveillance and outbreak dataset that describes when and where NiV occurs in Bangladesh, how cases are exposed and linked, and how the disease behaves clinically in the population.

We accessed publicly available, aggregate-level data downloaded from the Nipah Situation Dashboard [13]. These data were used to generate the map shown in Figure B.6 and the plot of naïve CFR estimates over time shown in Figure B.8.

#### A.4 Reporting estimate uncertainty and population variability

When a parameter is reported, it is often presented as a central estimate together with either *estimate uncertainty* or *population variability*. These concepts reflect different sources of variation and should not be conflated.

Estimate uncertainty reflects uncertainty around the estimate itself and arises from multiple sources; examples, although not exhaustive, include data limitations such as small sample size and measurement error, as well as choices made during estimation, such as model specification. Among the parameters extracted, uncertainty was most commonly indicated using 95% confidence intervals or 95% credible intervals, although we also captured single-measure uncertainty such as standard errors.

Population variability reflects heterogeneity within the underlying population from which the central estimate is generated. Capturing true population variability is typically not possible and, in practice, it is commonly approximated using either sample-based measures of variability or estimated more directly as a parameter of a fitted distribution. In the latter case, the estimated population variability parameter may itself be subject to estimate uncertainty. Among the parameters extracted, sample-based variability measures such as interquartile ranges and full ranges were most common. Nikolay et al. [8] was the only study in our review to estimate population variability directly, by estimating the standard deviation of a fitted gamma distribution, for both the serial interval and the incubation period; uncertainty around the variability parameter was reported only for the incubation period.

While most studies reported a central estimate for parameters of interest, some instead reported a range of values or we applied the 'rule of 3' to extract a range as outlined in Section A.2.3. These ranges should not be interpreted as measures of population variability. Rather, they represent ranges of central values and are included when we refer to "central estimates".

Throughout the forest plots used in our manuscript, we distinguish between these quantities as indicated below. Note that in these plots we indicate only two-sided estimate uncertainty and population variability. Two studies reported standard deviations for variability, which are consequently not shown [14, 8].

Estimate uncertainty is displayed using a solid black error bar

Population variability is displayed using a dashed black error bar

A range of central values is displayed using thick coloured bands

### B Additional Results

#### B.1 Quality Assessment Results

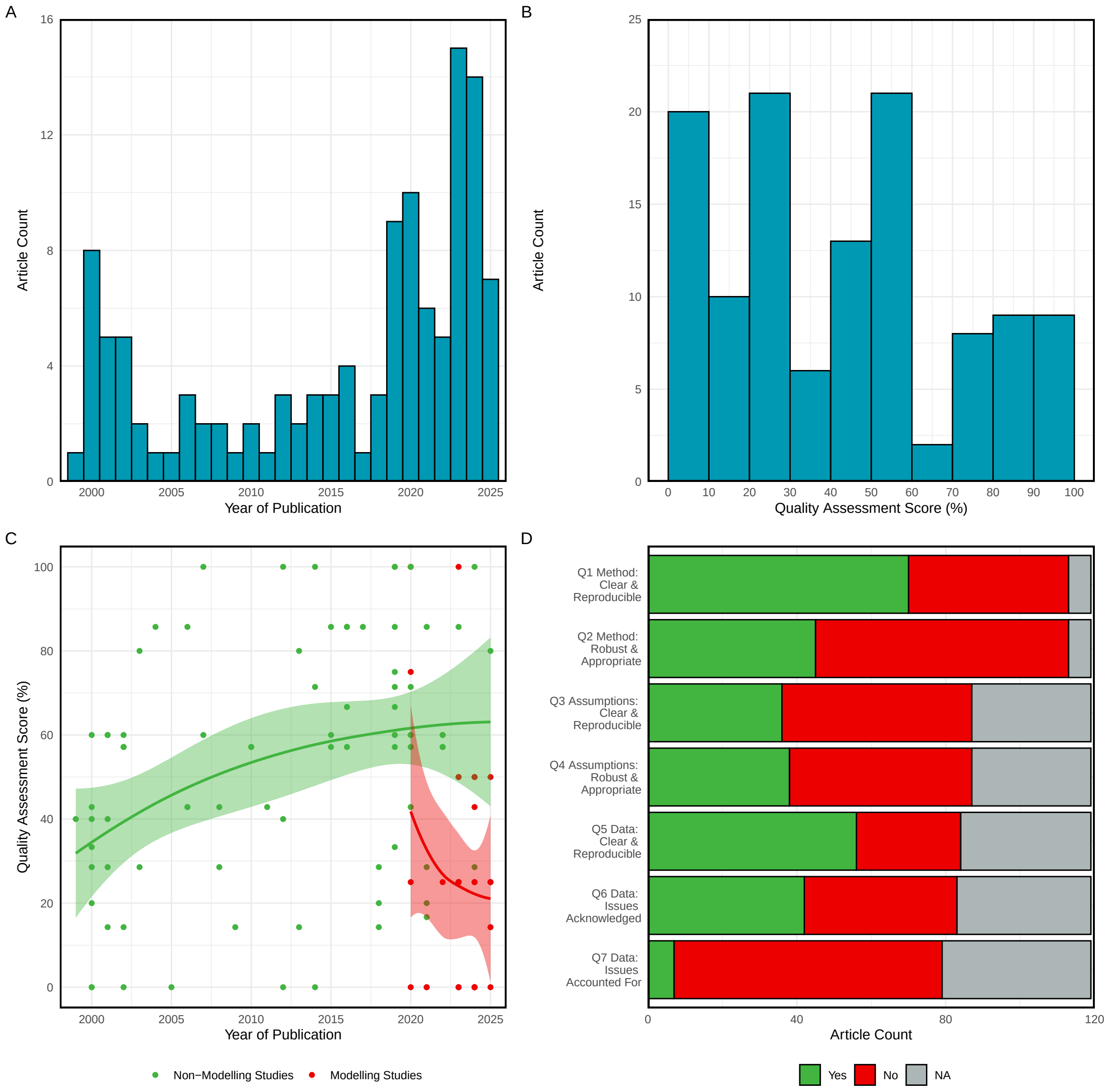

Figure B.2: (A) Article count by year of publication; (B) article count by quality assessment score (defined as percentage of 'Yes' answers relative to sum of 'Yes' and 'No' answers for each paper, removing 'NAs'); (C) quality assessment score by year of publication (time trends for articles with and without transmission models fitted via local polynomial regression); (D) article count for each quality assessment question scoring 'Yes', 'No' or 'Non-Applicable'.

#### B.2 Parameter and Model Overview

##### B.2.1 Parameters

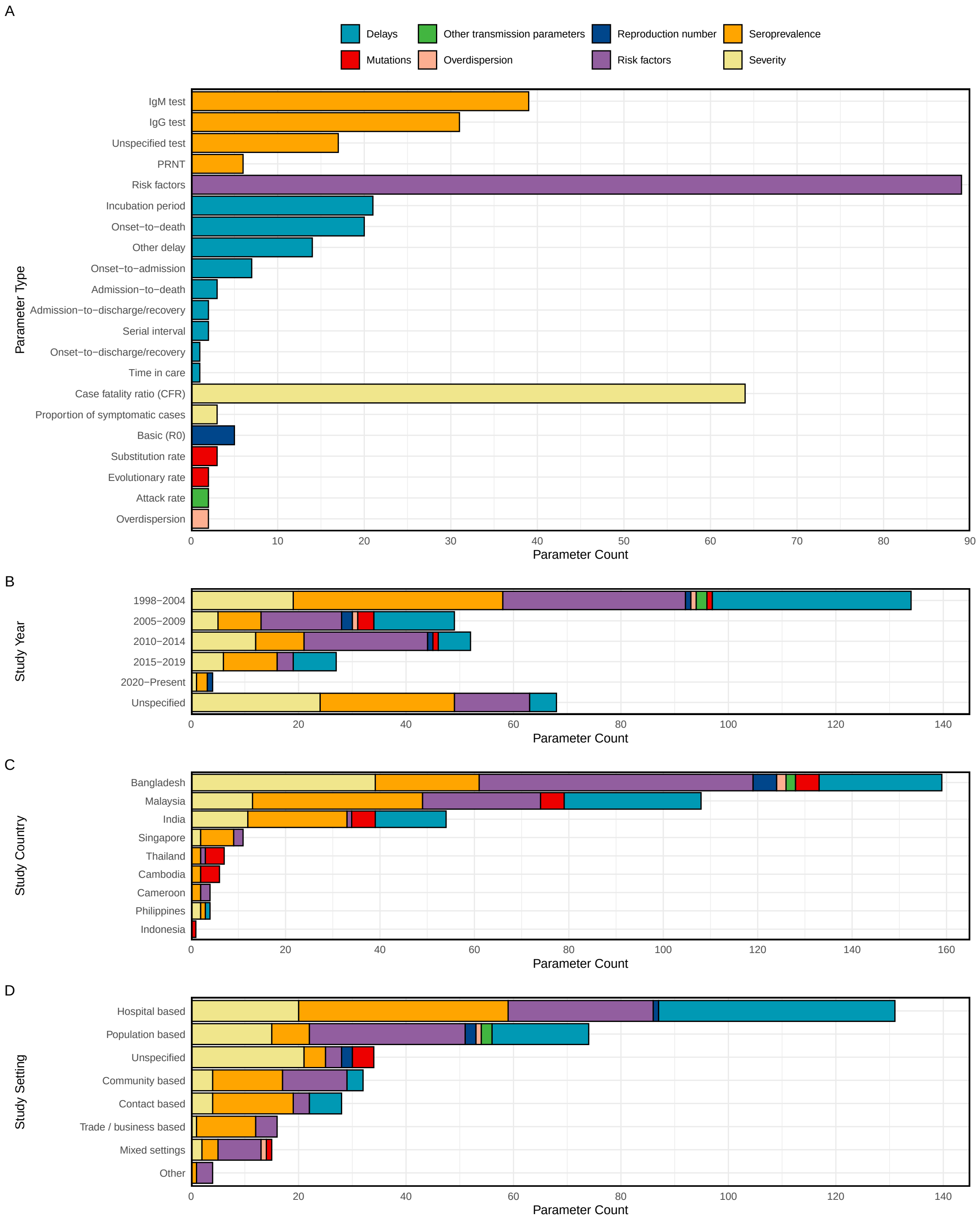

Figure B.3: Parameter count by (A) parameter type, (B) study setting (C) survey period (D) study country. Colour corresponds to the parameter class. Note that in (D), the same parameter may be attributed to multiple countries. Consequently, the total number of models displayed (352) exceeds the number of unique parameters (332).

| Parameter class | Total (no QA) | | QA > 0.5 | |
|---|---|---|---|---|
| | No. parameters | No. papers | No. parameters | No. papers |
| Attack rate | 2 | 1 | 2 | 1 |
| Human delay | 70 | 30 | 34 | 16 |
| Mutations | 5 | 5 | 4 | 4 |
| Overdispersion | 2 | 2 | 2 | 2 |
| Reproduction number | 5 | 5 | 3 | 3 |
| Risk factors | 89 | 38 | 58 | 23 |
| Seroprevalence | 93 | 43 | 50 | 24 |
| Severity | 67 | 38 | 29 | 22 |

Table B.6: Total parameters extracted and total number of papers these parameters were extracted from by parameter class. Middle column includes all parameters. Right column only includes those papers that scored higher than 0.5 for quality assessment (see Table A.5). QA is used as an acronym for quality assessment.

| Parameter class | Total (no QA) | | QA > 0.5 | |
|---|---|---|---|---|
| | No. parameters | No. papers | No. parameters | No. papers |
| Attack rate | 2 | 1 | 2 | 1 |
| Delay - admission>death | 3 | 3 | 2 | 2 |
| Delay - admission>discharge/recovery | 2 | 1 | 2 | 1 |
| Delay - incubation period | 21 | 16 | 11 | 7 |
| Delay - serial interval | 1 | 1 | 1 | 1 |
| Delay - onset>admission | 7 | 7 | 3 | 3 |
| Delay - onset>death | 20 | 18 | 13 | 11 |
| Delay - onset>discharge/recovery | 1 | 1 | | |
| Delay - time in care (length of stay) | 1 | 1 | | |
| Delay - other | 14 | 8 | 2 | 1 |
| Mutations - evolutionary rate | 2 | 2 | 1 | 1 |
| Mutations - substitution rate | 3 | 3 | 3 | 3 |
| Overdispersion | 2 | 2 | 2 | 2 |
| Reproduction number (Basic R0) | 5 | 5 | 3 | 3 |
| Risk factors | 89 | 38 | 58 | 23 |
| Seroprevalence - IgG | 31 | 22 | 17 | 12 |
| Seroprevalence - IgM | 39 | 28 | 21 | 15 |
| Seroprevalence - PRNT | 6 | 5 | 6 | 5 |
| Seroprevalence - Unspecified | 17 | 10 | 6 | 4 |
| Severity - case fatality rate (CFR) | 64 | 36 | 26 | 20 |
| Severity - proportion of symptomatic cases | 3 | 3 | 3 | 3 |

Table B.7: Total parameters extracted and the total number of papers these parameters were extracted from by parameter type. Middle column includes all parameters. Right column only includes those papers that scored higher than 0.5 for quality assessment (see Table A.5). QA is used as an acronym for quality assessment.

##### B.2.2 Models

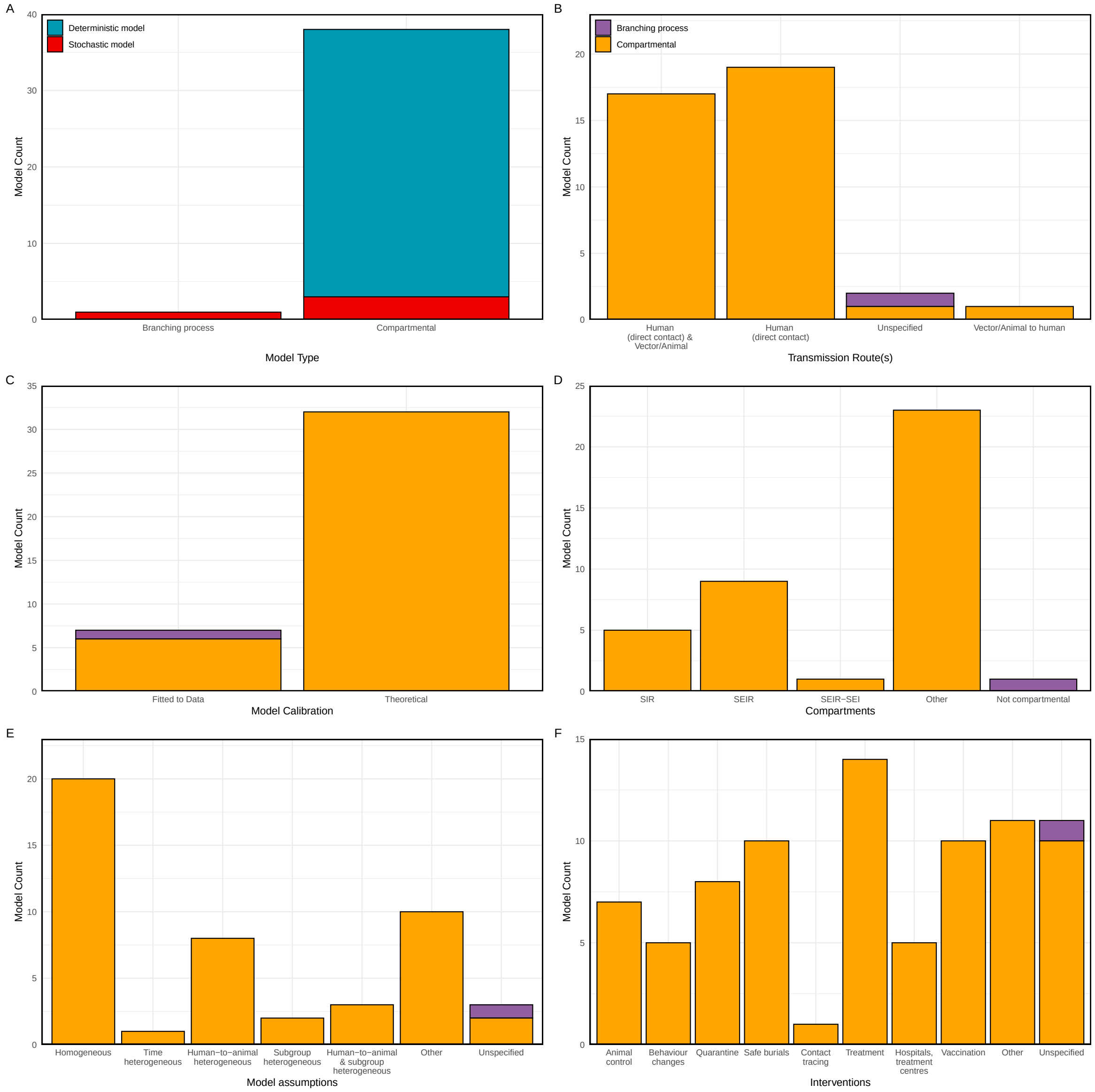

Figure B.4: Total published Nipah transmission model count by (A) model type, (B) transmission routes, (C) model output calibration, (D) compartmental model form (where applicable, otherwise listed as not compartmental), (E) model assumptions, all columns excluding other and unspecified relate to transmission heterogeneity, (F) interventions incorporated. Colour corresponds to the deterministic or stochastic nature of the model in (A) and the model type in (B-F). Note that in the final panel row, individual assumptions or interventions within the same model are shown separately. Consequently, the total number of models displayed (48 in panel E and 83 in panel F) exceeds the number of unique models (40).

#### B.3 Extracted Data

This section provides all extracted outbreak, model, and parameter data.

##### B.3.1 Outbreaks

Table B.9 lists all extracted outbreaks, in alphabetical order of outbreak country:

1. Bangladesh
2. India
3. Malaysia
4. Malaysia, Singapore
5. Singapore

| Location | Outbreak Dates | Outbreak Source | Confirmation Method | Suspected Cases | Probable Cases | Confirmed Cases | Unspecified Cases | Deaths | Sex Disaggre-gation | Prop male cases | Source |
|---|---|---|---|---|---|---|---|---|---|---|---|
| **Bangladesh** | | | | | | | | | | | |
| Barishal Division | 2020 | | Confirmed & Suspected | | | 1 | | | Unspecified | | Satter (2023) |
| | Dec 2013 - Apr 2014 | Date palm sap | Confirmed & Suspected | 3 | 1 | | | 2 | | | Hassan (2018) |
| | Dec 2013 - Apr 2014 | Date palm sap | Confirmed & Suspected | 3 | 1 | | | 4 | | | Hassan (2018) |
| | Dec 2013 - Apr 2014* | Date palm sap | Confirmed & Suspected | 1 | 1 | | | 2 | | | Hassan (2018) |
| | Dec 2013 - Apr 2014* | Date palm sap | Confirmed & Suspected | 1 | 1 | | | 2 | | | Hassan (2018) |
| Gaibandha, Natore, Rajshahi, Naogaon, Rajbari, Pabna, Jhenaidah, Mymensingh | Jan 2013 - Feb 2013 | | | | | | 12 | 10 | | | Kulkarni (2013) |
| Mymensingh Division | 2013 | | Confirmed & Suspected | | | 2 | | | | | Satter (2023) |
| Joypurhat, Rajshahi, Natore, Rajbari, Gopalganj | Feb 2012 - Unspecified | | | | | | 12 | 10 | | | Kulkarni (2013) |
| Lalmohirhat, Dinajpur, Comilla, Nilphamari, Rangpur | Jan 2011 - Feb 2011 | | | | | | 44 | 40 | | | Kulkarni (2013) |
| Chattogram Division | 2011 | Other | Confirmed & Suspected | | | 1 | | | Unspecified | | Satter (2023) |
| Lalmonirhat, Dinajpur, Rajbari, Rangpur | 23 Dec 2010 - 1 Mar 2011 | Date palm sap, Other | Confirmed & Suspected | 23 | | 20 | | 38 | Other | 0.7 | Chakraborty (2016) |
| Faridpur, Rajbari, Gopalganj, Madaripur | Feb 2010 - Mar 2010 | | | | | | 16 | 14 | | | Kulkarni (2013) |

continued on next page

continued from previous page

| Location | Outbreak Dates | Outbreak Source | Confirmation Method | Suspected Cases | Probable Cases | Confirmed Cases | Unspecified Cases | Deaths | Sex Disaggre-gation | Prop male cases | Source |
|---|---|---|---|---|---|---|---|---|---|---|---|
| Faridpur | 7 Jan 2010 - 28 Jan 2010 | Date palm sap, Other | Confirmed & Suspected | 5 | | 11 | | 14 | Unspecified | 0.56 | Sazzad (2013) |
| Gaibandha, Rangpur, Nilphamari | Jan 2009 - Unspecified | | | | | | 3 | 0 | | | Kulkarni (2013) |
| Rajbari | Jan 2009 - Unspecified | | | | | | 1 | 1 | | | Kulkarni (2013) |
| Rajbari, Faridpur | Apr 2008 - Unspecified | | | | | | 7 | 7 | | | Kulkarni (2013) |
| Manikgonj | 6 Feb 2008 - 10 Mar 2008 | Date palm sap | Molecular (PCR etc) | | | | 4 | | | | Rahman (2012) |
| Rajbari | 6 Feb 2008 - 10 Mar 2008 | Date palm sap | Molecular (PCR etc) | | | | 6 | | | | Rahman (2012) |
| Manikgonj | Feb 2008 - Unspecified | | | | | | 4 | 4 | | | Kulkarni (2013) |
| Naogaon | Apr 2007 - Unspecified | | | | | | 3 | 1 | | | Kulkarni (2013) |
| Sadar Upazila | Mar 2007 - Apr 2007 | Unknown | Confirmed & Suspected | 3 | | | 11 | 5 | | | Homaira (2010b) |
| Kushtia, Pabna, Natore | Mar 2007 - Unspecified | | | | | | 8 | 5 | | | Kulkarni (2013) |
| Haripur Upazila (Subdistrict) Of Thakurgaon District | 15 Jan 2007 - 28 Feb 2007 | Unknown | Confirmed & Suspected | 5 | 2 | | 5 | | | | Homaira (2010a) |
| Thakurgaon | Jan 2007 - Feb 2007 | | | | | | 7 | 3 | | | Kulkarni (2013) |
| Tangail | Jan 2005 - Mar 2005 | | | | | | 12 | 11 | | | Kulkarni (2013) |
| Tangail | 15 Dec 2004 - 31 Jan 2005 | Date palm sap | Symptoms | 12 | 124 | | | 11 | Suspected | 0.58 | Luby (2006) |
| Faridpur | Apr 2004 - Unspecified | | | | | | 36 | 27 | | | Kulkarni (2013) |
| Faridpur | 19 Feb 2004 - 16 Apr 2004 | Unknown | Symptoms | 23 | 13 | | | 27 | | 0.59 | Gurley (2007a) |
| Faridpur | Feb 2004 - Apr 2004 | Domestic animal, Date palm sap | Confirmed & Suspected | | 36 | | | 27 | | | Hossain (2008) |
| Dhaka Division | 2004 - 2021 | | Confirmed & Suspected | | | 147 | | | | | Satter (2023) |
| Rangpur Division | 2004 - 2015 | Unknown | Confirmed & Suspected | | | 82 | | | | | Satter (2023) |
| Rajbari, 7 Other Northwestern Districts | Jan 2004 - Apr 2004 | Domestic animal, Date palm sap | Confirmed & Suspected | | 31 | | | 23 | | | Hossain (2008) |
| Rajbari | Jan 2004 - Feb 2004 | | | | | | | | | | Gurley (2007b) |

continued on next page

continued from previous page

| Location | Outbreak Dates | Outbreak Source | Confirmation Method | Suspected Cases | Probable Cases | Confirmed Cases | Unspecified Cases | Deaths | Sex Disaggre-gation | Prop male cases | Source |
|---|---|---|---|---|---|---|---|---|---|---|---|
| Rajbari, Faridpur | 2004 | | Confirmed & Suspected | | | | 48 | | | | Harcourt (2005) |
| Unspecified 7 Districts | 2004 | | Confirmed & Suspected | | | | 19 | | | | Harcourt (2005) |
| Rajbari | Jan 2004 - Unspecified | | | | | | 31 | 23 | | | Kulkarni (2013) |
| Naogaon | 11 Jan 2003 - 28 Jan 2003 | Unknown | Symptoms | 4 | 8 | | | 8 | Other | 0.67 | Hsu (2004) |
| Rajshahi Division | 2003 - 2021 | Unknown | Confirmed & Suspected | | | 53 | | | | | Satter (2023) |
| Naogaon | Jan 2003 | Domestic animal, Date palm sap | Confirmed & Suspected | | 12 | | | 8 | | | Hossain (2008) |
| Naogaon | Jan 2003 - Unspecified | | | | | | 12 | 8 | | | Kulkarni (2013) |
| Meherpur | 20 Apr 2001 - 20 May 2001 | Unknown | Symptoms | 4 | 9 | | | 9 | Other | 0.46 | Hsu (2004) |
| Meherpur | Apr 2001 - May 2001 | | | | | | 13 | 9 | | | Kulkarni (2013) |
| Meherpur | Apr 2001 - May 2001 | Domestic animal, Date palm sap | Confirmed & Suspected | | 13 | | | 9 | | | Hossain (2008) |
| Khulna Division | 2001 - 2015 | | Confirmed & Suspected | | | 36 | | | | | Satter (2023) |
| **India** | | | | | | | | | | | |
| Kozhikode | 25 Aug 2023 - 29 Sep 2023 | Unknown | Molecular (PCR etc) | 6 | | | | 2 | Confirmed | 1 | As (2024) |
| Kozhikode | 4 Sep 2021 - Unspecified | Unknown | Confirmed & Suspected | 1 | | | | 1 | | | Yadav (2022) |
| Kozhikode | 2 May 2018 - 31 May 2018 | Wild animal | Confirmed & Suspected | 18 | | 5 | | 21 | Confirmed | 0.65 | Pallivalappil (2020) |
| Kerala | 2 May 2018 - 29 May 2018 | Unknown | Symptoms | 18 | | 4 | 1 | 21 | Unspecified | 0.65 | Arunkumar (2019) |
| Kozhikode, Kerala | May 2018 | Wild animal, Other | | 18 | 147 | | | 16 | | 0.61 | Thomas (2019) |
| Kerala | May 2018 - Unspecified | Unknown | Molecular (PCR etc) | 18 | | 4 | | 20 | Unspecified | | Thulaseedaran (2018) |
| West Bengal | 9 Apr 2007 - 28 Apr 2007 | Date palm sap | Confirmed & Suspected | 3 | | 2 | | 5 | Unspecified | | Arankalle (2011) |
| Nadia | Apr 2007 - Unspecified | | | | | | 5 | 5 | | | Kulkarni (2013) |
| Siliguri, West Bengal | 31 Jan 2001 - 23 Feb 2001 | Unknown | Symptoms | | 66 | | | 45 | Suspected | | Harit (2006) |

continued on next page

continued from previous page

| Location | Outbreak Dates | Outbreak Source | Confirmation Method | Suspected Cases | Probable Cases | Confirmed Cases | Unspecified Cases | Deaths | Sex Disaggre-gation | Prop male cases | Source |
|---|---|---|---|---|---|---|---|---|---|---|---|
| Siliguri, West Bangal | 31 Jan 2001 - 23 Feb 2001 | Unknown | Confirmed & Suspected | 10 | 56 | | | | | | Chadha (2006) |
| Siliguri | Jan 2001 - Feb 2001 | | | | | | 66 | 45 | | | Kulkarni (2013) |
| **Malaysia** | | | | | | | | | | | |
| Negri Sembilan State | Feb 1999 - Jun 1999 | Domestic animal | Symptoms | | 94 | | | 30 | | | Goh (2000) |
| | 29 Sep 1998 - Dec 1999 | Domestic animal, Unknown | Unspecified | | | | 283 | 109 | Unspecified | 0.82 | Chua (2003) |
| Negeri Sembilan | 29 Sep 1998 - Dec 1999 | Domestic animal, Unknown | Unspecified | | | | 231 | 86 | Unspecified | | Chua (2003) |
| Selangor | 29 Sep 1998 - Dec 1999 | Domestic animal, Unknown | Unspecified | | | | 24 | 8 | Unspecified | | Chua (2003) |
| | Sep 1998 - Unspecified | Domestic animal | Symptoms | | | | 265 | 105 | | | Chua (2000b) |
| Perak | 29 Aug 1998 - Dec 1999 | Domestic animal, Unknown | Unspecified | | | | 28 | 15 | Unspecified | | Chua (2003) |
| **Malaysia, Singapore** | | | | | | | | | | | |
| | Sep 1998 - Apr 1999 | Domestic animal | Symptoms | | | | 246 | | | | Pulliam (2012) |
| **Philippines** | | | | | | | | | | | |
| Senator Ninoy Aquino | 3 Mar 2014 - 24 May 2014 | Domestic animal | Confirmed & Suspected | 17 | | | | 9 | | | Ching (2015) |
| **Singapore** | | | | | | | | | | | |
| | 16 Mar 1999 | Domestic animal | Symptoms | 11 | 24 | | | 1 | Confirmed | | Paton (1999) |
| | 13 Mar 1999 - Unspecified+ | Domestic animal | Molecular (PCR etc) | 22 | | | | | Confirmed | 1 | Chan (2002) |
| | Mar 1999 - Unspecified | Domestic animal | Symptoms | | | | 11 | 1 | | | Chua (2000b) |

Table B.8: Characteristics of extracted ‘outbreaks’ reported by study authors grouped by country. *indicates that the number of asymptomatic cases is available and +indicates that asymptomatic transmission was observed. Within each country (sorted alphabetically), outbreaks are sorted in descending order of starting date and of ending date.

##### B.3.2 Models

Table B.9 lists all extracted transmission models, in order of:

1. Branching process models
2. Compartmental models
   (a) Deterministic
   (b) Stochastic
3. Other deterministic models

| Transmission Route(s) | Human Transmission Heterogeneity | Compartments | Fitted | Interventions | Spatial | Spillover | Uncertainty | Data | Code (Language) | Source |
|---|---|---|---|---|---|---|---|---|---|---|
| **Branching process** | | | | | | | | | | |
| Human-human | | | Yes* | | No | No | Yes | Yes | Yes+ (R) | Bradbury (2023) |
| **Compartmental - deterministic** | | | | | | | | | | |
| Animal-human | Animal | Other | No | | No | Yes | No | Yes | No | Agarwal (2020) |
| Animal-human | Animal | SIR | No | | No | Yes | No | No | Yes (Matlab) | Royce (2020) |
| Human-human | | Other | Yes | Safe burials | No | No | No | Yes | No | Barua (2023b) |
| Human-human | | Other | Yes* | Vaccination, animal control, treatment, safe burials, | No | No | No | No | No | Khan (2024) |
| Human-human | | Other | No | Safe burials | No | No | No | Yes | No | Kubra (2024) |
| Human-human | | Other | No | Safe burials | No | No | No | Yes | No | Zewdie (2020) |
| Human-human | | Other | No | Quarantine, hospitals | No | No | No | Yes | No | Tyagi (2021) |
| Human-human | | Other | No | Safe burials | No | No | No | Yes | No | Evirgen (2023) |
| Human-human | | Other | No | | No | No | No | No | No | Shah (2023) |
| Human-human | | SEIR | No | Quarantine, treatment, behaviour changes, other | No | No | No | Yes | No | Raza (2024) |
| Human-human | | SEIR | No | Quarantine, treatment | No | No | No | Yes | No (Python) | Ramzan (2024) |
| Human-human | | SEIR | No | Treatment | No | No | No | Yes | No | Ali (2022) |
| Human-human | | SIR | No | Safe burials | No | No | No | Yes | No | Baleanu (2023) |
| Human-human | | SIR | No | | No | No | No | Yes | No | Shah (2024) |
| Human-human | | SIR | Yes* | | No | No | No | No | No | Sikder (2023) |

continued on next page

continued from previous page

| Transmission Route(s) | Human Transmission Heterogeneity | Compartments | Fitted | Interventions | Spatial | Spillover | Uncertainty | Data | Code (Language) | Source |
|---|---|---|---|---|---|---|---|---|---|---|
| Human-human | | SIR | No | | No | No | No | Yes | No | Sundar (2024) |
| Human-human, animal-human | | Other | No | Vaccination, behaviour changes | No | Yes | No | Yes | No (Python) | Ozioko (2023a) |
| Human-human, animal-human | | Other | No | Quarantine, animal control, other | No | Yes | No | Yes | No (Matlab) | Zewdie (2023) |
| Human-human, animal-human | | Other | No | Treatment, behaviour changes, other | No | Yes | No | No | No | Efat (2024) |
| Human-human, animal-human | | Other | No | Vaccination | No | Yes | No | Yes | No (Matlab) | Ahmed (2025) |
| Human-human, animal-human | | Other | No | Quarantine, treatment, other | No | Yes | No | Yes | No (Matlab) | Barman (2025) |
| Human-human, animal-human | | Other | No | Treatment, hospitals, behaviour changes, other | No | No | Yes | Yes | No | Dutta (2024) |
| Human-human, animal-human | | Other | Yes* | Vaccination, treatment | No | Yes | No | No | No (Matlab) | Yu (2025) |
| Human-human, animal-human | Animal | Other | Yes* | Animal control, other | No | Yes | Yes | Yes | No | Barua (2023a) |
| Human-human, animal-human | Animal | Other | No | Vaccination, treatment | No | Yes | No | Yes | No (Python) | Scott (2023) |
| Human-human, animal-human | Animal | Other | No | Animal control, other | No | Yes | Yes | Yes | No | Das (2020) |
| Human-human, animal-human | Animal | Other | No | Quarantine, animal control, treatment, safe burials | No | Yes | No | No | No | Samreen (2023) |
| Human-human, animal-human | Animal | Other | Yes* | Animal control, other | No | No | No | No | No | Li (2024) |
| Human-human, animal-human | Animal | Other | No | Treatment, hospitals, safe burials | No | Yes | No | Yes | No | Li (2023) |
| Human-human, animal-human | Groups, animal | Other | No | Vaccination, treatment | No | Yes | No | Yes | No (Other) | Abang (2024) |
| Human-human, animal-human | Groups, animal | SEIR | No | | Yes | Yes | No | Yes | No | Mabotsa (2025) |
| Human-human, animal-human | Time | SEIR | No | | No | Yes | No | Yes | No (Other) | Barua (2023c) |
| Human-human, animal-human | Groups, animal | SEIR-SEI | No | Vaccination, quarantine, animal control, treatment, safe burials, behaviour changes, other | No | Yes | No | Yes | No (Python) | Ozioko (2023b) |
| Human-human, animal-human, sexual | | Other | No | Vaccination, treatment, treatment centres, safe burials, other | No | Yes | No | Yes | No (Python) | Ozioko (2024) |

continued on next page

continued from previous page

| Transmission Route(s) | Human Transmission Heterogeneity | Compartments | Fitted | Interventions | Spatial | Spillover | Uncertainty | Data | Code (Language) | Source |
|---|---|---|---|---|---|---|---|---|---|---|
| Human-human, sexual | Groups | SEIR | No | Vaccination, other | No | No | No | Yes | No | Ozioko (2025) |
| **Compartmental - stochastic** | | | | | | | | | | |
| Human-human | | SEIR | No | | No | No | Yes | Yes | No | Raza (2021) |
| Human-human | | SEIR | No | Quarantine | No | No | No | Yes | No | Almoajel (2025) |
| Human-human | Groups | SEIR | No | Vaccination, contact tracing, hospitals | Yes | Yes | Yes | Yes | Yes+ (Python) | Carlson (2024) |

Table B.9: Characteristics of extracted transmission models grouped by model class. *indicates that the fitting method was specified and +indicates that a code README is available. Within each model class, models are sorted alphabetically by transmission route , by compartments where applicable (Other last), and by interventions.

##### B.3.3 Transmission parameters

Table B.10 lists all extracted transmission parameters, in order of:

1. Basic Reproduction Numbers ($R_0$)
2. Overdispersion
3. Primary attack rate
4. Proportion of symptomatic cases
5. Evolutionary rate
6. Substitution rate

| Parameter | Uncertainty | Disaggregated By | Gene | Method | Sample Size | Country | Study Dates | Study Setting | Study Group | Source |
|---|---|---|---|---|---|---|---|---|---|---|
| **Reproduction number R0** | | | | | | | | | | |
| 0.2 | 95% CI: 0.1 - 0.4 | Time | | Empirical (contact tracing) | | Bangladesh | 2007 - 2018 | Unspecified | Persons Under Investigation | Nikolay (2020) |
| 0.33 | 95% CI: 0.19 - 0.59 | Age, Method, Sex, Symptoms, Time, Other | | Empirical (contact tracing) | 248 | Bangladesh | Apr 2001 - Apr 2014 | Population | Persons Under Investigation | Nikolay (2019) |
| 0.3333 | | Time | | Compartmental model | | Bangladesh | 2021 | Unspecified | Other | Sikder (2023) |
| 0.48 | | | | Empirical (contact tracing) | | Bangladesh | 2001 - 2007 | Population | Persons Under Investigation | Luby (2009) |
| 0.75 - 0.88 | | Other | | Empirical (contact tracing) | | Bangladesh | Feb 2006 - Sep 2011 | Hospital | Persons Under Investigation | Naser (2015) |
| **Overdispersion** | | | | | | | | | | |
| 21 mnc | | | | | 248 | Bangladesh | Apr 2001 - Apr 2014 | Population | Persons Under Investigation | Nikolay (2019) |
| 22 mnc | | | | | | Bangladesh | 19 Feb 2004 - 17 Apr 2004 | Mixed | Other | Gurley (2007a) |
| **Primary attack rate** | | | | | | | | | | |
| 1.1 % | | | | | | Bangladesh | 11 Jan 2003 - 28 Jan 2003 | Population | General Population | Hsu (2004) |
| 2.1 % | | | | | | Bangladesh | 20 Apr 2001 - 20 May 2001 | Population | Persons Under Investigation | Hsu (2004) |
| **Proportion of symptomatic cases** | | | | | | | | | | |
| 78.3 % | | | | | 279 | India | 2 Jul 2018 - 13 Jul 2018 | Contact | Other | Kumar (2019) |

continued on next page

continued from previous page

| Parameter | Uncertainty | Disaggregated By | Gene | Method | Sample Size | Country | Study Dates | Study Setting | Study Group | Source |
|---|---|---|---|---|---|---|---|---|---|---|
| 96 % | | | | | 25 | Malaysia | 1998 | Community | Other | Ong (2025) |
| 100 % | | | | | | Bangladesh | 19 Feb 2004 - 17 Apr 2004 | Mixed | Other | Gurley (2007a) |
| **Evolutionary rate** | | | | | | | | | | |
| 4.64 s/s/y $10^{-4}$ | HPDI 95%: 3.43 - 6.04 | | Nucleocapsid | | 61 | Bangladesh, Cambodia, India, Malaysia, Thailand | 1999 - 2018 | Unspecified | Unspecified | Rahman (2021) |
| 6.5 s/s/y $10^{-4}$ | HPDI 95%: 2.3 - 11.8 | | Nucleocapsid | | 29 | Bangladesh, Cambodia, India, Malaysia, Thailand | 1999 - 2010 | Mixed | Mixed Groups | Lo Presti (2016) |
| **Substitution rate** | | | | | | | | | | |
| 4.5 s/s/y $10^{-4}$ | 95% CI: 2.9 - 6 | | Nucleocapsid | | | Bangladesh, Cambodia, India, Indonesia, Malaysia, Thailand | 1999 - 2020 | Unspecified | Unspecified | Cortes-Azuero (2024) |
| 11 s/s/y $10^{-4}$ | HPDI 95%: 7.37 - 15 | Other | Nucleocapsid | | 113 | Bangladesh, Cambodia, India, Malaysia, Thailand | 1999 - 2018 | Unspecified | Unspecified | Li (2020) |
| 2.18 s/s/y $10^{-4}$ | HPDI 95%: 1.6 - 2.8 | Method, Other | Whole genome | | 63 | Bangladesh, India, Malaysia | 1999 - 2018 | Unspecified | Unspecified | Whitmer (2021) |

Table B.10: Transmission parameters grouped by parameter type. Study characteristics are reported for each parameter estimate. Note that s/s/y stands for substitutions per site per year and mnc stands for maximum number of secondary cases caused by an infector. If an exponent is stated for the parameter value the same exponent applies for the uncertainty parameters (without stating it again). Within each parameter type, parameters are sorted in ascending order of value.

##### B.3.4 Natural history

Table B.11 lists all extracted natural history parameters. These are parameters capturing delays from one epidemiological period/event to another. These are given in order of:

1. Incubation period
2. Symptom onset to admission to care
3. Symptom onset to death
4. Symptom onset to recovery/death
5. Symptom onset to discharge/recovery
6. Admission to care to death
7. Admission to care to discharge/recovery
8. Serial interval
9. Other delays categorised as symptom onset to severe illness:
   (a) Symptom onset to respiratory difficulty
   (b) Symptom onset to intubation
   (c) Symptom onset to fever with altered mental status
   (d) Symptom onset to nadir
   (e) Symptom onset to lymphopaenia
   (f) Symptom onset to thrombocytopaenia
10. Other delays:
   (a) Symptom onset to diagnosis/test result
   (b) Duration of ventilation
   (c) Duration of severe illness
   (d) Duration of febrile illness
   (e) Duration between initial neurological episodes
   (f) Exposure - neurological episode

| Delay | Statistic | Uncertainty | Variability | Disaggregated By | Sample Size | Country | Study Dates | Study Setting | Study Group | Source |
|---|---|---|---|---|---|---|---|---|---|---|
| **Incubation period** | | | | | | | | | | |
| 4 days | Median | | Range: 2 - 7 days | | 6 | Bangladesh | 6 Feb 2008 - 10 Mar 2008 | Population | Persons Under Investigation | Rahman (2012) |
| 5 days | Mean | | Other: 0 days | | 4 | Malaysia | Sep 1998 - May 1999 | Hospital | Persons Under Investigation | Chong (2001a) |
| 8 days | Median | | Range: 3 - 20 days | Occupation | 15 | Philippines | 3 Mar 2014 - 24 May 2014 | Hospital | Persons Under Investigation | Ching (2015) |
| 9 days | Median | | IQR: 8 - 11 days | | 82 | Bangladesh | Apr 2001 - Apr 2014 | Population | Persons Under Investigation | Nikolay (2019) |
| 9 days | Median | | Range: 6 - 14 days | | 11 | Bangladesh | Apr 2001 - Apr 2014 | Population | Persons Under Investigation | Nikolay (2019) |
| 9 days | Median | | Range: 6 - 11 days | | 14 | Bangladesh | 2001 - 2007 | Contact | Persons Under Investigation | Luby (2009) |
| 9 days | Median | | Range: 6 - 11 days | | 11 | Bangladesh | Apr 2001 - Apr 2004 | Hospital | Persons With Symptoms | Hossain (2008) |
| 9.3 days | Mean | Other: 1.9 | Range: 6 - 14 days | | 18 | India | May 2018 | Hospital | Persons Under Investigation | Thomas (2019) |
| 9.4 days | Mean | | Other: 8.8 days | | 123 | Malaysia | Sep 1998 - May 1999 | Hospital | Persons Under Investigation | Chong (2001a) |
| 9.5 days | Median | | Range: 6 - 14 days | | | India | 2 May 2018 - 29 May 2018 | Contact | Mixed Groups | Arunkumar (2019) |
| 9.5 days | Median | | Range: 4 - 14 days | Other | 22 | India | 2 May 2018 - 5 Jun 2018 | Contact | Persons Under Investigation | Pallivalappil (2020) |
| 9.7 days | Mean | | SD: 2.2 days$^{+}$ | | 82 | Bangladesh | Apr 2001 - Apr 2014 | Population | Persons Under Investigation | Nikolay (2019)* |
| 10 days | Median | | Range: 9 - 12 days | | 4 | Bangladesh | 6 Feb 2008 - 10 Mar 2008 | Population | Persons Under Investigation | Rahman (2012) |
| 10 days | Median | | Range: 8 - 15 days | | 11 | India | May 2018 | Hospital | Persons Under Investigation | Chandni (2019) |
| 10 days | Unspecified | | Range: 5 - 20 days | | 43 | India | 31 Jan 2001 - 23 Feb 2001 | Population | Persons Under Investigation | Harit (2006) |
| 10 days | Mean | | Other: 8.7 days | | 49 | Malaysia | 26 Dec 1998 - 21 Apr 1999 | Hospital | Persons Under Investigation | Chong (2002) |
| 13 days | Mean | | Range: 2 - 30 days | | 6 | Malaysia | Sep 1998 - May 1999 | Hospital | Persons Under Investigation | Sim (2002) |
| | Unspecified | | Range: 12 - 14 days | | 4 | India | 9 Apr 2007 - 28 Apr 2007 | Contact | Persons Under Investigation | Arankalle (2011) |
| | Unspecified | | Range: 6 - 12 days | Age, Sex | 9 | India | May 2018 - Unspecified | Hospital | Persons Under Investigation | Thulaseedaran (2018) |
| | Unspecified | | Range: 4 - 62 days | | | Malaysia | 29 Sep 1998 - Dec 1999 | Population | Persons Under Investigation | Chua (2003) |
| | Unspecified | | Other: 14 days | | | Malaysia | 29 Sep 1998 - Dec 1999 | Population | Persons Under Investigation | Chua (2003) |
| **Onset - admission** | | | | | | | | | | |
| 3.1 days | Mean | | Range: 1 - 5 days | | 20 | Malaysia | Unspecified | Hospital | Persons Under Investigation | Tan (2002) |

continued on next page

continued from previous page

| Delay | Statistic | Uncertainty | Variability | Disaggregated By | Sample Size | Country | Study Dates | Study Setting | Study Group | Source |
|---|---|---|---|---|---|---|---|---|---|---|
| 3.3 days | Mean | | Range: 1 - 7 days | Age, Sex, Other | 31 | Malaysia | 1998 - 1999 | Hospital | Persons With Symptoms | Wong (2002) |
| 3.5 days | Mean | | Range: 1 - 14 days | | 94 | Malaysia | Feb 1999 - Jun 1999 | Hospital | Persons Under Investigation | Goh (2000) |
| 4 days | Median | | IQR: 3 - 6 days | | 222 | Bangladesh | Apr 2001 - Apr 2014 | Population | Persons Under Investigation | Nikolay (2019) |
| 4 days | Median | | IQR: 1 - 7 days | | 11 | India | May 2018 | Hospital | Persons Under Investigation | Chandni (2019) |
| 4 days | Mean | | | | 142 | Malaysia | Dec 1998 - 31 Dec 1999 | Hospital | Persons Under Investigation | Ramasundrum (2000) |
| | Unspecified | | Range: 1 - 13 days | Age, Occupation, Sex, Symptoms, Other | 18 | Malaysia | Sep 1998 - May 1999 | Hospital | Persons Under Investigation | Sim (2002) |
| **Onset - death** | | | | | | | | | | |
| 4 days | Mean | | Range: 1 - 7 days | | 5 | Bangladesh | Mar 2007 - Apr 2007 | Community | Persons With Symptoms | Homaira (2010b) |
| 4 days | Mean | | Range: 2 - 7 days | | 8 | Bangladesh | 11 Jan 2003 - 28 Jan 2003 | Population | Persons Under Investigation | Hsu (2004) |
| 5 days | Median | | Range: 3 - 17 days | Disease Generation | 14 | Bangladesh | 7 Jan 2010 - 28 Jan 2010 | Community | Persons Under Investigation | Sazzad (2013) |
| 5 days | Median | | Range: 4 - 9 days | | 12 | Bangladesh | 15 Dec 2004 - 31 Jan 2005 | Hospital | Persons With Symptoms | Luby (2006) |
| 5.5 days | Median | | Range: 1 - 47 days | Region | 43 | Bangladesh | 23 Dec 2010 - 1 Mar 2011 | Population | Persons Under Investigation | Chakraborty (2016) |
| 5.6 days | Mean | | Range: 5 - 7 days | | 3 | Bangladesh | 15 Jan 2007 - 28 Feb 2007 | Community | Persons With Symptoms | Homaira (2010a) |
| 6 days | Mean | | Range: 1 - 10 days | | 9 | Bangladesh | 6 Feb 2008 - 10 Mar 2008 | Population | Persons Under Investigation | Rahman (2012) |
| 6 days | Median | | | | 8 | Bangladesh | Jan 2011 - Feb 2014 | Population | Persons Under Investigation | Islam (2016) |
| 6 days | Median | | IQR: 4 - 8 days | | 248 | Bangladesh | Apr 2001 - Apr 2014 | Population | Persons Under Investigation | Nikolay (2019) |
| 6 days | Mean | | | | 9 | Bangladesh | 20 Apr 2001 - 20 May 2001 | Population | Persons Under Investigation | Hsu (2004) |
| 7 days | Mean | | SD: 4.6 days | | 67 | Bangladesh | Apr 2001 - Apr 2004 | Hospital | Persons With Symptoms | Hossain (2008) |
| 9 days | Median | | Range: days | | 74 | Malaysia | Sep 1998 - Jun 1999 | Hospital | Persons Under Investigation | Chong (2001b) |
| 9.5 days | Mean | | Range: 2 - 34 days | | 31 | Malaysia | 1998 - 1999 | Hospital | Persons With Symptoms | Wong (2002) |
| 10 days | Median | | Range: 4 - 39 days | | 18 | Malaysia | Sep 1998 - May 1999 | Hospital | Persons Under Investigation | Sim (2002) |
| 10 days | Unspecified | | Other: 6.8 days | | | Malaysia | 26 Dec 1998 - 21 Apr 1999 | Hospital | Persons Under Investigation | Chong (2002) |

continued on next page

continued from previous page

| Delay | Statistic | Uncertainty | Variability | Disaggregated By | Sample Size | Country | Study Dates | Study Setting | Study Group | Source |
|---|---|---|---|---|---|---|---|---|---|---|
| 10.3 days | Mean | | Range: 5 - 29 days | | 94 | Malaysia | Feb 1999 - Jun 1999 | Hospital | Persons Under Investigation | Goh (2000) |
| 16 days | Mean | | Range: 4 - 39 days | | 18 | Malaysia | Sep 1998 - May 1999 | Hospital | Persons Under Investigation | Sim (2002) |
| 7 - 30 days | Unspecified | | | | | India | 31 Jan 2001 - 23 Feb 2001 | Hospital | Persons Under Investigation | Chadha (2006) |
| | Unspecified | | Range: 2 - 6 days | | 5 | India | 9 Apr 2007 - 28 Apr 2007 | Contact | Persons Under Investigation | Arankalle (2011) |
| | Unspecified | | Range: 5 - 6 days | | | India | May 2018 - Unspecified | Hospital | Persons Under Investigation | Thulaseedaran (2018) |
| **Onset - recovery/death** | | | | | | | | | | |
| 6.4 days | Mean | | | | 19 | India | 2 May 2018 - 5 Jun 2018 | Contact | Persons Under Investigation | Pallivalappil (2020) |
| **Onset - discharge/recovery** | | | | | | | | | | |
| 4 - 83 days | Mean | | | Other | 142 | Malaysia | Dec 1998 - 31 Dec 1999 | Hospital | Persons Under Investigation | Ramasundrum (2000) |
| **Admission - death** | | | | | | | | | | |
| 2 days | Median | | IQR: 1 - 3 days | | 193 | Bangladesh | Apr 2001 - Apr 2014 | Population | Persons Under Investigation | Nikolay (2019) |
| 2 days | Median | | Other: 4 days | | 10 | India | May 2018 | Hospital | Persons Under Investigation | Chandni (2019) |
| 3.4 days | Mean | | Range: 1 - 9 days | Other | 18 | India | May 2018 | Hospital | Other | Thomas (2019) |
| **Admission - discharge/recovery** | | | | | | | | | | |
| 18.8 days | Mean | | Other: 33.5 days | | 184 | Malaysia | Sep 1998 - May 1999 | Hospital | Persons Under Investigation | Chong (2001a) |
| 38.4 days | Mean | | Other: 95.8 days | | 10 | Malaysia | Sep 1998 - May 1999 | Hospital | Persons Under Investigation | Chong (2001a) |
| **Time in care (length of stay)** | | | | | | | | | | |
| 7.3 - 24.7 days | Unspecified | | | Other | 194 | Malaysia | Sep 1998 - Jun 1999 | Hospital | Persons Under Investigation | Chong (2001b) |
| **Serial interval** | | | | | | | | | | |
| 12.7 days | Mean | | SD: 3 days | | 79 | Bangladesh | Apr 2001 - Apr 2014 | Population | Persons Under Investigation | Nikolay (2019)* |
| 13 days | Median | | IQR: 12 - 14 days | | 79 | Bangladesh | Apr 2001 - Apr 2014 | Population | Persons Under Investigation | Nikolay (2019) |
| **Onset - respiratory difficulty** | | | | | | | | | | |
| 4 days | Median | | Range: 0 - 13 days | | | Bangladesh | Apr 2001 - Apr 2004 | Hospital | Persons With Symptoms | Hossain (2008) |
| **Onset - intubation** | | | | | | | | | | |

continued on next page

continued from previous page

| Delay | Statistic | Uncertainty | Variability | Disaggregated By | Sample Size | Country | Study Dates | Study Setting | Study Group | Source |
|---|---|---|---|---|---|---|---|---|---|---|
| 5.8 days | Unspecified | | Other: 4.2 days | | 62 | Malaysia | 26 Dec 1998 - 21 Apr 1999 | Hospital | Persons Under Investigation | Chong (2002) |
| **Onset - fever with altered mental status** | | | | | | | | | | |
| 4 days | Median | | Range: 0 - 10 days | | 82 | Bangladesh | Apr 2001 - Apr 2004 | Hospital | Persons With Symptoms | Hossain (2008) |
| **Onset - nadir** | | | | | | | | | | |
| 6.9 days | Mean | | Range: 3 - 31 days | | | Malaysia | Feb 1999 - Jun 1999 | Hospital | Persons Under Investigation | Goh (2000) |
| **Onset - lymphopaenia** | | | | | | | | | | |
| 5.8 days | Unspecified | | Other: 4.8 days | | 62 | Malaysia | 26 Dec 1998 - 21 Apr 1999 | Hospital | Persons Under Investigation | Chong (2002) |
| **Onset - thrombocytopaenia** | | | | | | | | | | |
| 6 days | Unspecified | | Other: 4.3 days | | 62 | Malaysia | 26 Dec 1998 - 21 Apr 1999 | Hospital | Persons Under Investigation | Chong (2002) |
| **Onset - diagnosis/test result** | | | | | | | | | | |
| | Unspecified | | Range: 1 - 10 days | Other | | India | 31 Jan 2001 - 23 Feb 2001 | Hospital | Persons Under Investigation | Chadha (2006) |
| **Duration of ventilation** | | | | | | | | | | |
| 6.6 days | Unspecified | | Other: 5.4 days | | 62 | Malaysia | 26 Dec 1998 - 21 Apr 1999 | Hospital | Persons Under Investigation | Chong (2002) |
| 3.8 - 10.1 days | Unspecified | | | Other | 194 | Malaysia | Sep 1998 - Jun 1999 | Hospital | Persons Under Investigation | Chong (2001b) |
| **Duration of severe illness** | | | | | | | | | | |
| 2 days | Median | | IQR: 1 - 4 days | | 85 | Bangladesh | Dec 2010 - Apr 2014 | Hospital | Persons Under Investigation | Lee (2020) |
| **Duration of febrile illness** | | | | | | | | | | |
| 3 days | Median | | IQR: 2 - 5 days | | 94 | Bangladesh | Dec 2010 - Apr 2014 | Hospital | Persons Under Investigation | Lee (2020) |
| **Duration between initial neurological episodes** | | | | | | | | | | |
| 7.6 months | Mean | | Range: 1 - 7 months | Age, Sex, Symptoms, Other | 3 | Malaysia | Unspecified | Hospital | Persons Under Investigation | Tan (2002) |
| **Exposure - neurological episode** | | | | | | | | | | |
| 8.4 months | Mean | | Range: 9 - 365 days | Age, Sex, Symptoms, Other | 22 | Malaysia | Unspecified | Hospital | Persons Under Investigation | Tan (2002) |

Table B.11: Extracted natural history estimates grouped by published parameter. Study characteristics are reported for each parameter value. Within each published parameter, parameters are first sorted in ascending order of delay length (the midpoint is used where a range is provided) and second by country.
$^{+}$indicates that uncertainty on the variability value is available; this is not shown in the table for readability.
$*$indicates that the row relates to estimated model parameters; in both cases the rows report the estimate mean and standard deviation of a gamma distribution. See the appendix of Nikolay et al. [8] for more details.

##### B.3.5 CFR

Table B.12 lists all extracted Case Fatality Ratios (CFRs) by country/countries of reporting.

| CFR (%) | Uncertainty | Disaggregated By | Method | Deaths | Sample Size | Study Dates | Study Setting | Study Group | Source |
|---|---|---|---|---|---|---|---|---|---|
| **Bangladesh** | | | | | | | | | |
| 100.0 | | | Unspecified | 4 | 4 | Unspecified | Unspecified | Unspecified | Kulkarni (2013) |
| 100.0 | | | Unspecified | 1 | 1 | Unspecified | Unspecified | Unspecified | Kulkarni (2013) |
| 95.0 | | | Naive | 21 | 22 | Sep 2013 - Mar 2017 | Hospital | Persons Under Investigation | Das (2019) |
| 92.0 | | | Unspecified | 11 | 12 | Unspecified | Unspecified | Unspecified | Kulkarni (2013) |
| 92.0 | | | Unspecified | 11 | 12 | 15 Dec 2004 - 31 Jan 2005 | Hospital | Persons With Symptoms | Luby (2006) |
| 91.0 | | | Unspecified | 40 | 44 | Unspecified | Unspecified | Unspecified | Kulkarni (2013) |
| 90.0 | | | Adjusted | 9 | 10 | 6 Feb 2008 - 10 Mar 2008 | Population | Persons Under Investigation | Rahman (2012) |
| 89.0 | | | Naive | 49 | 55 | 2007 - 2014 | Hospital | Mixed Groups | Hegde (2019) |
| 88.0 | | Disease Generation | Naive | 14 | 16 | 7 Jan 2010 - 28 Jan 2010 | Community | Persons Under Investigation | Sazzad (2013) |
| 88.0 | | Region | Naive | 38 | 43 | 23 Dec 2010 - 1 Mar 2011 | Population | Persons Under Investigation | Chakraborty (2016) |
| 87.5 | | | Unspecified | 14 | 16 | Unspecified | Unspecified | Unspecified | Kulkarni (2013) |
| 86.0 | 95.0% CI: 79.0 - 92.0 | Time | Unspecified | | | 2007 - 2018 | Unspecified | Unspecified | Nikolay (2020) |
| 85.0 | | | Naive | 80 | 94 | Dec 2010 - Apr 2014 | Hospital | Persons Under Investigation | Lee (2020) |
| 83.0 | | | Unspecified | 10 | 12 | Unspecified | Unspecified | Unspecified | Kulkarni (2013) |
| 83.0 | | | Unspecified | 10 | 12 | Unspecified | Unspecified | Unspecified | Kulkarni (2013) |
| 78.0 | | | Naive | 193 | 248 | Apr 2001 - Apr 2014 | Population | Persons Under Investigation | Nikolay (2019) |
| 76.0 | | | Naive | 68 | 90 | 2007 - 2014 | Hospital | Mixed Groups | Hegde (2019) |
| 75.0 | | | Unspecified | 27 | 36 | 19 Feb 2004 - 17 Apr 2004 | Mixed | Persons With Symptoms | Gurley (2007a) |
| 75.0 | | | Unspecified | 27 | 36 | Unspecified | Unspecified | Unspecified | Kulkarni (2013) |
| 74.0 | | | Unspecified | 23 | 31 | Unspecified | Unspecified | Unspecified | Kulkarni (2013) |
| 73.0 | | Method, Region, Symptoms, Time | Naive | 67 | 92 | Apr 2001 - Apr 2004 | Hospital | Persons With Symptoms | Hossain (2008) |
| 71.0 | | | Unspecified | 5 | 7 | Unspecified | Unspecified | Unspecified | Kulkarni (2013) |
| 71.0 | | | Naive | 228 | 322 | Apr 2001 - 2021 | Hospital | Persons With Symptoms | Satter (2023) |
| 71.0 | | | Naive | 87 | 122 | 2001 - 2007 | Population | Persons Under Investigation | Luby (2009) |
| 69.0 | | | Unspecified | 9 | 13 | Unspecified | Unspecified | Unspecified | Kulkarni (2013) |

continued on next page

continued from previous page

| CFR (%) | Uncertainty | Disaggregated By | Method | Deaths | Sample Size | Study Dates | Study Setting | Study Group | Source |
|---|---|---|---|---|---|---|---|---|---|
| 69.0 | | | Adjusted | 9 | 13 | 20 Apr 2001 - 20 May 2001 | Population | Persons Under Investigation | Hsu (2004) |
| 67.0 | | | Unspecified | 8 | 12 | Unspecified | Unspecified | Unspecified | Kulkarni (2013) |
| 67.0 | | | Adjusted | 8 | 12 | 11 Jan 2003 - 28 Jan 2003 | Population | Persons Under Investigation | Hsu (2004) |
| 63.0 | | | Unspecified | 5 | 8 | Unspecified | Unspecified | Unspecified | Kulkarni (2013) |
| 62.5* | | | Naive | 5 | 8 | Mar 2007 - Apr 2007 | Community | Persons With Symptoms | Homaira (2010b) |
| 57.1* | | Disease Generation | Unspecified | 8 | 14 | Jan 2011 - Feb 2014 | Population | Persons Under Investigation | Islam (2016) |
| 46.0 | 95.0% CI: 29.0 - 63.0 | Time | Unspecified | | | 2007 - 2018 | Unspecified | Unspecified | Nikolay (2020) |
| 43.0 | | | Unspecified | 3 | 7 | Unspecified | Unspecified | Unspecified | Kulkarni (2013) |
| 42.9* | | | Unspecified | 3 | 7 | 15 Jan 2007 - 28 Feb 2007 | Community | Persons With Symptoms | Homaira (2010a) |
| 33.0 | | | Unspecified | 1 | 3 | Unspecified | Unspecified | Unspecified | Kulkarni (2013) |
| 24.4* | | | Naive | 44 | 180 | 2007 - 2014 | Hospital | Persons Under Investigation | Hegde (2019) |
| 0.0 | | | Unspecified | 0 | 3 | Unspecified | Unspecified | Unspecified | Kulkarni (2013) |
| **Bangladesh, India** | | | | | | | | | |
| 75.7 | | Region, Time | Unspecified | 221 | 292 | Unspecified | Unspecified | Unspecified | Kulkarni (2013) |
| **India** | | | | | | | | | |
| 100.0 | | | Unspecified | 5 | 5 | 9 Apr 2007 - 28 Apr 2007 | Contact | Persons Under Investigation | Arankalle (2011) |
| 100.0 | | | Unspecified | 5 | 5 | Unspecified | Unspecified | Unspecified | Kulkarni (2013) |
| 91.3 | | | Naive | 21 | 23 | 2 May 2018 - 5 Jun 2018 | Contact | Persons Under Investigation | Pallivalappil (2020) |
| 91.0 | | | Naive | 21 | 23 | 2 May 2018 - 29 May 2018 | Contact | Mixed Groups | Arunkumar (2019) |
| 88.8 | | | Naive | 16 | 18 | May 2018 | Hospital | Persons Under Investigation | Thomas (2019) |
| 83.3 | 95.0% CI: 54.9 - 97.1 | | Naive | 10 | 12 | May 2018 | Hospital | Persons Under Investigation | Chandni (2019) |
| 74.0 | | | Naive | | 66 | 31 Jan 2001 - 23 Feb 2001 | Hospital | Persons Under Investigation | Chadha (2006) |
| 68.0 | | | Unspecified | 45 | 66 | Unspecified | Unspecified | Unspecified | Kulkarni (2013) |
| 68.0 | | Age | Naive | 45 | 66 | 31 Jan 2001 - 23 Feb 2001 | Population | Persons Under Investigation | Harit (2006) |
| 33.3 | | | Naive | 2 | 6 | 25 Aug 2023 - 29 Sep 2023 | Population | General Population | As (2024) |
| **Malaysia** | | | | | | | | | |
| 94.4* | | | Naive | 17 | 18 | Unspecified | Hospital | Mixed Groups | Chua (2000a) |

continued on next page

continued from previous page

| CFR (%) | Uncertainty | Disaggregated By | Method | Deaths | Sample Size | Study Dates | Study Setting | Study Group | Source |
|---|---|---|---|---|---|---|---|---|---|
| 61.0 | | Age, Occupation, Sex, Symptoms, Other | Naive | 11 | 18 | Sep 1998 - May 1999 | Hospital | Persons Under Investigation | Sim (2002) |
| 53.6* | | | Naive | 15 | 28 | 29 Sep 1998 - Dec 1999 | Population | Persons Under Investigation | Chua (2003) |
| 41.0 | | Age, Symptoms, Other | Unspecified | | | 26 Dec 1998 - 21 Apr 1999 | Hospital | Persons Under Investigation | Chong (2002) |
| 39.6* | | | Naive | 105 | 265 | Sep 1998 - Jun 1999 | Population | Persons Under Investigation | Chua (2000b) |
| 38.5 | | Region | Naive | 109 | 283 | 29 Sep 1998 - Dec 1999 | Population | Persons Under Investigation | Chua (2003) |
| 38.0 | | Other | Adjusted | 74 | 194 | Sep 1998 - Jun 1999 | Hospital | Persons Under Investigation | Chong (2001b) |
| 37.2* | | | Naive | 86 | 231 | 29 Sep 1998 - Dec 1999 | Population | Persons Under Investigation | Chua (2003) |
| 33.3* | | | Naive | 8 | 24 | 29 Sep 1998 - Dec 1999 | Population | Persons Under Investigation | Chua (2003) |
| 32.1* | | | Naive | 27 | 84 | Unspecified | Hospital | Mixed Groups | Chua (2000a) |
| 30.0* | | Symptoms | Naive | 6 | 20 | Unspecified | Hospital | Mixed Groups | Chua (2001) |
| 18.0 | | Age, Sex, Symptoms, Other | Naive | 4 | 22 | Unspecified | Hospital | Persons Under Investigation | Tan (2002) |
| **Philippines** | | | | | | | | | |
| 82.0 | | Occupation | Unspecified | 9 | 11 | 3 Mar 2014 - 24 May 2014 | Hospital | Persons Under Investigation | Ching (2015) |
| 52.9* | | Occupation | Unspecified | 9 | 17 | 3 Mar 2014 - 24 May 2014 | Hospital | Persons Under Investigation | Ching (2015) |
| **Singapore** | | | | | | | | | |
| 9.1* | | | Naive | 1 | 11 | 16 Mar 1999 - Unspecified | Trade/business | Abattoir Workers | Paton (1999) |
| 9.1* | | | Naive | 1 | 11 | Mar 1999 | Population | Persons Under Investigation | Chua (2000b) |

Table B.12: CFRs grouped by country. Study characteristics are reported for each CFR estimate. Within each country (sorted alphabetically), parameters are sorted in descending order of CFRs.
*indicates that only deaths and a sample size were provided, and that the (naïve) CFR shown has been calculated as $\frac{\text{Deaths}}{\text{Sample size}}$.

##### B.3.6 Seroprevalence

Table B.13 lists all extracted seroprevalence estimates by country of reporting.

| Seroprevalence (%) | Uncertainty | Disaggregated By | Assay | Number Seropositive | Sample Size | Study Dates | Study Setting | Study Group | Source |
|---|---|---|---|---|---|---|---|---|---|
| **Bangladesh** | | | | | | | | | |
| 30.8* | | | IgG | 4 | 13 | 20 Apr 2001 - 20 May 2001 | Hospital | Persons Under Investigation | Hsu (2004) |
| 20.0* | | | IgG | 2 | 10 | Feb 2008 | Unspecified | Persons Under Investigation | Lo (2012) |
| 11.8* | | | IgG | 2 | 17 | Dec 2009 - Mar 2010 | Unspecified | Persons Under Investigation | Lo (2012) |
| 1.9* | | | IgG | 2 | 105 | 24 Mar 2004 - 30 Mar 2004 | Hospital | Healthcare Workers | Gurley (2007b) |
| 82.4* | | Other | IgM | 42 | 51 | Apr 2001 - Apr 2004 | Hospital | Persons With Symptoms | Hossain (2008) |
| 75.0* | | | IgM | 3 | 4 | Mar 2007 - Apr 2007 | Community | Persons With Symptoms | Homaira (2010b) |
| 70.6* | | | IgM | 12 | 17 | Dec 2009 - Mar 2010 | Unspecified | Persons Under Investigation | Lo (2012) |
| 45.5* | | | IgM | 5 | 11 | 15 Jan 2007 - 28 Feb 2007 | Community | Persons With Symptoms | Homaira (2010a) |
| 40.0* | | | IgM | 4 | 10 | Feb 2008 | Unspecified | Persons Under Investigation | Lo (2012) |
| 22.0 | | | IgM | 28 | 127 | Feb 2006 - Sep 2011 | Hospital | Mixed Groups | Naser (2015) |
| 12.0 | | | IgM | 12 | 106 | Jan 2010 - Apr 2010 | Population | Persons Under Investigation | Sazzad (2013) |
| 8.0 | | | IgM | 22 | 268 | Sep 2013 - Mar 2017 | Hospital | Persons Under Investigation | Das (2019) |
| 4.0 | | Time | IgM | 23 | 608 | Feb 2006 - Sep 2011 | Hospital | Persons With Symptoms | Naser (2015) |
| 3.0 | | | IgM | 9 | 312 | Dec 2013 - Apr 2014 | Hospital | Persons With Symptoms | Hassan (2018) |
| 0.0* | | | IgM | 0 | 13 | 20 Apr 2001 - 20 May 2001 | Hospital | Persons Under Investigation | Hsu (2004) |
| 0.0* | | | IgM | 0 | 105 | 24 Mar 2004 - 30 Mar 2004 | Hospital | Healthcare Workers | Gurley (2007b) |
| 0.0* | | | IgM | 0 | 11 | 24 Mar 2004 - 30 Mar 2004 | Hospital | Other | Gurley (2007b) |
| 100.0* | | | Unspecified | 4 | 4 | 18 Feb 2004 - 22 Feb 2004 | Community | Persons With Symptoms | Montgomery (2008) |
| 14.8* | | | Unspecified | 4 | 27 | 19 Feb 2004 - 17 Apr 2004 | Mixed | Persons With Symptoms | Gurley (2007a) |
| 1.4 | | | Unspecified | 35 | 2494 | Apr 2007 - Apr 2014 | Contact | Persons Under Investigation | Nikolay (2019) |
| 0.0* | | | Unspecified | 0 | 1863 | Apr 2007 - Apr 2014 | Contact | Persons Under Investigation | Nikolay (2019) |

continued on next page

continued from previous page

| Seroprevalence (%) | Uncertainty | Disaggregated By | Assay | Number Seropositive | Sample Size | Study Dates | Study Setting | Study Group | Source |
|---|---|---|---|---|---|---|---|---|---|
| 0.0* | | | Unspecified | 0 | 13 | 18 Feb 2004 - 22 Feb 2004 | Community | General Population | Montgomery (2008) |
| **Cambodia** | | | | | | | | | |
| 0.0* | | | Unspecified | 0 | 164 | May 2016 - Sep 2016 | Mixed | Mixed Groups | Cappelle (2020) |
| 0.0* | | | Unspecified | 0 | 254 | May 2016 - Sep 2016 | Mixed | Mixed Groups | Cappelle (2020) |
| **Cameroon** | | | | | | | | | |
| 3.0 | | | PRNT | 7 | 227 | Feb 2001 - Jan 2003 | Community | Other | Pernet (2014) |
| 0.0 | | | PRNT | 0 | 260 | Feb 2001 - Jan 2003 | Community | Other | Pernet (2014) |
| **India** | | | | | | | | | |
| 58.8* | | | IgG | 10 | 17 | 31 Jan 2001 - 23 Feb 2001 | Hospital | Persons Under Investigation | Chadha (2006) |
| 22.2* | | Age, Level of Exposure, Sex, Symptoms, Time, Other | IgG | 4 | 18 | 2 May 2018 - 29 May 2018 | Contact | Mixed Groups | Arunkumar (2019) |
| 2.8* | | | IgG | 1 | 36 | 9 Apr 2007 - 28 Apr 2007 | Contact | Persons Under Investigation | Arankalle (2011) |
| 0.0* | | | IgG | 0 | 49 | Sep 2019 | Contact | Persons Under Investigation | Ramachandran (2022) |
| 0.0* | | | IgG | 0 | 9 | May 2018 | Contact | Persons Under Investigation | Yadav (2019) |
| 0.0* | | | IgG | 0 | 5 | 5 Sep 2023 - 29 Sep 2023 | Hospital | Persons Under Investigation | As (2024) |
| 0.0 | | Level of Exposure, Symptoms | IgG | | | Sep 2023 - Unspecified | Contact | Mixed Groups | As (2024) |
| 0.0* | | | IgG | 0 | 130 | Sep 2023 - Unspecified | Population | Persons Under Investigation | As (2024) |
| 0.0* | | | IgG | 0 | 61 | 6 Sep 2021 - Unspecified | Population | Persons Under Investigation | Yadav (2022) |
| 0.0* | | Level of Exposure, Symptoms | IgG | 0 | 64 | 6 Sep 2021 - Unspecified | Contact | Persons Under Investigation | Yadav (2022) |
| 83.3* | | | IgM | 5 | 6 | May 2018 | Contact | Persons Under Investigation | Yadav (2019) |
| 80.0* | | | IgM | 4 | 5 | 5 Sep 2023 - 29 Sep 2023 | Hospital | Persons Under Investigation | As (2024) |
| 72.2* | | Age, Level of Exposure, Sex, Symptoms, Time, Other | IgM | 13 | 18 | 2 May 2018 - 29 May 2018 | Contact | Mixed Groups | Arunkumar (2019) |
| 52.9* | | | IgM | 9 | 17 | 31 Jan 2001 - 23 Feb 2001 | Hospital | Persons Under Investigation | Chadha (2006) |

continued on next page

continued from previous page

| Seroprevalence (%) | Uncertainty | Disaggregated By | Assay | Number Seropositive | Sample Size | Study Dates | Study Setting | Study Group | Source |
|---|---|---|---|---|---|---|---|---|---|
| 5.6* | | | IgM | 2 | 36 | 9 Apr 2007 - 28 Apr 2007 | Contact | Persons Under Investigation | Arankalle (2011) |
| 0.0* | | | IgM | 0 | 49 | Sep 2019 | Contact | Persons Under Investigation | Ramachandran (2022) |
| 0.0 | | Level of Exposure, Symptoms | IgM | | | Sep 2023 - Unspecified | Contact | Mixed Groups | As (2024) |
| 0.0* | | | IgM | 0 | 130 | Sep 2023 - Unspecified | Population | Persons Under Investigation | As (2024) |
| 0.0* | | | IgM | 0 | 61 | 6 Sep 2021 - Unspecified | Population | Persons Under Investigation | Yadav (2022) |
| 0.0* | | Level of Exposure, Symptoms | IgM | 0 | 64 | 6 Sep 2021 - Unspecified | Contact | Persons Under Investigation | Yadav (2022) |
| 1.1 | 95.0% CI: 0.4 - 3.1 | Other | Unspecified | 3 | 279 | 2 Jul 2018 - 13 Jul 2018 | Contact | Other | Kumar (2019) |
| **Malaysia** | | | | | | | | | |
| 100.0* | | | IgG | 22 | 22 | Unspecified | Hospital | Persons Under Investigation | Tan (2002) |
| 92.3*+ | | | IgG | 12 | 13 | Unspecified | Hospital | Persons Under Investigation | Tan (2002) |
| 84.0 | | Other | IgG | 21 | 25 | Unspecified | Community | Other | Ong (2025) |
| 7.0 - 100.0 | | Symptoms, Time, Other | IgG | | 142 | Dec 1998 - 31 Dec 1999 | Hospital | Persons Under Investigation | Ramasundrum (2000) |
| 29.0+ | | Age, Sex, Other | IgG | 9 | 31 | 1998 - 1999 | Hospital | Persons With Symptoms | Wong (2002) |
| 12.5 | 95.0% CI: 1.6 - 38.4 | Age, Sex, Other | IgG | 2 | 16 | Unspecified | Community | General Population | Yong (2020) |
| 11.7 | 95.0% CI: 6.0 - 20.0 | Age, Sex, Other | IgG | 11 | 94 | Unspecified | Community | General Population | Yong (2020) |
| 9.0 | 95.0% CI: 3.4 - 18.5 | Age, Sex, Other | IgG | 6 | 67 | Unspecified | Community | General Population | Yong (2020) |
| 1.6 | | Level of Exposure, Occupation, Region | IgG | 7 | 435 | 6 Apr 1999 - 20 Apr 1999 | Trade/business | Abattoir Workers | Sahani (2001) |
| 1.0* | | | IgG | 3 | 293 | 1999 | Hospital | Persons Under Investigation | Mounts (2001) |
| 0.0 | 95.0% CI: 0.0 - 1.9 | | IgG | 0 | 153 | 20 Sep 2001 - 27 Apr 2002 | Community | General Population | Chong (2003) |
| 0.0* | | | IgG | 0 | 363 | 2 Mar 1999 | Hospital | Persons Under Investigation | Mounts (2001) |
| 0.0* | | | IgG | 0 | 288 | 2 Mar 1999 | Hospital | Healthcare Workers | Mounts (2001) |
| 100.0* | | Age, Occupation, Sex, Symptoms, Other | IgM | 17 | 17 | Sep 1998 - May 1999 | Hospital | Persons Under Investigation | Sim (2002) |

continued on next page

continued from previous page

| Seroprevalence (%) | Uncertainty | Disaggregated By | Assay | Number Seropositive | Sample Size | Study Dates | Study Setting | Study Group | Source |
|---|---|---|---|---|---|---|---|---|---|
| 90.0+ | | Age, Sex, Other | IgM | 28 | 31 | 1998 - 1999 | Hospital | Persons With Symptoms | Wong (2002) |
| 85.0* | | Age, Sex, Other | IgM | 17 | 20 | Unspecified | Hospital | Mixed Groups | Chua (2001) |
| 81.0 | | | IgM | 91 | 103 | 26 Dec 1998 - 21 Apr 1999 | Hospital | Persons Under Investigation | Chong (2002) |
| 44.0 - 100.0 | | Symptoms, Time | IgM | | 142 | Dec 1998 - 31 Dec 1999 | Hospital | Persons Under Investigation | Ramasundrum (2000) |
| 58.0+ | | | IgM | 31 | | 26 Dec 1998 - 21 Apr 1999 | Hospital | Persons Under Investigation | Chong (2002) |
| 45.5* | | | IgM | 10 | 22 | Unspecified | Hospital | Persons Under Investigation | Tan (2002) |
| 30.0*+ | | Age, Occupation, Sex, Symptoms, Other | IgM | 3 | 10 | Sep 1998 - May 1999 | Hospital | Persons Under Investigation | Sim (2002) |
| 15.4*+ | | | IgM | 2 | 13 | Unspecified | Hospital | Persons Under Investigation | Tan (2002) |
| 1.4* | | Occupation, Region | IgM | 6 | 435 | 6 Apr 1999 - 20 Apr 1999 | Trade/business | Abattoir Workers | Sahani (2001) |
| 0.0* | | | IgM | 0 | 363 | 2 Mar 1999 | Hospital | Persons Under Investigation | Mounts (2001) |
| 0.0* | | | IgM | 0 | 288 | 2 Mar 1999 | Hospital | Healthcare Workers | Mounts (2001) |
| 0.0* | | | IgM | 0 | 293 | 1999 | Hospital | Persons Under Investigation | Mounts (2001) |
| 72.0 | | | PRNT | 18 | 25 | Unspecified | Community | Other | Ong (2025) |
| 0.0* | | | PRNT | 0 | 3 | 1999 | Hospital | Persons Under Investigation | Mounts (2001) |
| 79.0 | | | Unspecified | 80 | 101 | Mar 1999 | Hospital | Persons With Symptoms | Parashar (2000) |
| 75.4 | | | Unspecified | 52 | 69 | 1 Mar 1999 - 15 May 1999 | Hospital | Persons With Symptoms | Amal (2000) |
| 56.5* | | | Unspecified | 39 | 69 | Unspecified | Hospital | Persons With Symptoms | Chua (2000a) |
| 28.0*+ | | | Unspecified | 14 | 50 | Unspecified | Hospital | Persons With Symptoms | Chua (2000a) |
| 11.0 | | | Unspecified | 20 | 178 | Mar 1999 | Other | Mixed Groups | Parashar (2000) |
| 6.0 | | | Unspecified | 10 | 166 | Mar 1999 | Community | Mixed Groups | Parashar (2000) |
| 3.6* | | | Unspecified | 1 | 28 | 1999 | Trade/business | Animal Workers | Premalatha (2000) |
| 0.0* | | | Unspecified | 0 | 20 | 1999 | Trade/business | Other | Premalatha (2000) |
| **Philippines** | | | | | | | | | |
| 100.0 | | | PRNT | 3 | 3 | 3 Mar 2014 - 24 May 2014 | Hospital | Persons Under Investigation | Ching (2015) |

continued on next page

continued from previous page

| Seroprevalence (%) | Uncertainty | Disaggregated By | Assay | Number Seropositive | Sample Size | Study Dates | Study Setting | Study Group | Source |
|---|---|---|---|---|---|---|---|---|---|
| **Singapore** | | | | | | | | | |
| 24.1* | | | IgG | 13 | 54 | 22 Feb 1999 - 22 Apr 1999 | Trade/business | Mixed Groups | Chew (2000) |
| 2.9* | | Age | IgG | 1 | 35 | 16 Mar 1999 - Unspecified | Trade/business | Abattoir Workers | Paton (1999) |
| 1.4* | | | IgG | 21 | 1469 | 14 Feb 1999 - Unspecified | Trade/business | Mixed Groups | Chan (2002) |
| 31.4* | | Age | IgM | 11 | 35 | 16 Mar 1999 - Unspecified | Trade/business | Abattoir Workers | Paton (1999) |
| 24.1* | | | IgM | 13 | 54 | 22 Feb 1999 - 22 Apr 1999 | Trade/business | Mixed Groups | Chew (2000) |
| 1.4* | | | IgM | 20 | 1469 | 14 Feb 1999 - Unspecified | Trade/business | Mixed Groups | Chan (2002) |
| 57.1* | | | PRNT | 4 | 7 | 14 Feb 1999 - Unspecified | Trade/business | Abattoir Workers | Chan (2002) |
| **Thailand** | | | | | | | | | |
| 0.0* | | | IgG | 0 | 418 | Nov 2010 - Dec 2010 | Population | General Population | Wacharapluesadee (2021) |
| 0.0* | | | Unspecified | 0 | 128 | Nov 2010 - Dec 2010 | Population | General Population | Wacharapluesadee (2021) |

Table B.13: Extracted seroprevalence estimates grouped by assay type used. Study characteristics are reported for each seroprevalence estimate. Within each country (sorted alphabetically), parameters are sorted first by parameter type (alphabetically) and second by seropositivity in descending order (the midpoint is used where a range is provided).

*indicates that only number seropositive and a sample size were provided, and that the (naïve) prevalence shown has been calculated as $\frac{\text{Number seropositive}}{\text{Sample size}}$.

+indicates that the serology test was performed on a cerebrospinal fluid sample.

##### B.3.7 Risk factors

Table B.14 lists all extracted risk factors, including whether they were found to be significant or not significant. These are given in order of risks associated with:

1. Infection
2. Serology
3. Death
4. Neurological symptoms
5. Other

| Risk Factor | Significance | Method | Sample Size | Country | Study Dates | Study Setting | Study Group | Source |
|---|---|---|---|---|---|---|---|---|
| **Infection** | | | | | | | | |
| Age, Other | Significant | Adjusted | 248 | Bangladesh | Apr 2001 - Apr 2014 | Population | Persons Under Investigation | Nikolay (2019) |
| Close contact | Significant | Adjusted | | Bangladesh | 19 Feb 2004 - 17 Apr 2004 | Mixed | Other | Gurley (2007a) |
| Close contact, Contact with animal, Sex, Other | Significant | Adjusted | 43 | Bangladesh | 23 Dec 2010 - 1 Mar 2011 | Population | Mixed Groups | Chakraborty (2016) |
| Close contact, Other | Significant | Adjusted | | Bangladesh | 2004 - 2012 | Population | Persons Under Investigation | Hegde (2016) |
| Close contact, Other | Significant | Adjusted | 2600 | Bangladesh | Apr 2007 - Apr 2014 | Contact | Persons Under Investigation | Nikolay (2019) |
| Other | Significant | Adjusted | 367 | Bangladesh | 2004 - 2012 | Population | Persons Under Investigation | Hegde (2016) |
| Other | Significant | Adjusted | 10 | Bangladesh | 6 Feb 2008 - 10 Mar 2008 | Population | Persons Under Investigation | Rahman (2012) |
| Age, Close contact, Contact with animal | Significant | Not adjusted | 96 | Bangladesh | 20 Apr 2001 - 20 May 2001 | Population | General Population | Hsu (2004) |
| Age, Sex, Other | Significant | Not adjusted | 248 | Bangladesh | Apr 2001 - Apr 2014 | Population | Persons Under Investigation | Nikolay (2019) |
| Close contact, Contact with animal, Sex, Other | Significant | Not adjusted | 43 | Bangladesh | 23 Dec 2010 - 1 Mar 2011 | Population | Mixed Groups | Chakraborty (2016) |
| Close contact, Environmental, Other | Significant | Not adjusted | | Bangladesh | 19 Feb 2004 - 17 Apr 2004 | Mixed | Other | Gurley (2007a) |
| Close contact, Occupation, Other | Significant | Not adjusted | 73 | Bangladesh | 7 Jan 2010 - 28 Jan 2010 | Community | Mixed Groups | Sazzad (2013) |
| Close contact, Other | Significant | Not adjusted | 7 | Bangladesh | 15 Jan 2007 - 28 Feb 2007 | Community | Persons With Symptoms | Homaira (2010a) |
| Close contact, Other | Significant | Not adjusted | 48 | Bangladesh | 18 Feb 2004 - 22 Feb 2004 | Community | General Population | Montgomery (2008) |

continued on next page

continued from previous page

| Risk Factor | Significance | Method | Sample Size | Country | Study Dates | Study Setting | Study Group | Source |
|---|---|---|---|---|---|---|---|---|
| Close contact, Sex, Other | Significant | Not adjusted | 2600 | Bangladesh | Apr 2007 - Apr 2014 | Contact | Persons Under Investigation | Nikolay (2019) |
| Contact with animal, Occupation | Significant | Not adjusted | | Malaysia | 1 Mar 1999 - 15 May 1999 | Hospital | Persons With Symptoms | Amal (2000) |
| Contact with animal, Occupation, Sex, Other | Significant | Not adjusted | | Malaysia | Mar 1999 | Mixed | Mixed Groups | Parashar (2000) |
| Contact with animal, Other | Significant | Not adjusted | 10 | Bangladesh | 6 Feb 2008 - 10 Mar 2008 | Population | Persons Under Investigation | Rahman (2012) |
| Occupation, Other | Significant | Not adjusted | | Malaysia | Mar 1999 | Mixed | Mixed Groups | Parashar (2000) |
| Close contact, Contact with animal, Occupation, Other | Not significant | Adjusted | 10 | Bangladesh | 6 Feb 2008 - 10 Mar 2008 | Population | Persons Under Investigation | Rahman (2012) |
| Contact with animal, Occupation, Other | Not significant | Adjusted | | Bangladesh | 2004 - 2012 | Population | Persons Under Investigation | Hegde (2016) |
| Contact with animal, Other | Not significant | Adjusted | 43 | Bangladesh | 23 Dec 2010 - 1 Mar 2011 | Population | Mixed Groups | Chakraborty (2016) |
| Environmental, Other | Not significant | Adjusted | | Bangladesh | 19 Feb 2004 - 17 Apr 2004 | Mixed | Other | Gurley (2007a) |
| Age, Close contact | Not significant | Not adjusted | 248 | Bangladesh | Apr 2007 - Apr 2014 | Population | Persons Under Investigation | Nikolay (2019) |
| Age, Sex, Other | Not significant | Not adjusted | | Malaysia | Mar 1999 | Mixed | Mixed Groups | Parashar (2000) |
| Close contact, Contact with animal, Occupation, Other | Not significant | Not adjusted | 10 | Bangladesh | 6 Feb 2008 - 10 Mar 2008 | Population | Persons Under Investigation | Rahman (2012) |
| Close contact, Contact with animal, Sex, Other | Not significant | Not adjusted | 8 | Bangladesh | Mar 2007 - Apr 2007 | Community | Persons With Symptoms | Homaira (2010b) |
| Contact with animal, Environmental, Other | Not significant | Not adjusted | | Bangladesh | 19 Feb 2004 - 17 Apr 2004 | Mixed | Other | Gurley (2007a) |
| Contact with animal, Occupation | Not significant | Not adjusted | | Malaysia | 1 Mar 1999 - 15 May 1999 | Hospital | Persons With Symptoms | Amal (2000) |
| Contact with animal, Occupation, Sex | Not significant | Not adjusted | 96 | Bangladesh | 20 Apr 2001 - 20 May 2001 | Population | General Population | Hsu (2004) |
| Contact with animal, Other | Not significant | Not adjusted | 43 | Bangladesh | 23 Dec 2010 - 1 Mar 2011 | Population | Mixed Groups | Chakraborty (2016) |
| Contact with animal, Other | Not significant | Not adjusted | 48 | Bangladesh | 18 Feb 2004 - 22 Feb 2004 | Community | General Population | Montgomery (2008) |
| Contact with animal, Sex, Other | Not significant | Not adjusted | 73 | Bangladesh | 7 Jan 2010 - 28 Jan 2010 | Community | Mixed Groups | Sazzad (2013) |
| Contact with animal, Sex, Other | Not significant | Not adjusted | 7 | Bangladesh | 15 Jan 2007 - 28 Feb 2007 | Community | Persons With Symptoms | Homaira (2010a) |
| Other | Not significant | Not adjusted | 248 | Bangladesh | Apr 2001 - Apr 2014 | Population | Persons Under Investigation | Nikolay (2019) |
| Other | Not significant | Not adjusted | | Malaysia | Mar 1999 | Mixed | Mixed Groups | Parashar (2000) |
| **Serology** | | | | | | | | |
| Age | Significant | Not adjusted | 177 | Malaysia | Unspecified | Community | General Population | Yong (2020) |
| Contact with animal, Environmental, Other | Significant | Not adjusted | 487 | Cameroon | Feb 2001 - Jan 2003 | Community | Unspecified | Pernet (2014) |

continued on next page

continued from previous page

| Risk Factor | Significance | Method | Sample Size | Country | Study Dates | Study Setting | Study Group | Source |
|---|---|---|---|---|---|---|---|---|
| Contact with animal, Other | Significant | Not adjusted | | Singapore | 22 Feb 1999 - 22 Apr 1999 | Trade/business | Mixed Groups | Chew (2000) |
| Age, Contact with animal, Other | Significant | Unspecified | 22 | Bangladesh | Sep 2013 - Mar 2017 | Hospital | Persons Under Investigation | Das (2019) |
| Other | Significant | Unspecified | 435 | Malaysia | 6 Apr 1999 - 20 Apr 1999 | Trade/business | Abattoir Workers | Sahani (2001) |
| Age, Contact with animal, Sex, Other | Not significant | Not adjusted | 177 | Malaysia | Unspecified | Community | General Population | Yong (2020) |
| Age, Close contact, Contact with animal, Sex, Other | Not significant | Not adjusted | 25 | Malaysia | Unspecified | Community | Other | Ong (2025) |
| Age, Contact with animal, Sex, Other | Not significant | Not adjusted | | Singapore | 22 Feb 1999 - 22 Apr 1999 | Trade/business | Mixed Groups | Chew (2000) |
| Other | Not significant | Not adjusted | 487 | Cameroon | Feb 2001 - Jan 2003 | Community | Unspecified | Pernet (2014) |
| Contact with animal, Other | Not significant | Unspecified | | Malaysia | 6 Apr 1999 - 20 Apr 1999 | Trade/business | Abattoir Workers | Sahani (2001) |
| Sex | Not significant | Unspecified | 22 | Bangladesh | Sep 2013 - Mar 2017 | Hospital | Persons Under Investigation | Das (2019) |
| **Death** | | | | | | | | |
| Age, Other | Significant | Adjusted | 103 | Malaysia | 26 Dec 1998 - 21 Apr 1999 | Hospital | Persons Under Investigation | Chong (2002) |
| Other | Significant | Adjusted | 194 | Malaysia | Sep 1998 - May 1999 | Hospital | Persons Under Investigation | Chong (2001a) |
| Age, Other | Significant | Not adjusted | | Malaysia | Feb 1999 - Jun 1999 | Hospital | Persons Under Investigation | Goh (2000) |
| Age, Other | Significant | Not adjusted | 194 | Malaysia | Sep 1998 - Jun 1999 | Hospital | Persons Under Investigation | Chong (2001b) |
| Age, Other | Significant | Not adjusted | 92 | Bangladesh | Apr 2001 - Apr 2004 | Hospital | Persons With Symptoms | Hossain (2008) |
| Contact with animal | Not significant | Adjusted | 194 | Malaysia | Sep 1998 - May 1999 | Hospital | Persons Under Investigation | Chong (2001a) |
| Sex, Other | Not significant | Not adjusted | 92 | Bangladesh | Apr 2001 - Apr 2004 | Hospital | Persons With Symptoms | Hossain (2008) |
| Other | Not significant | Not adjusted | 145 | Bangladesh | 2007 - 2014 | Hospital | Mixed Groups | Hegde (2019) |
| Other | Not significant | Not adjusted | 12 | Malaysia | Unspecified | Hospital | Persons Under Investigation | Tan (2002) |
| **Neurological symptoms** | | | | | | | | |
| Other | Not significant | Adjusted | 194 | Malaysia | Sep 1998 - Jun 1999 | Hospital | Persons Under Investigation | Chong (2001b) |
| **Spillover** | | | | | | | | |
| Contact with animal, Other | Significant | Adjusted | | Bangladesh | Sep 1998 - 2015 | Population | General Population | Walsh (2015) |
| Environmental, Other | Significant | Adjusted | | Bangladesh | Feb - Dec | Population | Other | Hahn (2014a) |
| Temperature | Significant | Adjusted | | Bangladesh | Apr 2001 - Jun 2019 | Population | General Population | Latinne (2022) |

continued on next page

continued from previous page

| Risk Factor | Significance | Method | Sample Size | Country | Study Dates | Study Setting | Study Group | Source |
|---|---|---|---|---|---|---|---|---|
| Other | Significant | Adjusted | | Bangladesh | Dec 2011 - Feb 2013 | Other | Other | Gurley (2017) |
| Environmental, Other | Significant | Not adjusted | | Bangladesh | Feb - Dec | Population | Other | Hahn (2014a) |
| Environmental, Temperature, Rainfall | Significant | Not adjusted | | Bangladesh | Apr 2001 - May 2019 | Population | General Population | Latinne (2022) |
| Temperature | Significant | Not adjusted | | Bangladesh | 2001 - 2018 | Population | Other | Mckee (2021) |
| Temperature, Rainfall, Other | Significant | Not adjusted | | Bangladesh | Jan 2007 - Dec 2013 | Unspecified | Unspecified | Cortes (2018) |
| Other | Significant | Unspecified | | Bangladesh | Dec 2011 - Feb 2012 | Other | Other | Hahn (2014b) |
| Environmental | Not significant | Adjusted | | Bangladesh | Sep 1998 - 2015 | Population | General Population | Walsh (2015) |
| Environmental, Other | Not significant | Adjusted | | Bangladesh | Feb - Dec | Population | Other | Hahn (2014a) |
| Environmental, Rainfall | Not significant | Adjusted | | Bangladesh | Apr 2001 - Jun 2019 | Population | General Population | Latinne (2022) |
| Other | Not significant | Adjusted | | Bangladesh | Dec 2011 - Feb 2013 | Unspecified | Unspecified | Gurley (2017) |
| Environmental, Other | Not significant | Not adjusted | | Bangladesh | Feb - Dec | Population | Other | Hahn (2014a) |
| Environmental, Other | Not significant | Not adjusted | | Bangladesh | 2001 - 2018 | Population | Other | Mckee (2021) |
| Temperature, Rainfall, Other | Not significant | Not adjusted | | Bangladesh | Jan 2007 - Dec 2013 | Unspecified | Unspecified | Cortes (2018) |
| Environmental | Unspecified | Unspecified | | Bangladesh | Apr 2001 - Jun 2019 | Population | General Population | Latinne (2022) |
| Environmental, Other | Unspecified | Unspecified | | Thailand | Unspecified | Other | Other | Thanapongtharm (2019) |
| **Other** | | | | | | | | |
| Hospitalisation, Sex, Other | Significant | Adjusted | | Bangladesh | Dec 2010 - Apr 2014 | Hospital | Persons Under Investigation | Lee (2020) |
| Other | Significant | Adjusted | | Bangladesh | 15 Dec 2004 - 31 Jan 2005 | Hospital | Unspecified | Luby (2006) |
| Other | Significant | Not adjusted | 84 | Malaysia | Unspecified | Hospital | Persons With Symptoms | Chua (2000a) |
| Other | Significant | Not adjusted | 20 | Malaysia | Unspecified | Hospital | Mixed Groups | Chua (2001) |
| Other | Significant | Unspecified | 279 | India | 2 Jul 2018 - 13 Jul 2018 | Contact | Other | Kumar (2019) |
| Other | Not significant | Adjusted | 194 | Malaysia | Sep 1998 - Jun 1999 | Hospital | Persons Under Investigation | Chong (2001b) |
| Age, Comorbidity, Sex, Other | Not significant | Not adjusted | 20 | Malaysia | Unspecified | Hospital | Mixed Groups | Chua (2001) |
| Sex, Other | Not significant | Not adjusted | 84 | Malaysia | Unspecified | Hospital | Persons With Symptoms | Chua (2000a) |
| Other | Not significant | Not adjusted | 103 | Malaysia | 26 Dec 1998 - 21 Apr 1999 | Hospital | Persons Under Investigation | Chong (2002) |
| Other | Not significant | Not adjusted | 12 | Malaysia | Unspecified | Hospital | Persons Under Investigation | Tan (2002) |

continued on next page

continued from previous page

| Risk Factor | Significance | Method | Sample Size | Country | Study Dates | Study Setting | Study Group | Source |
|---|---|---|---|---|---|---|---|---|
| Contact with animal | Not significant | Unspecified | | Bangladesh | 15 Dec 2004 - 31 Jan 2005 | Hospital | Unspecified | Luby (2006) |
| Age, Hospitalisation, Sex, Other | Unspecified | Unspecified | | Bangladesh | Dec 2010 - Apr 2014 | Hospital | Persons Under Investigation | Lee (2020) |
| Contact with animal, Sex, Other | Unspecified | Unspecified | | Bangladesh | 15 Dec 2004 - 31 Jan 2005 | Hospital | Unspecified | Luby (2006) |
| Other | Unspecified | Unspecified | | Bangladesh | 15 Dec 2004 - 31 Jan 2005 | Hospital | Unspecified | Luby (2006) |

Table B.14: Extracted risk factors grouped by outcome. Study characteristics are reported for each set of reported risk factors. Within each risk factor outcome, parameters are sorted by significance, method, and risk factor (alphabetically).

#### B.4 Additional figures

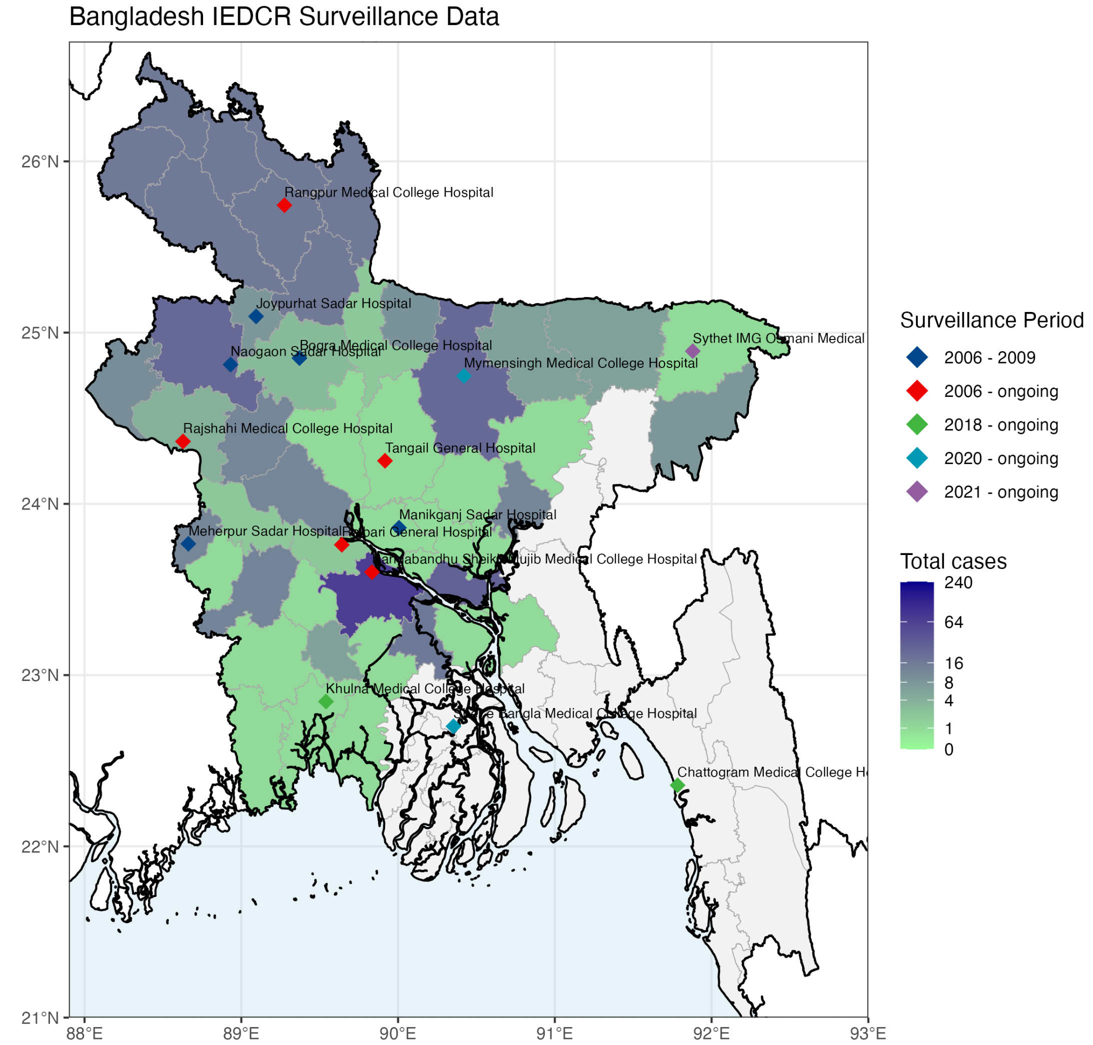

Figure B.5: Map of outbreaks (reported cases) in Bangladesh based on IEDCR Surveillance Data, with surveillance hospital sites highlighted (and period of operation).

##### B.4.1 Severity

###### Extracted outbreaks

| Study or Subgroup | Events | Total | GLMM, Fixed + Random, 95% CI |
|---|---|---|---|
| **Bangladesh** | | | |
| Luby (2006) | 11 | 12 | 0.917 [0.615; 0.998] |
| Hsu (2004) | 9 | 13 | 0.692 [0.386; 0.909] |
| Hsu (2004) | 8 | 12 | 0.667 [0.349; 0.901] |
| Gurley (2007a) | 27 | 36 | 0.750 [0.578; 0.879] |
| Kulkarni (2013) | 23 | 31 | 0.742 [0.554; 0.881] |
| Kulkarni (2013) | 3 | 7 | 0.429 [0.099; 0.816] |
| Kulkarni (2013) | 5 | 8 | 0.625 [0.245; 0.915] |
| Kulkarni (2013) | 1 | 3 | 0.333 [0.008; 0.906] |
| Kulkarni (2013) | 4 | 4 | 1.000 [0.398; 1.000] |
| Kulkarni (2013) | 7 | 7 | 1.000 [0.590; 1.000] |
| Kulkarni (2013) | 0 | 3 | 0.000 [0.000; 0.708] |
| Kulkarni (2013) | 1 | 1 | 1.000 [0.025; 1.000] |
| Kulkarni (2013) | 14 | 16 | 0.875 [0.617; 0.984] |
| Kulkarni (2013) | 40 | 44 | 0.909 [0.783; 0.975] |
| Kulkarni (2013) | 10 | 12 | 0.833 [0.516; 0.979] |
| Kulkarni (2013) | 10 | 12 | 0.833 [0.516; 0.979] |
| Satter (2023) | 0 | 1 | 0.000 [0.000; 0.975] |
| **Total (common effect, 95% CI)** | **173** | **222** | **0.779 [0.720; 0.829]** |
| **Total (random effect, 95% CI)** | | | **0.769 [0.674; 0.842]** |
| Heterogeneity: Tau$^2$ = 0.3618; Chi$^2$ = 15.59, df = 16 (P = 0.4821); I$^2$ = 0% | | | |
| **India** | | | |
| Harit (2006) | 45 | 66 | 0.682 [0.556; 0.791] |
| As (2024) | 2 | 6 | 0.333 [0.043; 0.777] |
| Yadav (2022) | 1 | 1 | 1.000 [0.025; 1.000] |
| Arankalle (2011) | 5 | 5 | 1.000 [0.478; 1.000] |
| Thomas (2019) | 16 | 18 | 0.889 [0.653; 0.986] |
| Thulaseedaran (2018) | 20 | 22 | 0.909 [0.708; 0.989] |
| **Total (common effect, 95% CI)** | **89** | **118** | **0.754 [0.669; 0.824]** |
| **Total (random effect, 95% CI)** | | | **0.812 [0.592; 0.928]** |
| Heterogeneity: Tau$^2$ = 0.8874; Chi$^2$ = 9.69, df = 5 (P = 0.0845); I$^2$ = 48.4% | | | |
| **Malaysia** | | | |
| Chua (2003) | 86 | 231 | 0.372 [0.310; 0.438] |
| Chua (2003) | 15 | 28 | 0.536 [0.339; 0.725] |
| Chua (2003) | 8 | 24 | 0.333 [0.156; 0.553] |
| **Total (common effect, 95% CI)** | **109** | **283** | **0.385 [0.330; 0.443]** |
| **Total (random effect, 95% CI)** | | | **0.385 [0.330; 0.443]** |
| Heterogeneity: Tau$^2$ = 0; Chi$^2$ = 3.03, df = 2 (P = 0.2197); I$^2$ = 34% | | | |
| **Philippines** | | | |
| Ching (2015) | 9 | 17 | 0.529 [0.278; 0.770] |
| **Singapore** | | | |
| Chua (2000b) | 1 | 11 | 0.091 [0.002; 0.413] |
| **Total (common effect, 95% CI)** | **381** | **651** | **0.585 [0.547; 0.623]** |
| **Total (random effect, 95% CI)** | | | **0.698 [0.578; 0.797]** |
| Heterogeneity: Tau$^2$ = 1.3127; Chi$^2$ = 103.10, df = 27 (P < 0.0001); I$^2$ = 73.8% | | | |
| Test for subgroup differences (common effect): Chi$^2$ = 95.85, df = 4 (P < 0.0001) | | | |
| Test for subgroup differences (random effects): Chi$^2$ = 50.30, df = 4 (P < 0.0001) | | | |

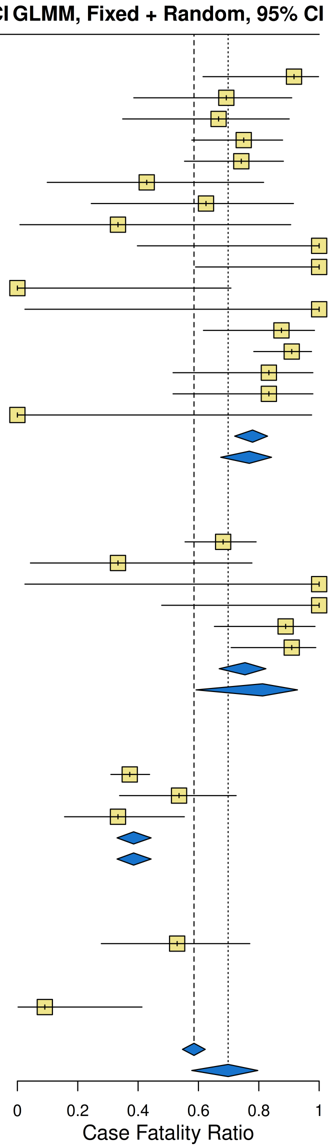

Figure B.6: Data synthesis of naïve CFR estimates by country based on extracted outbreak data using logit-transformed proportions and a generalised linear mixed-effects model. Squares represent individual CFR esimates, with black horizontal lines indicating 95% CIs; diamonds represent pooled estimates from common-effect and random-effects models, with widths corresponding to 95% CIs.

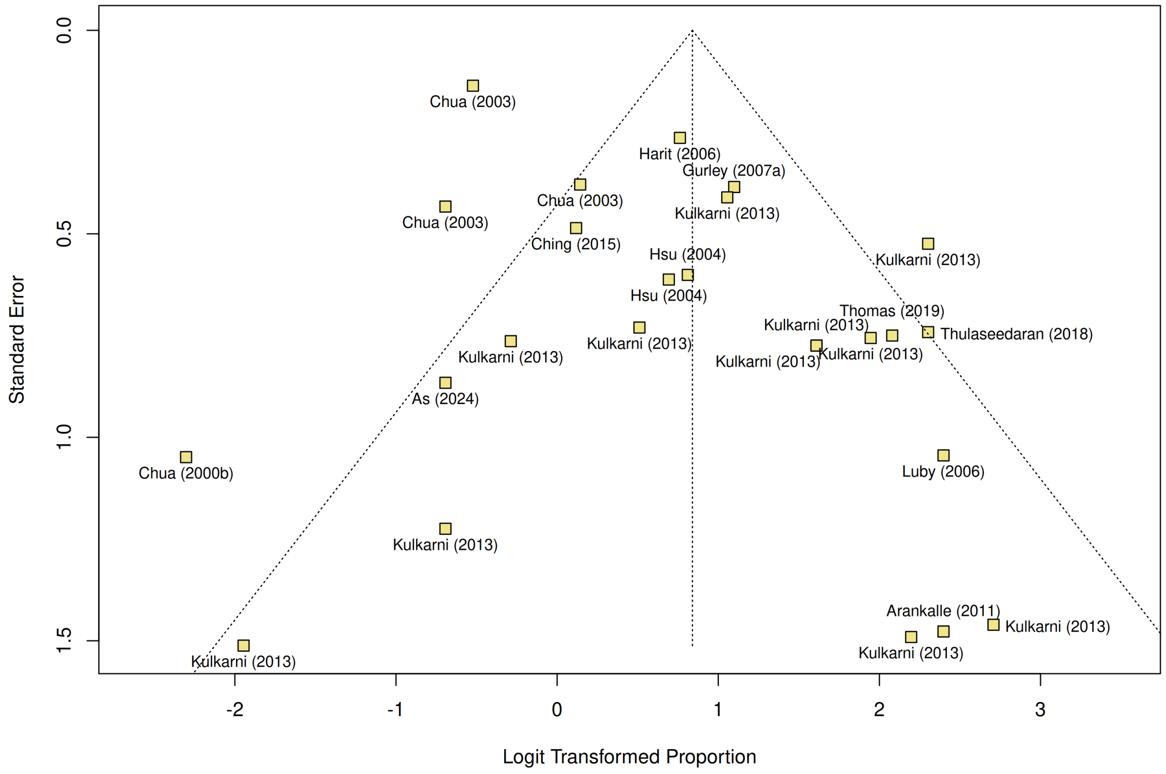

Figure B.7: Funnel plot for the deduplicated extracted outbreak CFR data synthesis. Squares represent individual study estimates. Dotted black lines represent overall pooled random effects estimate and the 95% confidence interval.

##### IEDCR surveillance

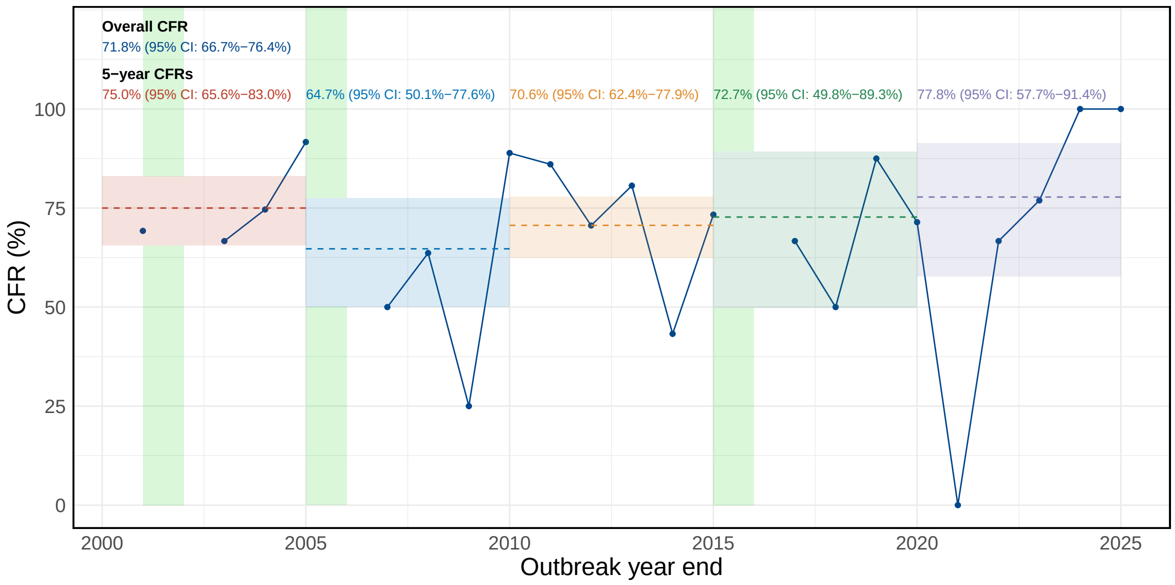

Figure B.8: Naïve CFR estimates over time based on Bangladesh IEDCR surveillance data. Each point indicates the CFR at the end of the year. Overall and 5-year naïve CFR with binomial confidence intervals estimates are annotated, and the 5-year estimates are indicated using shaded horizontal segments in a colour corresponding to the annotated segment. The vertical green segments indicate years where there were no cases were recorded (specifically 2002, 2006, and 2016).

#### Extracted parameters

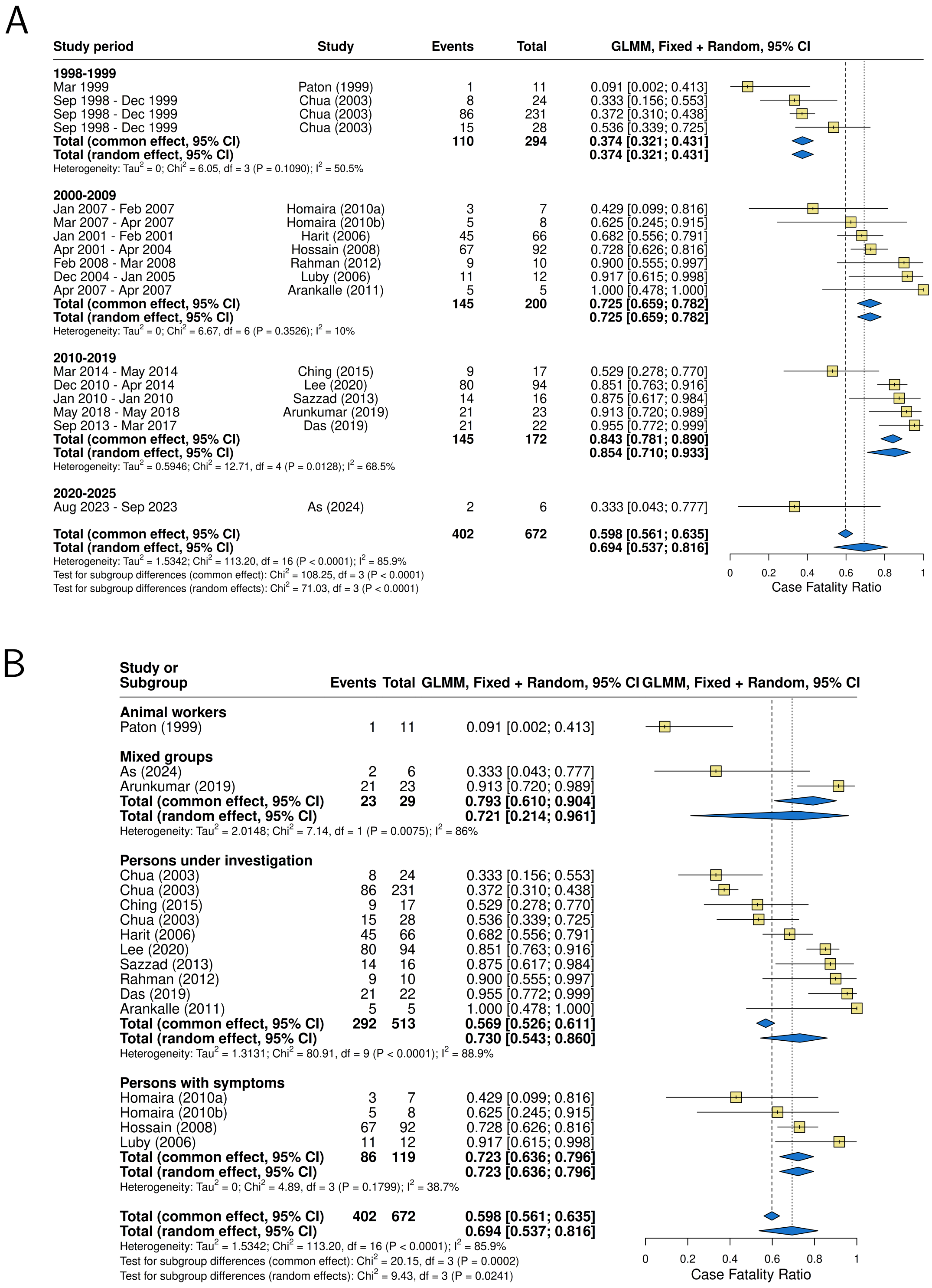

Figure B.9: Meta-analyses of deduplicated CFR estimates across (A) outbreak start year and (B) population group under study, based on extracted parameter data using logit-transformed proportions and a generalised linear mixed-effects model. Squares represent individual CFR esimates, with black horizontal lines indicating 95% CIs; diamonds represent pooled estimates from common-effect and random-effects models, with widths corresponding to 95% CIs.

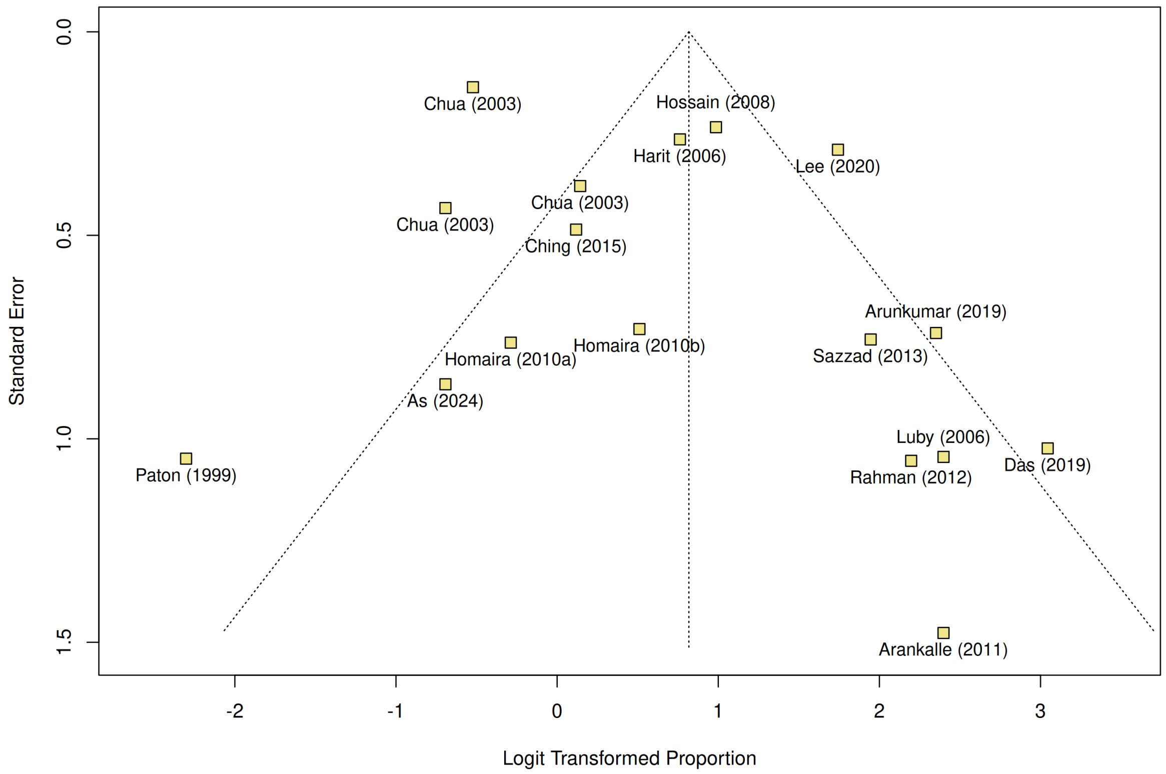

Figure B.10: Funnel plot for the deduplicated extracted parameter CFR meta-analysis. Squares represent individual study estimates. Dotted black lines represent overall pooled random effects estimate and the 95% confidence interval.

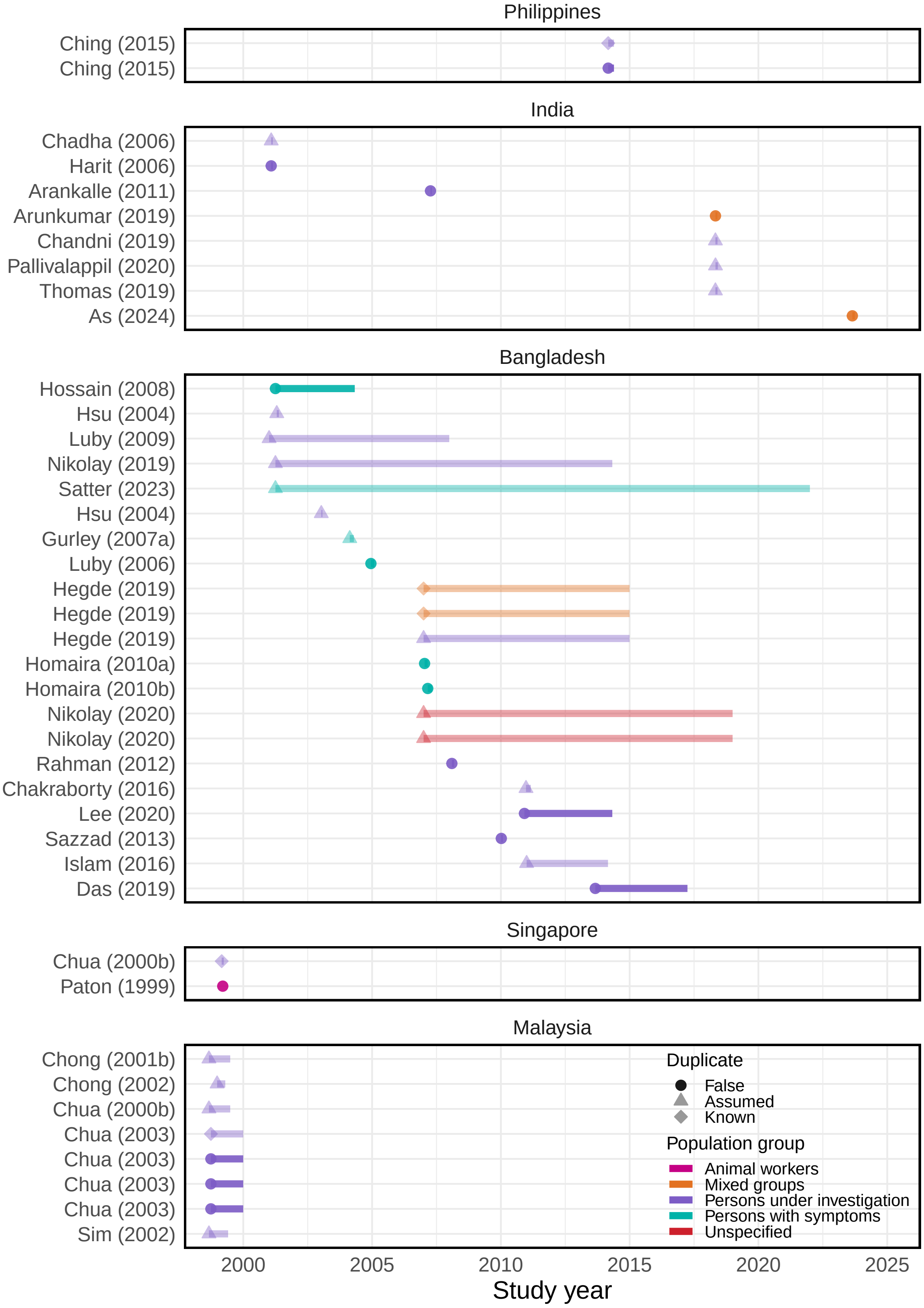

Figure B.11: Forest plot of study periods for extracted CFR parameters. Colour represents country; circles, triangles, and diamonds represent unique parameters, assumed duplicates, and known duplicates, respectively; thick coloured bands indicate the reported study period; and a single marker denotes that only a start or end date was available. Known and assumed duplicates are shown in a lighter shade. Unique parameters (circles and darker shade bands) represent the periods included in the meta-analysis of deduplicated extracted CFR estimates.

| Study or Subgroup | Events | Total | GLMM, Fixed + Random, 95% CI |
|---|---|---|---|
| **Bangladesh** | | | |
| Kulkarni (2013) | 0 | 3 | 0.000 [0.000; 0.708] |
| Hegde (2019) | 44 | 180 | 0.244 [0.184; 0.314] |
| Kulkarni (2013) | 1 | 3 | 0.333 [0.008; 0.906] |
| Homaira (2010a) | 3 | 7 | 0.429 [0.099; 0.816] |
| Kulkarni (2013) | 3 | 7 | 0.429 [0.099; 0.816] |
| Islam (2016) | 8 | 14 | 0.571 [0.289; 0.823] |
| Homaira (2010b) | 5 | 8 | 0.625 [0.245; 0.915] |
| Kulkarni (2013) | 5 | 8 | 0.625 [0.245; 0.915] |
| Kulkarni (2013) | 8 | 12 | 0.667 [0.349; 0.901] |
| Hsu (2004) | 8 | 12 | 0.667 [0.349; 0.901] |
| Kulkarni (2013) | 9 | 13 | 0.692 [0.386; 0.909] |
| Hsu (2004) | 9 | 13 | 0.692 [0.386; 0.909] |
| Kulkarni (2013) | 5 | 7 | 0.714 [0.290; 0.963] |
| Satter (2023) | 228 | 322 | 0.708 [0.655; 0.757] |
| Luby (2009) | 87 | 122 | 0.713 [0.624; 0.791] |
| Hossain (2008) | 67 | 92 | 0.728 [0.626; 0.816] |
| Kulkarni (2013) | 23 | 31 | 0.742 [0.554; 0.881] |
| Gurley (2007a) | 27 | 36 | 0.750 [0.578; 0.879] |
| Kulkarni (2013) | 27 | 36 | 0.750 [0.578; 0.879] |
| Nikolay (2019) | 193 | 248 | 0.778 [0.721; 0.828] |
| Kulkarni (2013) | 10 | 12 | 0.833 [0.516; 0.979] |
| Kulkarni (2013) | 10 | 12 | 0.833 [0.516; 0.979] |
| Lee (2020) | 80 | 94 | 0.851 [0.763; 0.916] |
| Kulkarni (2013) | 14 | 16 | 0.875 [0.617; 0.984] |
| Sazzad (2013) | 14 | 16 | 0.875 [0.617; 0.984] |
| Chakraborty (2016) | 38 | 43 | 0.884 [0.749; 0.961] |
| Rahman (2012) | 9 | 10 | 0.900 [0.555; 0.997] |
| Kulkarni (2013) | 40 | 44 | 0.909 [0.783; 0.975] |
| Kulkarni (2013) | 11 | 12 | 0.917 [0.615; 0.998] |
| Luby (2006) | 11 | 12 | 0.917 [0.615; 0.998] |
| Das (2019) | 21 | 22 | 0.955 [0.772; 0.999] |
| Kulkarni (2013) | 4 | 4 | 1.000 [0.398; 1.000] |
| Kulkarni (2013) | 1 | 1 | 1.000 [0.025; 1.000] |
| **Total (common effect, 95% CI)** | **1023** | **1472** | **0.695 [0.671; 0.718]** |
| **Total (random effect, 95% CI)** | | | **0.750 [0.681; 0.808]** |
| Heterogeneity: $Tau^2 = 0.5973$; $Chi^2 = 185.87$, df = 32 ($P < 0.0001$); $I^2 = 82.8\%$ | | | |
| **Bangladesh; India** | | | |
| Kulkarni (2013) | 221 | 292 | 0.757 [0.703; 0.805] |
| **India** | | | |
| As (2024) | 2 | 6 | 0.333 [0.043; 0.777] |
| Kulkarni (2013) | 45 | 66 | 0.682 [0.556; 0.791] |
| Harit (2006) | 45 | 66 | 0.682 [0.556; 0.791] |
| Chadha (2006) | 49 | 66 | 0.742 [0.620; 0.842] |
| Chandni (2019) | 10 | 12 | 0.833 [0.516; 0.979] |
| Thomas (2019) | 16 | 18 | 0.889 [0.653; 0.986] |
| Arunkumar (2019) | 21 | 23 | 0.913 [0.720; 0.989] |
| Pallivalappil (2020) | 21 | 23 | 0.913 [0.720; 0.989] |
| Arankalle (2011) | 5 | 5 | 1.000 [0.478; 1.000] |
| Kulkarni (2013) | 5 | 5 | 1.000 [0.478; 1.000] |
| **Total (common effect, 95% CI)** | **219** | **290** | **0.755 [0.702; 0.801]** |
| **Total (random effect, 95% CI)** | | | **0.800 [0.688; 0.879]** |
| Heterogeneity: $Tau^2 = 0.3663$; $Chi^2 = 14.84$, df = 9 ($P = 0.0954$); $I^2 = 39.4\%$ | | | |
| **Malaysia** | | | |
| Tan (2002) | 4 | 22 | 0.182 [0.052; 0.403] |
| Chua (2001) | 6 | 20 | 0.300 [0.119; 0.543] |
| Chua (2000a) | 27 | 84 | 0.321 [0.224; 0.432] |
| Chua (2003) | 8 | 24 | 0.333 [0.156; 0.553] |
| Chua (2003) | 86 | 231 | 0.372 [0.310; 0.438] |
| Chong (2001b) | 74 | 194 | 0.381 [0.313; 0.454] |
| Chua (2000b) | 105 | 265 | 0.396 [0.337; 0.458] |
| Chua (2003) | 15 | 28 | 0.536 [0.339; 0.725] |
| Sim (2002) | 11 | 18 | 0.611 [0.357; 0.827] |
| **Total (common effect, 95% CI)** | **336** | **886** | **0.379 [0.348; 0.412]** |
| **Total (random effect, 95% CI)** | | | **0.379 [0.348; 0.412]** |
| Heterogeneity: $Tau^2 = 0$; $Chi^2 = 12.28$, df = 8 ($P = 0.1393$); $I^2 = 34.8\%$ | | | |
| **Philippines** | | | |
| Ching (2015) | 9 | 17 | 0.529 [0.278; 0.770] |
| **Singapore** | | | |
| Paton (1999) | 1 | 11 | 0.091 [0.002; 0.413] |
| **Total (common effect, 95% CI)** | **1809** | **2968** | **0.610 [0.592; 0.627]** |
| **Total (random effect, 95% CI)** | | | **0.689 [0.616; 0.753]** |
| Heterogeneity: $Tau^2 = 1.0710$; $Chi^2 = 469.38$, df = 54 ($P < 0.0001$); $I^2 = 88.5\%$ | | | |
| Test for subgroup differences (common effect): $Chi^2 = 285.87$, df = 5 ($P < 0.0001$) | | | |
| Test for subgroup differences (random effects): $Chi^2 = 185.55$, df = 5 ($P < 0.0001$) | | | |

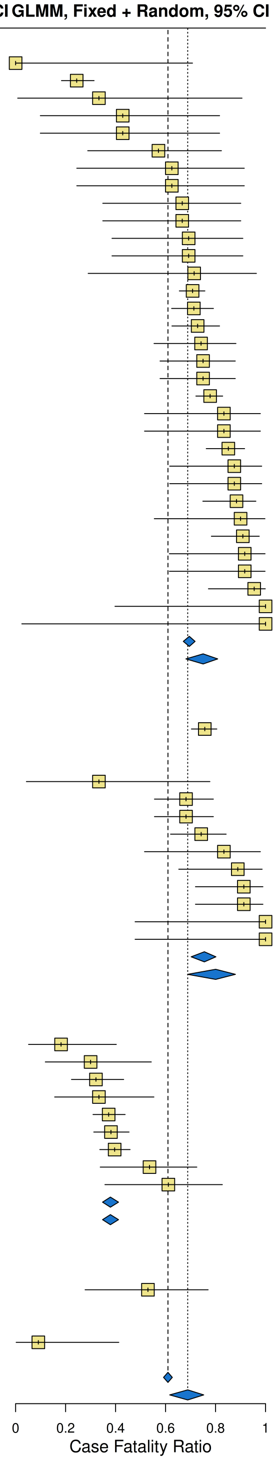

Figure B.12: Meta-analyses of CFR estimates by country based on extracted parameter data using logit-transformed proportions and a generalised linear mixed-effects model. Squares represent individual CFR esimates, with black horizontal lines indicating 95% CIs; diamonds represent pooled estimates from common-effect and random-effects models, with widths corresponding to 95% CIs. This expands on the deduplicated meta-analysis shown in Figure 3C by including assumed duplicates and excluding only known duplicates.

| Study period | Study | Events | Total | GLMM, Fixed + Random, 95% CI |
|---|---|---|---|---|
| **1998-1999** | | | | |
| Mar 1999 | Paton (1999) | 1 | 11 | 0.091 [0.002; 0.413] |
| Sep 1998 - Dec 1999 | Chua (2003) | 8 | 24 | 0.333 [0.156; 0.553] |
| Sep 1998 - Dec 1999 | Chua (2003) | 86 | 231 | 0.372 [0.310; 0.438] |
| Sep 1998 - Jun 1999 | Chong (2001b) | 74 | 194 | 0.381 [0.313; 0.454] |
| Sep 1998 - Jun 1999 | Chua (2000b) | 105 | 265 | 0.396 [0.337; 0.458] |
| Sep 1998 - Dec 1999 | Chua (2003) | 15 | 28 | 0.536 [0.339; 0.725] |
| Sep 1998 - May 1999 | Sim (2002) | 11 | 18 | 0.611 [0.357; 0.827] |
| **Total (common effect, 95% CI)** | | **300** | **771** | **0.389 [0.355; 0.424]** |
| **Total (random effect, 95% CI)** | | | | **0.389 [0.355; 0.424]** |
| Heterogeneity: $Tau^2 = 0$; $Chi^2 = 9.74$, df = 6 (P = 0.1360); $I^2 = 38.4\%$ | | | | |
| **2000-2009** | | | | |
| Jan 2007 - Feb 2007 | Homaira (2010a) | 3 | 7 | 0.429 [0.099; 0.816] |
| Mar 2007 - Apr 2007 | Homaira (2010b) | 5 | 8 | 0.625 [0.245; 0.915] |
| Jan 2003 - Jan 2003 | Hsu (2004) | 8 | 12 | 0.667 [0.349; 0.901] |
| Jan 2001 - Feb 2001 | Harit (2006) | 45 | 66 | 0.682 [0.556; 0.791] |
| Apr 2001 - May 2001 | Hsu (2004) | 9 | 13 | 0.692 [0.386; 0.909] |
| 2001 - 2007 | Luby (2009) | 87 | 122 | 0.713 [0.624; 0.791] |
| Apr 2001 - Apr 2004 | Hossain (2008) | 67 | 92 | 0.728 [0.626; 0.816] |
| Jan 2001 - Feb 2001 | Chadha (2006) | 49 | 66 | 0.742 [0.620; 0.842] |
| Feb 2004 - Apr 2004 | Gurley (2007a) | 27 | 36 | 0.750 [0.578; 0.879] |
| Apr 2001 - Apr 2014 | Nikolay (2019) | 193 | 248 | 0.778 [0.721; 0.828] |
| Feb 2008 - Mar 2008 | Rahman (2012) | 9 | 10 | 0.900 [0.555; 0.997] |
| Dec 2004 - Jan 2005 | Luby (2006) | 11 | 12 | 0.917 [0.615; 0.998] |
| Apr 2007 - Apr 2007 | Arankalle (2011) | 5 | 5 | 1.000 [0.478; 1.000] |
| **Total (common effect, 95% CI)** | | **518** | **697** | **0.743 [0.709; 0.774]** |
| **Total (random effect, 95% CI)** | | | | **0.743 [0.709; 0.774]** |
| Heterogeneity: $Tau^2 = 0$; $Chi^2 = 10.49$, df = 12 (P = 0.5729); $I^2 = 0\%$ | | | | |
| **2010-2019** | | | | |
| 2007 - 2014 | Hegde (2019) | 44 | 180 | 0.244 [0.184; 0.314] |
| Mar 2014 - May 2014 | Ching (2015) | 9 | 17 | 0.529 [0.278; 0.770] |
| Jan 2011 - Feb 2014 | Islam (2016) | 8 | 14 | 0.571 [0.289; 0.823] |
| Apr 2001 - 2021 | Satter (2023) | 228 | 322 | 0.708 [0.655; 0.757] |
| May 2018 | Chandni (2019) | 10 | 12 | 0.833 [0.516; 0.979] |
| Dec 2010 - Apr 2014 | Lee (2020) | 80 | 94 | 0.851 [0.763; 0.916] |
| Jan 2010 - Jan 2010 | Sazzad (2013) | 14 | 16 | 0.875 [0.617; 0.984] |
| Dec 2010 - Mar 2011 | Chakraborty (2016) | 38 | 43 | 0.884 [0.749; 0.961] |
| May 2018 | Thomas (2019) | 16 | 18 | 0.889 [0.653; 0.986] |
| May 2018 - May 2018 | Arunkumar (2019) | 21 | 23 | 0.913 [0.720; 0.989] |
| May 2018 - Jun 2018 | Pallivalappil (2020) | 21 | 23 | 0.913 [0.720; 0.989] |
| Sep 2013 - Mar 2017 | Das (2019) | 21 | 22 | 0.955 [0.772; 0.999] |
| **Total (common effect, 95% CI)** | | **510** | **784** | **0.651 [0.616; 0.683]** |
| **Total (random effect, 95% CI)** | | | | **0.801 [0.664; 0.892]** |
| Heterogeneity: $Tau^2 = 1.2343$; $Chi^2 = 156.04$, df = 11 (P < 0.0001); $I^2 = 93\%$ | | | | |
| **2020-2025** | | | | |
| Aug 2023 - Sep 2023 | As (2024) | 2 | 6 | 0.333 [0.043; 0.777] |
| **Unspecified** | | | | |
| Unspecified | Kulkarni (2013) | 0 | 3 | 0.000 [0.000; 0.708] |
| Unspecified | Tan (2002) | 4 | 22 | 0.182 [0.052; 0.403] |
| Unspecified | Chua (2001) | 6 | 20 | 0.300 [0.119; 0.543] |
| Unspecified | Chua (2000a) | 27 | 84 | 0.321 [0.224; 0.432] |
| Unspecified | Kulkarni (2013) | 1 | 3 | 0.333 [0.008; 0.906] |
| Unspecified | Kulkarni (2013) | 3 | 7 | 0.429 [0.099; 0.816] |
| Unspecified | Kulkarni (2013) | 5 | 8 | 0.625 [0.245; 0.915] |
| Unspecified | Kulkarni (2013) | 8 | 12 | 0.667 [0.349; 0.901] |
| Unspecified | Kulkarni (2013) | 45 | 66 | 0.682 [0.556; 0.791] |
| Unspecified | Kulkarni (2013) | 9 | 13 | 0.692 [0.386; 0.909] |
| Unspecified | Kulkarni (2013) | 5 | 7 | 0.714 [0.290; 0.963] |
| Unspecified | Kulkarni (2013) | 23 | 31 | 0.742 [0.554; 0.881] |
| Unspecified | Kulkarni (2013) | 27 | 36 | 0.750 [0.578; 0.879] |
| Unspecified | Kulkarni (2013) | 221 | 292 | 0.757 [0.703; 0.805] |
| Unspecified | Kulkarni (2013) | 10 | 12 | 0.833 [0.516; 0.979] |
| Unspecified | Kulkarni (2013) | 10 | 12 | 0.833 [0.516; 0.979] |
| Unspecified | Kulkarni (2013) | 14 | 16 | 0.875 [0.617; 0.984] |
| Unspecified | Kulkarni (2013) | 40 | 44 | 0.909 [0.783; 0.975] |
| Unspecified | Kulkarni (2013) | 11 | 12 | 0.917 [0.615; 0.998] |
| Unspecified | Kulkarni (2013) | 5 | 5 | 1.000 [0.478; 1.000] |
| Unspecified | Kulkarni (2013) | 4 | 4 | 1.000 [0.398; 1.000] |
| Unspecified | Kulkarni (2013) | 1 | 1 | 1.000 [0.025; 1.000] |
| **Total (common effect, 95% CI)** | | **479** | **710** | **0.675 [0.639; 0.708]** |
| **Total (random effect, 95% CI)** | | | | **0.688 [0.563; 0.791]** |
| Heterogeneity: $Tau^2 = 1.1126$; $Chi^2 = 96.98$, df = 21 (P < 0.0001); $I^2 = 78.3\%$ | | | | |
| **Total (common effect, 95% CI)** | | **1809** | **2968** | **0.610 [0.592; 0.627]** |
| **Total (random effect, 95% CI)** | | | | **0.689 [0.616; 0.753]** |
| Heterogeneity: $Tau^2 = 1.0710$; $Chi^2 = 469.38$, df = 54 (P < 0.0001); $I^2 = 88.5\%$ | | | | |
| Test for subgroup differences (common effect): $Chi^2 = 217.66$, df = 4 (P < 0.0001) | | | | |
| Test for subgroup differences (random effects): $Chi^2 = 192.78$, df = 4 (P < 0.0001) | | | | |

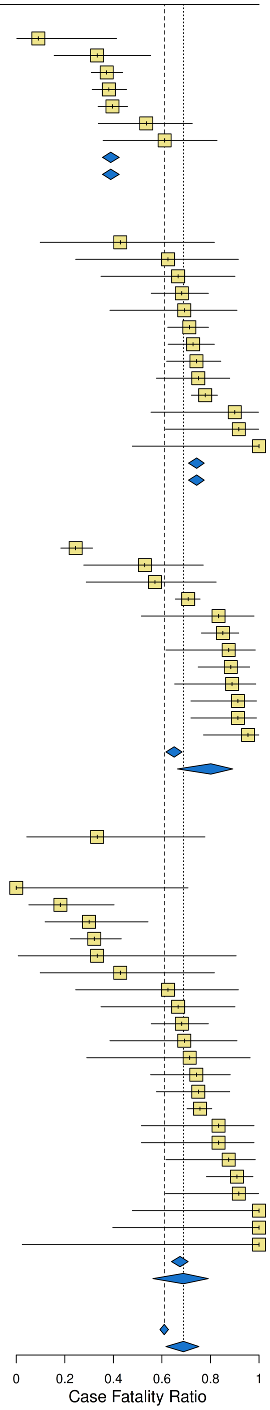

Figure B.13: Meta-analyses of CFR estimates by outbreak start year based on extracted parameter data using logit-transformed proportions and a generalised linear mixed-effects model. Squares represent individual CFR esimates, with black horizontal lines indicating 95% CIs; diamonds represent pooled estimates from common-effect and random-effects models, with widths corresponding to 95% CIs. This expands on the deduplicated meta-analysis shown in Figure B.9A by including assumed duplicates and excluding only known duplicates.

| Study or Subgroup | Events | Total | GLMM, Fixed + Random, 95% CI |
|---|---|---|---|
| **Animal workers** | | | |
| Paton (1999) | 1 | 11 | 0.091 [0.002; 0.413] |
| **Mixed groups** | | | |
| Kulkarni (2013) | 0 | 3 | 0.000 [0.000; 0.708] |
| Chua (2001) | 6 | 20 | 0.300 [0.119; 0.543] |
| Chua (2000a) | 27 | 84 | 0.321 [0.224; 0.432] |
| Kulkarni (2013) | 1 | 3 | 0.333 [0.008; 0.906] |
| As (2024) | 2 | 6 | 0.333 [0.043; 0.777] |
| Kulkarni (2013) | 3 | 7 | 0.429 [0.099; 0.816] |
| Kulkarni (2013) | 5 | 8 | 0.625 [0.245; 0.915] |
| Kulkarni (2013) | 8 | 12 | 0.667 [0.349; 0.901] |
| Kulkarni (2013) | 45 | 66 | 0.682 [0.556; 0.791] |
| Kulkarni (2013) | 9 | 13 | 0.692 [0.386; 0.909] |
| Kulkarni (2013) | 5 | 7 | 0.714 [0.290; 0.963] |
| Kulkarni (2013) | 23 | 31 | 0.742 [0.554; 0.881] |
| Kulkarni (2013) | 27 | 36 | 0.750 [0.578; 0.879] |
| Kulkarni (2013) | 221 | 292 | 0.757 [0.703; 0.805] |
| Kulkarni (2013) | 10 | 12 | 0.833 [0.516; 0.979] |
| Kulkarni (2013) | 10 | 12 | 0.833 [0.516; 0.979] |
| Kulkarni (2013) | 14 | 16 | 0.875 [0.617; 0.984] |
| Kulkarni (2013) | 40 | 44 | 0.909 [0.783; 0.975] |
| Arunkumar (2019) | 21 | 23 | 0.913 [0.720; 0.989] |
| Kulkarni (2013) | 11 | 12 | 0.917 [0.615; 0.998] |
| Kulkarni (2013) | 5 | 5 | 1.000 [0.478; 1.000] |
| Kulkarni (2013) | 4 | 4 | 1.000 [0.398; 1.000] |
| Kulkarni (2013) | 1 | 1 | 1.000 [0.025; 1.000] |
| **Total (common effect, 95% CI)** | **498** | **717** | **0.695 [0.660; 0.727]** |
| **Total (random effect, 95% CI)** | | | **0.715 [0.606; 0.804]** |
| Heterogeneity: $Tau^2 = 0.9032$; $Chi^2 = 88.22$, df = 22 ($P < 0.0001$); $I^2 = 75.1\%$ | | | |
| **Persons under investigation** | | | |
| Tan (2002) | 4 | 22 | 0.182 [0.052; 0.403] |
| Hegde (2019) | 44 | 180 | 0.244 [0.184; 0.314] |
| Chua (2003) | 8 | 24 | 0.333 [0.156; 0.553] |
| Chua (2003) | 86 | 231 | 0.372 [0.310; 0.438] |
| Chong (2001b) | 74 | 194 | 0.381 [0.313; 0.454] |
| Chua (2000b) | 105 | 265 | 0.396 [0.337; 0.458] |
| Ching (2015) | 9 | 17 | 0.529 [0.278; 0.770] |
| Chua (2003) | 15 | 28 | 0.536 [0.339; 0.725] |
| Islam (2016) | 8 | 14 | 0.571 [0.289; 0.823] |
| Sim (2002) | 11 | 18 | 0.611 [0.357; 0.827] |
| Hsu (2004) | 8 | 12 | 0.667 [0.349; 0.901] |
| Harit (2006) | 45 | 66 | 0.682 [0.556; 0.791] |
| Hsu (2004) | 9 | 13 | 0.692 [0.386; 0.909] |
| Luby (2009) | 87 | 122 | 0.713 [0.624; 0.791] |
| Chadha (2006) | 49 | 66 | 0.742 [0.620; 0.842] |
| Nikolay (2019) | 193 | 248 | 0.778 [0.721; 0.828] |
| Chandni (2019) | 10 | 12 | 0.833 [0.516; 0.979] |
| Lee (2020) | 80 | 94 | 0.851 [0.763; 0.916] |
| Sazzad (2013) | 14 | 16 | 0.875 [0.617; 0.984] |
| Chakraborty (2016) | 38 | 43 | 0.884 [0.749; 0.961] |
| Thomas (2019) | 16 | 18 | 0.889 [0.653; 0.986] |
| Rahman (2012) | 9 | 10 | 0.900 [0.555; 0.997] |
| Pallivalappil (2020) | 21 | 23 | 0.913 [0.720; 0.989] |
| Das (2019) | 21 | 22 | 0.955 [0.772; 0.999] |
| Arankalle (2011) | 5 | 5 | 1.000 [0.478; 1.000] |
| **Total (common effect, 95% CI)** | **969** | **1763** | **0.550 [0.526; 0.573]** |
| **Total (random effect, 95% CI)** | | | **0.682 [0.571; 0.775]** |
| Heterogeneity: $Tau^2 = 1.1786$; $Chi^2 = 296.87$, df = 24 ($P < 0.0001$); $I^2 = 91.9\%$ | | | |
| **Persons with symptoms** | | | |
| Homaira (2010a) | 3 | 7 | 0.429 [0.099; 0.816] |
| Homaira (2010b) | 5 | 8 | 0.625 [0.245; 0.915] |
| Satter (2023) | 228 | 322 | 0.708 [0.655; 0.757] |
| Hossain (2008) | 67 | 92 | 0.728 [0.626; 0.816] |
| Gurley (2007a) | 27 | 36 | 0.750 [0.578; 0.879] |
| Luby (2006) | 11 | 12 | 0.917 [0.615; 0.998] |
| **Total (common effect, 95% CI)** | **341** | **477** | **0.715 [0.673; 0.754]** |
| **Total (random effect, 95% CI)** | | | **0.715 [0.673; 0.754]** |
| Heterogeneity: $Tau^2 = 0$; $Chi^2 = 5.17$, df = 5 ($P = 0.3956$); $I^2 = 3.3\%$ | | | |
| **Total (common effect, 95% CI)** | **1809** | **2968** | **0.610 [0.592; 0.627]** |
| **Total (random effect, 95% CI)** | | | **0.689 [0.616; 0.753]** |
| Heterogeneity: $Tau^2 = 1.0710$; $Chi^2 = 469.38$, df = 54 ($P < 0.0001$); $I^2 = 88.5\%$ | | | |
| Test for subgroup differences (common effect): $Chi^2 = 76.60$, df = 3 ($P < 0.0001$) | | | |
| Test for subgroup differences (random effects): $Chi^2 = 9.63$, df = 3 ($P = 0.0220$) | | | |

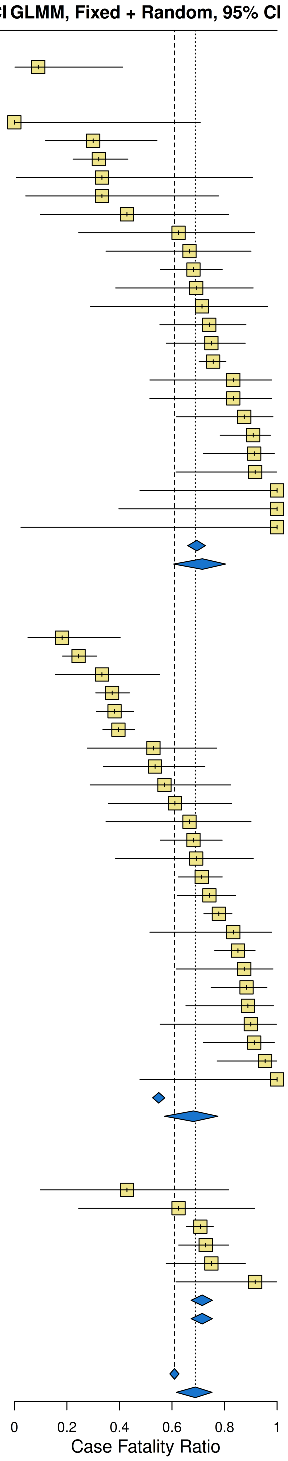

Figure B.14: Meta-analyses of CFR estimates by population group under study based on extracted parameter data using logit-transformed proportions and a generalised linear mixed-effects model. Squares represent individual CFR esimates, with black horizontal lines indicating 95% CIs; diamonds represent pooled estimates from common-effect and random-effects models, with widths corresponding to 95% CIs. This expands on the deduplicated meta-analysis shown in Figure B.9B by including assumed duplicates and excluding only known duplicates.

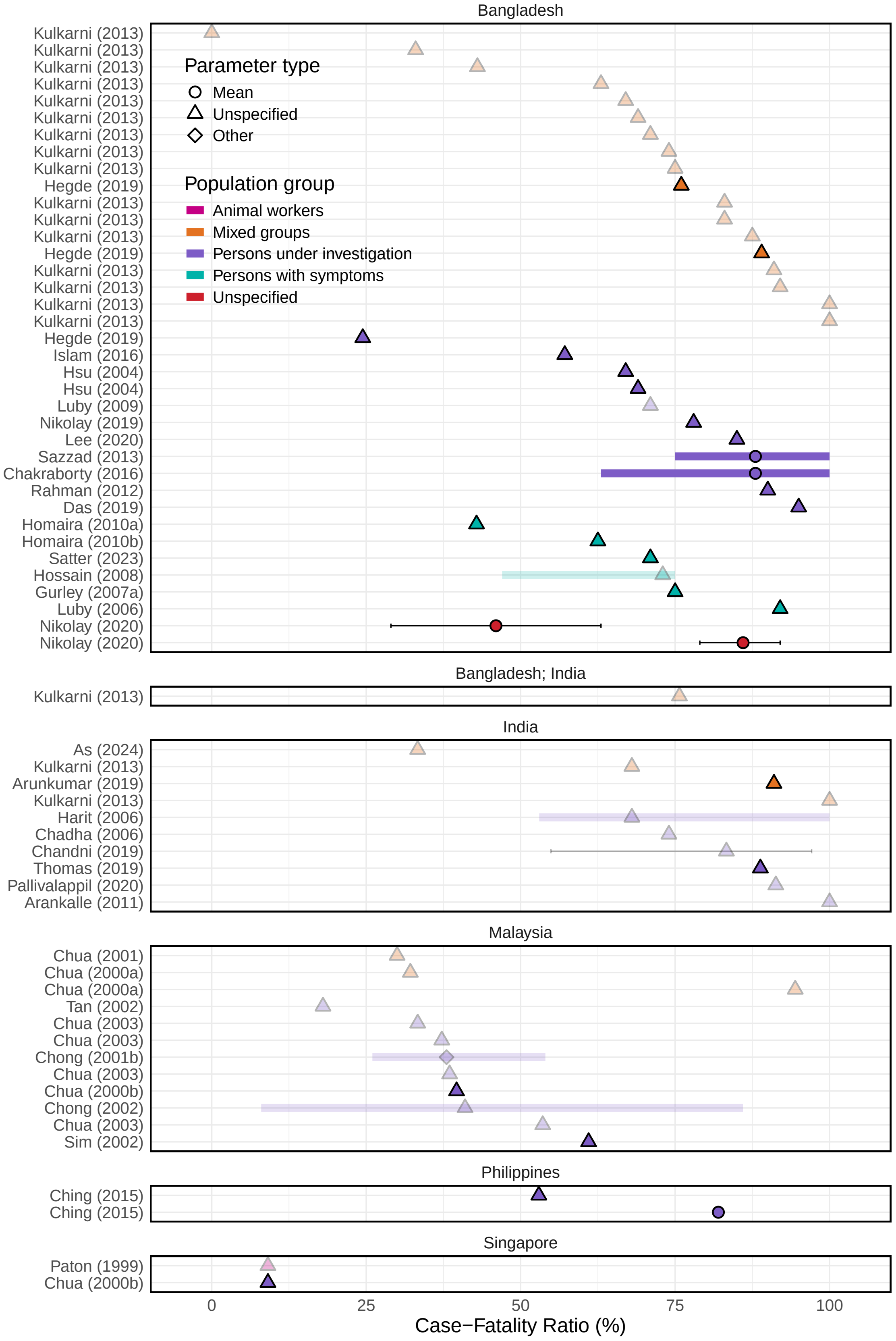

Figure B.15: CFR parameters extracted across all studies, including known and assumed duplicates stratified by country. Colours represent population group under study; parameters with a QA score $\leq 50\%$ have a lighter shade; circles, triangles, and diamonds represent mean, unspecified, and other respectively estimate types respectively; black error bars indicate estimate uncertainty, and thick coloured bands indicate ranges across central estimates of disaggregated parameters (e.g. age, sex, time).

##### B.4.2 Risk factor plots

In this section we plot the reported risk factors for different epidemiological outcomes. This includes both significant, not significant, and unspecified risk factors. We display risk factors for the following specific outcomes: infection, serology, death, and spillover risk. All other risk factors were grouped into an other category. Note, neurological symptoms were collected as a specific outcome but were grouped in the other category because only a single risk factor was extracted for this outcome.

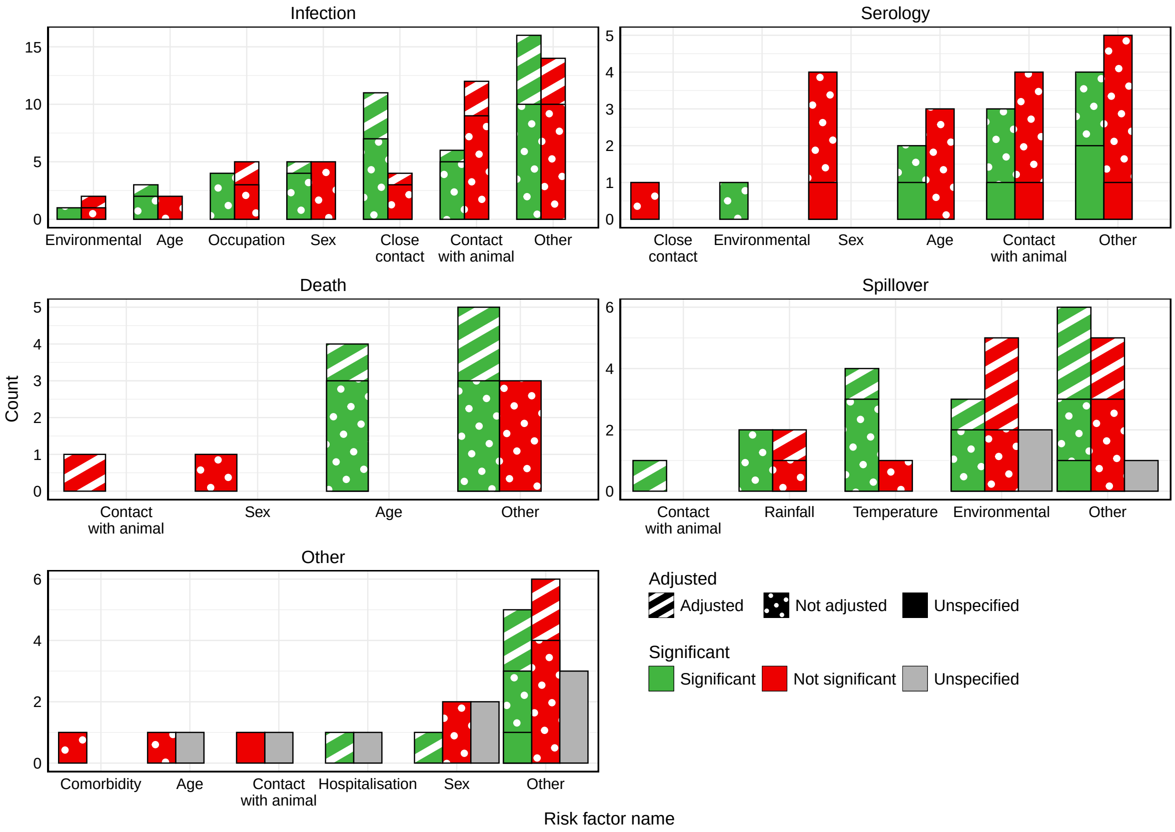

Figure B.16: Tally of reported significant, non significant, and unspecified risk factors associated with infection, serology, death, spillover, and other outcomes. Colours indicate significance and patterns indicate whether the analysis was adjusted.

#### C Other Systematic Reviews

Systematic reviews were excluded from our analysis, but the systematic reviews that were identified in the database search and used for validation purposes are presented in Table C.16.

| Study | Title | Journal | DOI |
|---|---|---|---|
| Glennon 2018 | Domesticated animals as hosts of henipaviruses and filoviruses: A systematic review. | The Veterinary Journal | 10.1016/j.tvjl.2017.12.024 |
| Hegde 2024 | Potential for Person-to-Person Transmission of Henipaviruses: A Systematic Review of the Literature. | The Journal of Infectious Diseases | 10.1093/infdis/jiad467 |
| Kenmoe 2019 | Case fatality rate and risk factors for Nipah virus encephalitis: A systematic review and meta-analysis. | Journal of Clinical Virology | 10.1016/j.jcv.2019.05.009 |
| Vasudevan 2024 | Global and regional mortality statistics of nipah virus from 1994 to 2023: a comprehensive systematic review and meta-analysis. | Pathogens and Global Health | 10.1080/20477724.2024.2380131 |
| Khan 2022 | Major bat-borne zoonotic viral epidemics in Asia and Africa: A systematic review and meta-analysis. | Veterinary Medicine and Science | 10.1002/vms3.835 |
| Suman 2024 | NIPAH Virus Encephalitis: Unveiling the Epidemiology, Risk Factors, and Clinical Outcomes - A Systematic Review and Meta-Analysis. | Journal of Pharmacy and Bioallied Sciences | 10.4103/jpbs.jpbs_935_23 |
| Nguyen 2024 | Emerging zoonotic diseases in Southeast Asia in the period 2011-2022: a systematic literature review. | Veterinary Quarterly | 10.1080/01652176.2023.2300965 |
| Bhattacharya 2020 | Detailed Molecular Biochemistry for Novel Therapeutic Design Against Nipah and Hendra Virus: A Systematic Review. | Current Molecular Pharmacology | 10.2174/1874467212666191023123732 |
| Shrestha 2020 | Non-malarial febrile illness: a systematic review of published aetiological studies and case reports from Southern Asia and South-eastern Asia, 1980-2015. | BMC Medicine | 10.1186/s12916-020-01745-0 |
| Tan 2024 | A systematic review on Nipah virus: global molecular epidemiology and medical countermeasures development. | Virus Evolution | 10.1093/ve/veae048 |
| Rees 2021 | Transmission modelling of environmentally persistent zoonotic diseases: a systematic review | The Lancet Planetary Health | 10.1016/S2542-5196(21)00137-6 |
| Sun 2024 | Mapping the distribution of Nipah virus infections: a geospatial modelling analysis. | The Lancet Planetary Health | 10.1016/S2542-5196(24)00119-0 |
| Riccò 2025 | Risk of Nipah Virus Seroprevalence in Healthcare Workers: A Systematic Review with Meta-Analysis | Viruses | 10.3390/v17010081 |
| Alla 2024 | A systematic review of case reports on mortality, modes of infection, diagnostic tests, and treatments for Nipah virus infection | Medicine | 10.1097/MD.0000000000039989 |

Table C.16: Other reviews of Nipah identified during manuscript screening.

#### D Excluded Studies

We list all excluded studies with reasons for exclusion in the Priority Pathogens GitHub repo. The full list can be access at https://github.com/mrc-ide/priority-pathogens/blob/main/data/Nipah_excluded_studies.csv

### E Software and epireview

In parallel with this work, we developed an R package called `epireview` to provide a central location to host and access the extracted data for the nine priority pathogens in our review series. The package allows for submissions of outbreak, model, and parameter data from new peer-reviewed papers via pull requests, and includes functions to produce the tables and figures included in this paper and update them with any additional data. This package will be updated as the overall project by the Pathogen Epidemiology Review Group (PERG) continues to extract modelling parameters for the rest of the nine priority pathogens as defined by WHO.

There are several vignettes available:

- A vignette that lists the options for each outbreak, model, or parameter field and describes how to access them using a function in the package.
- A vignette to explain the process of updating the database with new article, outbreak, model, or parameter data.

Vignettes for Marburg Virus Disease, Lassa fever, SARS-CoV-1, and Zika with tables and figures from the respective papers are available. These will be updated as data are added to the database.

We have provided a csv file with all article information of the papers which we extract and provide the below code to generate this in `R` from `epireview` (see Supplement E).

```
require(tidyverse)

is_epireview_available <- require("epireview")

if(!is_epireview_available)
{
is_remotes_available <- require("remotes")

if(!is_remotes_available) install.packages("remotes")

remotes::install_github("mrc-ide/epireview")
}

nipah <- epireview::load_epidata("nipah")
```

### F PRISMA 2020 Checklists

| Section & Topic | Item # | Checklist item | Reported (Yes/No) |
|---|---|---|---|
| **Title** | | | |
| Title | 1 | Identify the report as a systematic review. | Yes |
| **Background** | | | |
| Objectives | 2 | Provide an explicit statement of the main objective(s) or question(s) the review addresses. | Yes |
| **Methods** | | | |
| Eligibility criteria | 3 | Specify the inclusion and exclusion criteria for the review. | Yes |
| Information sources | 4 | Specify the information sources (e.g. databases, registers) used to identify studies and the date when each was last searched. | Yes |
| Risk of bias | 5 | Specify the methods used to assess risk of bias in the included studies. | Yes |
| Synthesis of results | 6 | Specify the methods used to present and synthesise results. | Yes |
| **Results** | | | |
| Included studies | 7 | Give the total number of included studies and participants and summarise relevant characteristics of studies. | Yes |
| Synthesis of results | 8 | Present results for main outcomes, preferably indicating the number of included studies and participants for each. If meta-analysis was done, report the summary estimate and confidence/credible interval. If comparing groups, indicate the direction of the effect (i.e. which group is favoured). | Yes |
| **Discussion** | | | |
| Limitations of evidence | 9 | Provide a brief summary of the limitations of the evidence included in the review (e.g. study risk of bias, inconsistency and imprecision). | Yes |
| Interpretation | 10 | Provide a general interpretation of the results and important implications. | Yes |
| **Other** | | | |
| Funding | 11 | Specify the primary source of funding for the review. | Yes |
| Registration | 12 | Provide the register name and registration number. | Yes |

Table F.17: PRISMA 2020 Abstracts Checklist. ([15])

| Section & Topic | Item # | Checklist item | Location where item is reported |
|---|---|---|---|
| **Title** | | | |
| Title | 1 | Identify the report as a systematic review. | page 1 |
| **Abstract** | | | |
| Abstract | 2 | See the PRISMA 2020 for Abstracts checklist. | Table F.17 |
| **Introduction** | | | |
| Rationale | 3 | Describe the rationale for the review in the context of existing knowledge. | page 2 |
| Objectives | 4 | Provide an explicit statement of the objective(s) or question(s) the review addresses. | page 2/3 |
| **Methods** | | | |
| Eligibility criteria | 5 | Specify the inclusion and exclusion criteria for the review and how studies were grouped for the syntheses. | page 3 |

Continued on next page

Continued from previous page

| Section & Topic | Item # | Checklist item | Location where item is reported |
|---|---|---|---|
| Information sources | 6 | Specify all databases, registers, websites, organisations, reference lists and other sources searched or consulted to identify studies. Specify the date when each source was last searched or consulted. | page 3 |
| Search strategy | 7 | Present the full search strategies for all databases, registers and websites, including any filters and limits used. | page 3 + Figure 1 |
| Selection process | 8 | Specify the methods used to decide whether a study met the inclusion criteria of the review, including how many reviewers screened each record and each report retrieved, whether they worked independently, and if applicable, details of automation tools used in the process. | page 3 |
| Data collection process | 9 | Specify the methods used to collect data from reports, including how many reviewers collected data from each report, whether they worked independently, any processes for obtaining or confirming data from study investigators, and if applicable, details of automation tools used in the process. | page 3/4 |
| Data items | 10a | List and define all outcomes for which data were sought. Specify whether all results that were compatible with each outcome domain in each study were sought (e.g. for all measures, time points, analyses), and if not, the methods used to decide which results to collect. | page 3/4 |
| | 10b | List and define all other variables for which data were sought (e.g. participant and intervention characteristics, funding sources). Describe any assumptions made about any missing or unclear information. | page 3/4 |
| Study risk of bias assessment | 11 | Specify the methods used to assess risk of bias in the included studies, including details of the tool(s) used, how many reviewers assessed each study and whether they worked independently, and if applicable, details of automation tools used in the process. | page 4 |
| Effect measures | 12 | Specify for each outcome the effect measure(s) (e.g. risk ratio, mean difference) used in the synthesis or presentation of results. | page 3/4 |
| Synthesis methods | 13a | Describe the processes used to decide which studies were eligible for each synthesis (e.g. tabulating the study intervention characteristics and comparing against the planned groups for each synthesis (item #5)). | page 4 |
| | 13b | Describe any methods required to prepare the data for presentation or synthesis, such as handling of missing summary statistics, or data conversions. | page 4 |
| | 13c | Describe any methods used to tabulate or visually display results of individual studies and syntheses. | page 4 |
| | 13d | Describe any methods used to synthesize results and provide a rationale for the choice(s). If meta-analysis was performed, describe the model(s), method(s) to identify the presence and extent of statistical heterogeneity, and software package(s) used. | page 4 |
| | 13e | Describe any methods used to explore possible causes of heterogeneity among study results (e.g. subgroup analysis, meta-regression). | - |
| | 13f | Describe any sensitivity analyses conducted to assess robustness of the synthesized results. | - |
| Reporting bias assessment | 14 | Describe any methods used to assess risk of bias due to missing results in a synthesis (arising from reporting biases). | page 4 |
| Certainty assessment | 15 | Describe any methods used to assess certainty (or confidence) in the body of evidence for an outcome. | page 4 |
| **Results** | | | |
| Study selection | 16a | Describe the results of the search and selection process, from the number of records identified in the search to the number of studies included in the review, ideally using a flow diagram. | page 4 |
| | 16b | Cite studies that might appear to meet the inclusion criteria, but which were excluded, and explain why they were excluded. | Table ?? |
| Study characteristics | 17 | Cite each included study and present its characteristics. | pages 4-13 |

Continued on next page

Continued from previous page

| Section & Topic | Item # | Checklist item | Location where item is reported |
|---|---|---|---|
| Risk of bias in studies | 18 | Present assessments of risk of bias for each included study. | page 7 |
| Results of individual studies | 19 | For all outcomes, present, for each study: (a) summary statistics for each group (where appropriate) and (b) an effect estimate and its precision (e.g. confidence/credible interval), ideally using structured tables or plots. | pages 4-13 |
| Results of syntheses | 20a | For each synthesis, briefly summarise the characteristics and risk of bias among contributing studies. | page 6/7 |
| | 20b | Present results of all statistical syntheses conducted. If meta-analysis was done, present for each the summary estimate and its precision (e.g. confidence/credible interval) and measures of statistical heterogeneity. If comparing groups, describe the direction of the effect. | page 4-13 |
| | 20c | Present results of all investigations of possible causes of heterogeneity among study results. | page 4-13 |
| | 20d | Present results of all sensitivity analyses conducted to assess the robustness of the synthesized results. | page 4-13 |
| Reporting biases | 21 | Present assessments of risk of bias due to missing results (arising from reporting biases) for each synthesis assessed. | page 7 |
| Certainty of evidence | 22 | Present assessments of certainty (or confidence) in the body of evidence for each outcome assessed. | pages 4-13 |
| **Discussion** | | | |
| Discussion | 23a | Provide a general interpretation of the results in the context of other evidence. | page 14/15 |
| | 23b | Discuss any limitations of the evidence included in the review. | page 14-15 |
| | 23c | Discuss any limitations of the review processes used. | page 14-15 |
| | 23d | Discuss implications of the results for practice, policy, and future research. | page 14-15 |
| **Other Information** | | | |
| Registration and protocol | 24a | Provide registration information for the review, including register name and registration number, or state that the review was not registered. | page 14 |
| | 24b | Indicate where the review protocol can be accessed, or state that a protocol was not prepared. | page 16 |
| | 24c | Describe and explain any amendments to information provided at registration or in the protocol. | n/a |
| Support | 25 | Describe sources of financial or non-financial support for the review, and the role of the funders or sponsors in the review. | page 15/16 |
| Competing interests | 26 | Declare any competing interests of review authors. | page 16 |
| Availability of data, code and other materials | 27 | Report which of the following are publicly available and where they can be found: template data collection forms; data extracted from included studies; data used for all analyses; analytic code; any other materials used in the review. | page 16 |

Table F.18: PRISMA 2020 Checklist. ([15])

#### G Pathogen Epidemiology Review Group (PERG) Membership

| First name | Surname | Affiliation |
|---|---|---|
| Aaron | Morris | University of Oxford |
| Alpha | Forna | University of Georgia |
| Amy | Dighe | Johns Hopkins |
| Anna | Vicco | Imperial College London |
| Anna-Maria | Hartner | Robert Koch Institute |
| Anne | Cori | Imperial College London |
| Arran | Hamlet | Imperial College London |
| Ben | Lambert | University of Oxford |
| Bethan | Cracknell Daniels | Imperial College London |
| Charles | Whittaker | Imperial College London |
| Christian | Morgenstern | Imperial College London |
| Cosmo | Santoni | Imperial College London |
| Cyril | Geismar | Imperial College London |
| Dariya | Nikitin | Imperial College London |
| David | Jorgensen | Imperial College London |
| Dominic | Dee | Imperial College London |
| Ed | Knock | Imperial College London |
| Gina | Cuomo-Dannenburg | Imperial College London |
| Hayley | Thompson | PATH |
| Ilaria | Dorigatti | Imperial College London |
| Isobel | Routledge | UCSF |
| Jack | Wardle | Imperial College London |
| Janetta | Skarp | Imperial College London |
| Joseph | Hicks | Imperial College London |
| Juliette | Unwin | University of Bristol |
| Kanchan | Parchani | Imperial College London |
| Keith | Fraser | Imperial College London |
| Kelly | Charniga | Imperial College London |
| Kelly | McCain | Imperial College London |
| Kieran | Drake | Imperial College London |
| Lily | Geidelberg | Imperial College London |
| Lorenzo | Cattarino | UKHSA |
| Mantra | Kusumgar | Imperial College London |
| Mara | Kont | Imperial College London |
| Marc | Baguelin | Imperial College London |
| Natsuko | Imai-Eaton | Imperial College London |
| Pablo | Perez Guzman | Imperial College London |
| Patrick | Doohan | Imperial College London |
| Paul | Lietar | Imperial College London |

Continued on next page

Continued from previous page

| First name | Surname | Affiliation |
|---|---|---|
| Paula | Christen | Imperial College London |
| Rebecca | Nash | Imperial College London |
| Rhys | Earl | National University of Singapore |
| Rich | Fitzjohn | Imperial College London |
| Richard | Sheppard | Imperial College London |
| Rob | Johnson | Imperial College London |
| Ruth | McCabe | Imperial College London |
| Sabine | van Elsland | Imperial College London |
| Sangeeta | Bhatia | Imperial College London |
| Sequoia | Leuba | Imperial College London |
| Shazia | Ruybal-Pesantez | Imperial College London |
| Sreejith | Radhakrishnan | UKHSA |
| Thomas | Rawson | Imperial College London |
| Tristan | Naidoo | Imperial College London |
| Zulma | Cucunuba Perez | Pontificia Universidad Javeriana |

Table G.19: Pathogen Epidemiology Review Group (PERG) membership as of Janaury 2025 (when extractions for NiV began).
